# Household contact tracing with intensified tuberculosis and HIV screening in South Africa: a cluster randomised trial

**DOI:** 10.1101/2021.10.21.21265356

**Authors:** Neil A Martinson, Limakatso Lebina, Emily L Webb, Andrew Ratsela, Ebrahim Varavia, Anthony Kinghorn, Sanjay G Lala, Jonathan E. Golub, Zama Bosch, Kegaugetswe P Motsomi, Peter MacPherson

**Affiliations:** Perinatal HIV Research Unit (PHRU), University of the Witwatersrand, Johannesburg, South Africa; Johns Hopkins University Center for TB Research, Baltimore, MD; MRC International Statistics and Epidemiology Group, London School of Hygiene and Tropical Medicine, UK; Department of Internal Medicine, University of Limpopo, Polokwane, South Africa; Department of Internal Medicine. Klerksdorp Tshepong Hospital Complex, North West Provincial Department of Health, and University of the Witwatersrand, South Africa; Department of Paediatrics and Child Health, University of the Witwatersrand, South Africa; Department of Clinical Sciences, Liverpool School of Tropical Medicine, UK; Malawi-Liverpool-Wellcome Trust Clinical Research Programme, Malawi; Clinical Research Department, London School of Hygiene and Tropical Medicine

**Author notes:** **Corresponding Author** Dr Peter MacPherson, Liverpool School of Tropical Medicine, Pembroke Place, Liverpool, United Kingdom, L3 5QA, 44. **Alternate Corresponding Author** Prof Neil Martinson, Perinatal HIV Research Unit, Chris Baragwanath Hospital, Soweto, South Africa.

**Keywords:** Tuberculosis, HIV, screening, diagnosis, contract tracing, randomised controlled trials

## Abstract

**Background:** Household contact tracing for tuberculosis (TB) may facilitate TB diagnosis and identify individuals who may benefit from TB preventive therapy (TPT). In this cluster-randomised trial, we investigated whether household contact tracing and intensive TB/HIV screening would improve TB-free survival.

**Methods:** Household contacts of index TB patients in two Provinces of South Africa were randomised to home tracing and intensive HIV/TB screening (sputum Xpert and culture; HIV testing with treatment linkage; and TPT, if eligible), or standard of care (SOC, clinic referral letters). The primary outcome was incident TB or death at 15-months. Secondary outcomes included tuberculin skin test (TST) positivity in children ≤14 years and undiagnosed HIV. (<u>ISRCTN16006202</u>).

**Results:** From December 2016-March 2019, 1,032 index patients (4,459 contacts) and 1,030 (4,129 contacts) were randomised to the intervention and SOC arms. 3.2% (69/2166) of intervention arm contacts had prevalent microbiologically-confirmed TB. At 15-months, the cumulative incidence of TB or death did not differ between the intensive screening (93/3230, 2.9%) and SOC (80/2600, 3.1%) arms (hazard ratio: 0.90, 95% confidence interval (CI): 0.66-1.24). TST positivity was higher in the intensive screening arm (38/845, 4.5%) compared to the SOC arm (15/800, 1.9%, odds ratio: 2.25, 95% CI: 1.07-4.72). Undiagnosed HIV was similar between arms (41/3185, 1.3% vs. 32/2543, 1.3%; odds ratio: 1.02, 95% CI: 0.64-1.64).

**Conclusions:** Household contact tracing with intensive screening and referral did not reduce incident TB or death. Providing referral letters to household contacts of index patients is an alternative strategy to home visits in high TB/HIV-prevalence settings.

**Author Summary:** In South Africa, household contacts of TB cases received referral letters or home-tracing with intensified TB/HIV screening. At 15-months, the cumulative incidence of TB or death did not differ between the intensive screening (93/3230, 2.9%) and SOC (80/2600, 3.1%) arms.

## Introduction

Contact tracing of people with tuberculosis (TB) has been advocated as part of TB control for many years [1–3] because it facilitates early diagnosis and treatment of infectious individuals and identifies those who could benefit from TB preventive treatment (TPT) [4]. Although World Health Organization (WHO) and numerous national guidelines recommend household TB contact tracing, these have not been widely implemented in high TB burden countries because of limited effectiveness data [5,6]. The COVID-19 pandemic has severely impacted TB care and prevention programmes in sub-Saharan Africa [7], and may have reversed recent improvements in TB diagnosis and treatment [8,9].

Previous randomised trials have investigated the effectiveness of TB household contact tracing interventions on screening completion, community TB prevalence, and TB notification, showing mixed results [10–13]. Evidence suggests more intensive TB screening approaches increase diagnostic yield and could reduce transmission by identifying and treating people with subclinical infectious TB earlier [5,14,15]. Moreover, universal HIV testing with immediate initiation of antiretroviral therapy (ART) together with TPT, reduces morbidity, mortality, and incident TB disease among people living with HIV [16–18].

We hypothesized that household contact tracing with intensive screening for TB and HIV with supported linkage to treatment and home initiation of TPT could result in earlier TB and HIV diagnosis and treatment, and reduce TB transmission.

## Methods

### Study design and participants

We conducted an open two-arm cluster randomised trial of household contact tracing and intensive TB/HIV screening in South Africa (<u>ISRCTN16006202</u>). Methods have been described previously (S1 Protocol) [19]. In brief, we recruited index TB patients diagnosed at two South African sites with large differences in annual TB incidence and HIV prevalence (Mangaung, Free State [20]; Capricorn, Limpopo [21,22]). During the study period there were few programmatic attempts made to identify and screen household contacts for TB.

Study teams identified consecutive eligible index TB patients at government clinics and hospitals within boundaries of study sites. We included TB patients of any age, but required those ≥7 years to have laboratory-confirmed pulmonary TB, whereas those <7 years could have physician-diagnosed TB of any organ, with or without laboratory confirmation. We additionally included TB patients who died within eight weeks of TB diagnosis. We excluded institutionalized TB patients and withdrew participants whose households we could not locate or from where no household member could be recruited. A list of household contacts was obtained at enrollment.

### Randomisation, allocation and blinding

Index cases and their households were block-randomised to either intervention or standard of care (SOC) in a 1:1 ratio, stratified by district. Investigator blinding was maintained until after the final participant household follow-up was completed.

### Procedures

In the intervention group, research fieldworkers visited households within 14 days of index TB patient enrollment (maximum three attempts), obtaining written individual or parental consent for adults and children under 18 years, respectively, with assent from older children. A questionnaire was administered to each household member (S2 Questionnaires), and sputum specimens obtained where possible (but not required from children <5 years) and were tested using Xpert and Mycobacterial Growth Indicator Tube (MGIT) culture. Household contacts received TST (from a variety of sources due to global shortages), administered and read within 72 hours [23]. Study nurses dispensed the first month of TPT (six months of daily isoniazid) to: (i) HIV-positive participants who tested negative for TB, (ii) HIV-negative participants with positive TST (≥10mm), and (iii) children under five years. Subsequent TPT was obtained from local clinics.

For household members without a confirmed HIV diagnosis, rapid point-of-care HIV testing was offered to participants ≥18 months, and PCR on dried blood spot for children <18 months whose maternal HIV status was unknown or positive. Participants with HIV had a CD4 count and were referred to their nearest clinic for assessment and initiation of ART.

Intervention households were visited approximately three months after enrollment to support treatment linkage.

In the SOC arm, index TB patients (or their representative, if deceased or a child) were given referral letters for every household member by the recruiting team at the health facility, recommending that each household contact should take their letter to their local clinic and be screened for TB and HIV.

### Outcomes

At 15-months after randomisation, study teams visited all households, updated the household membership list and recorded episodes of incident TB and death. We investigated household members for HIV (if untested) and TB (if symptomatic). All children ≤14 years old had TST placed, read at 48-72 hours.

The primary outcome was time to TB or death, measured among all household members included in the household census at baseline, from one month after randomisation through the final 15-month ascertainment visit. Primary analysis included all incident TB diagnoses, irrespective of diagnostic method; sensitivity analyses included only bacteriologically-confirmed incident cases of TB.

Secondary outcomes were: prevalence of TB infection (TST induration **≥**10mm) at month-15 among household children ≤14 years; time to initiation of TB treatment; and prevalence of undiagnosed or untreated HIV at month-15. Primary analyses for all outcomes were restricted to household contacts resident at baseline enumeration; supplementary analyses included all household contacts regardless of baseline residency. In protocol-specified subgroup analysis, we compared outcomes by trial site, and TST positivity by household contact age (<5 years, ≥5 years).

Ethical approval was granted by the University of Witwatersrand Human Research Ethics Committee (Medical), and the London School of Hygiene and Tropical Medicine.

### Statistical Methods

Assuming mean household size 5.5, 1,200 index cases per site (total 2,400), provided 80% power to detect a 30% overall difference in the primary outcome between groups with alpha 0.05 and intracluster correlation coefficient 0.3. All statistical analysis used Stata v16 (StataCorp, College Station, US). Analysis was done on an intention-to-treat basis.

This study is reported following CONSORT guidelines for cluster randomised trials (S3 Checklist). We summarised baseline index and household characteristics by trial arm. For the primary outcome, follow-up time began one month after randomisation (to avoid counting prevalent TB cases) and ended at the month-15 visit, or the date of TB or death. Cox proportional hazards regression with robust standard errors was used to assess the impact of the intervention on the primary outcome, with a time-by-treatment interaction term fitted to assess the proportionality assumption. Logistic regression with generalised estimating equations was used to assess the impact of the intervention on binary outcomes. Interaction terms were fitted to assess effect modification in planned subgroup analyses.

## Results

Between December 2016 and March 2019, we approached 2,393 potentially eligible TB index patients, of whom 2,062 were randomised (Figure 1). Characteristics of index cases were balanced between arms (Table 1). There were 4,459 household contacts identified in the intervention arm and 4,192 in the SOC arm (Table 2).

**Figure 1:**
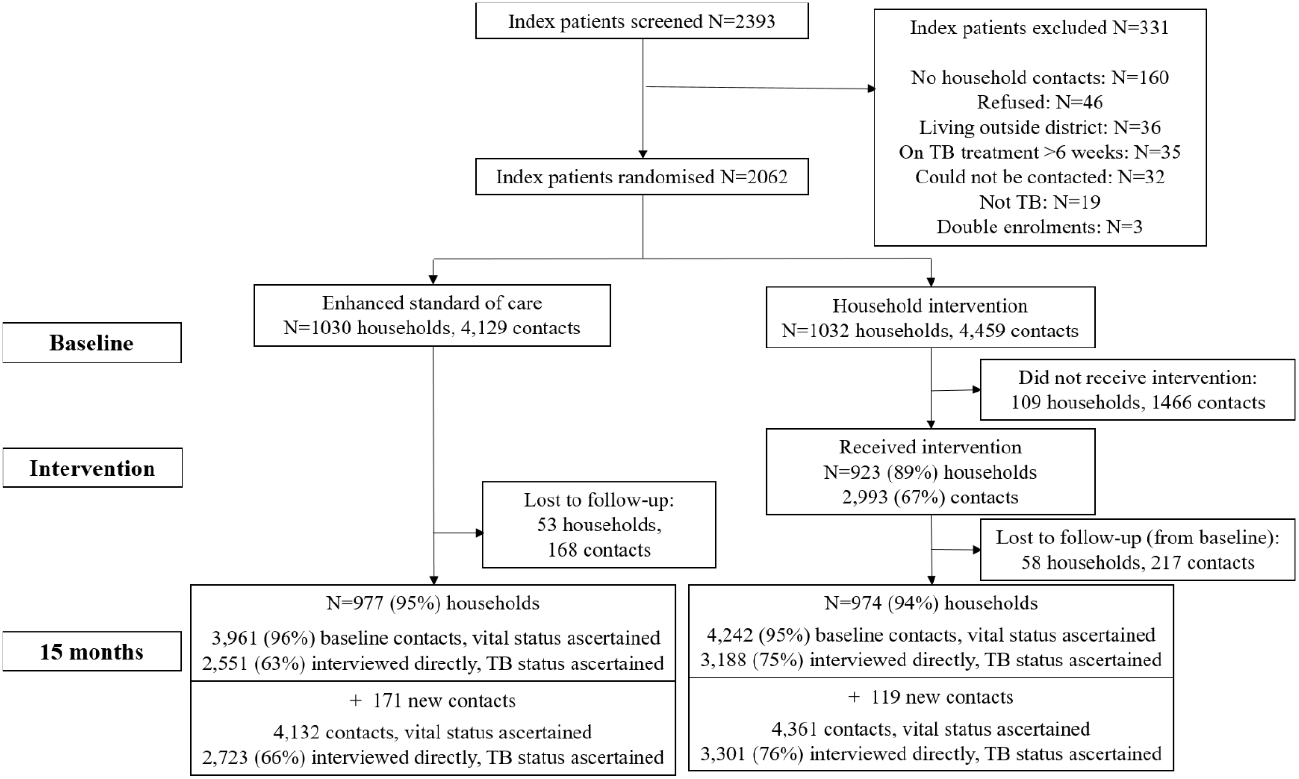
Consort Diagram of Cluster Randomised Trial.

**Table 1:** Characteristics of index patients randomised, by site and trial arm.

| Characteristic | Study site |  | Trial arm |  |
| --- | --- | --- | --- | --- |
|  | Mangaung<br>(n=1074) | Capricorn (n=988) | Standard of care<br>(n=1030) | Intervention<br>(n=1032) |
| Age (years), median (IQR) | 37 (28, 48) | 38 (29, 48) | 37 (28, 48) | 37 (28, 48) |
| Age group (years) |  |  |  |  |
| <8 | 19 (2%) | 26 (3%) | 25 (2%) | 20 (2%) |
| 8-14 | 9 (1%) | 11 (1%) | 14 (1%) | 6 (1%) |
| 15-19 | 55 (5%) | 36 (4%) | 48 (5%) | 43 (4%) |
| 20-29 | 249 (23%) | 179 (18%) | 211 (20%) | 217 (21%) |
| 30-39 | 323 (30%) | 337 (34%) | 325 (32%) | 335 (32%) |
| 40-49 | 191 (18%) | 209 (21%) | 200 (19%) | 200 (19%) |
| 50+ | 228 (21%) | 190 (19%) | 207 (20%) | 211 (20%) |
| Sex, male | 666 (62%) | 573 (58%) | 618 (60%) | 621 (60%) |
| Employment |  |  |  |  |
| Currently employed | 254 (24%) | 113 (11%) | 179 (17%) | 188 (18%) |
| Not employed | 715 (67%) | 679 (69%) | 712 (69%) | 682 (66%) |
| Student/child | 81 (8%) | 93 (9%) | 86 (8%) | 88 (9%) |
| Other | 24 (2%) | 103 (10%) | 53 (5%) | 74 (7%) |
| Income type |  |  |  |  |
| Salary | 216 (20%) | 136 (14%) | 162 (16%) | 190 (18%) |
| Wage | 70 (7%) | 41 (4%) | 50 (5%) | 61 (6%) |
| Grant | 235 (22%) | 215 (22%) | 226 (22%) | 224 (22%) |
| No income | 553 (51%) | 596 (60%) | 592 (57%) | 557 (54%) |
| Sputum Xpert |  |  |  |  |
| Positive | 1053 (98%) | 849 (86%) | 952 (92%) | 950 (92%) |
| Negative | 3 (0.3%) | 14 (1%) | 6 (1%) | 11 (1%) |
| Not done | 18 (2%) | 125 (13%) | 72 (7%) | 71 (7%) |
| Sputum smear |  |  |  |  |
| Positive | 11 (1%) | 295 (30%) | 148 (14%) | 158 (15%) |
| Negative | 8 (1%) | 52 (5%) | 32 (3%) | 28 (3%) |
| Not done | 1055 (98%) | 641 (65%) | 850 (83%) | 846 (82%) |
| Sputum culture |  |  |  |  |
| Positive | 0 (0%) | 59 (6%) | 22 (2%) | 37 (4%) |
| Negative | 4 (0.4%) | 6 (1%) | 7 (1%) | 3 (0.3%) |
| Not done | 1070 (99.6%) | 923 (93%) | 1001 (97%) | 992 (96%) |
| TB diagnosis |  |  |  |  |
| Microbiologically confirmed | 1054 (98%) | 939 (95%) | 990 (96%) | 1003 (97%) |
| Not microbiologically confirmed | 20 (2%) | 49 (5%) | 40 (4%) | 29 (3%) |
| HIV status (self-reported) |  |  |  |  |
| Positive | 590 (55%) | 521 (53%) | 555 (54%) | 556 (54%) |
| Negative | 421 (39%) | 442 (45%) | 434 (42%) | 429 (42%) |
| Unknown | 63 (6%) | 25 (3%) | 41 (4%) | 47 (5%) |
| On ART |  |  |  |  |
| Yes | 345 (58%) | 381 (73%) | 364 (66%) | 362 (65%) |
| No | 245 (42%) | 140 (27%) | 191 (34%) | 194 (35%) |
| BMI (kg/m<sup>2</sup>), mean (SD) | 19 (5) | 20 (5) | 19 (5) | 20 (5) |
| Karnofsky score, median (IQR) | 80 (70, 90) | 80 (70, 95) | 80 (70, 90) | 80 (70, 90) |
| Smoking (among 15+ years old) |  |  |  |  |
| Current | 254 (24%) | 113 (12%) | 176 (18%) | 191 (19%) |
| Previous | 268 (26%) | 216 (23%) | 238 (24%) | 246 (24%) |
| Never | 524 (50%) | 622 (65%) | 577 (58%) | 569 (57%) |
| Alcohol use (among 15+ years old) | 340 (33%) | 223 (24%) | 263 (27%) | 300 (30%) |

IQR: Interquartile range, TB: tuberculosis, HIV: human immunodeficiency virus, ART: antiretroviral therapy, BMI: body mass index.
Karnofsky score is a measure of participants' ability to undertake activities of daily living, and ranges from 0 (dead), to 100 (normal; no complaints; no evidence of disease).

**Table 2:**
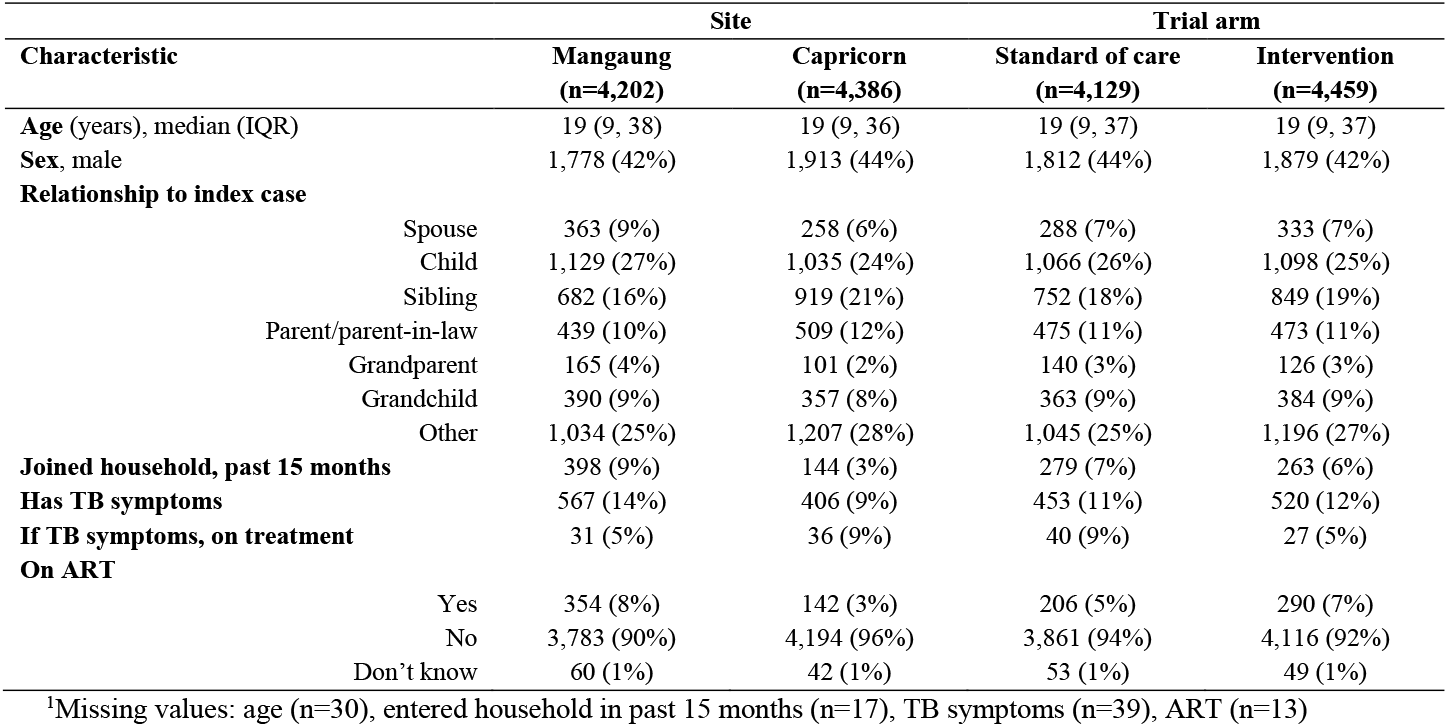
Characteristics of baseline household contacts of index patients, by site and trial arm.

| Characteristic | Site |  | Trial arm |  |
| --- | --- | --- | --- | --- |
|  | Mangaung<br>(n=4,202) | Capricorn<br>(n=4,386) | Standard of care<br>(n=4,129) | Intervention<br>(n=4,459) |
| Age (years), median (IQR) | 19 (9, 38) | 19 (9, 36) | 19 (9, 37) | 19 (9, 37) |
| Sex, male | 1,778 (42%) | 1,913 (44%) | 1,812 (44%) | 1,879 (42%) |
| Relationship to index case |  |  |  |  |
| Spouse | 363 (9%) | 258 (6%) | 288 (7%) | 333 (7%) |
| Child | 1,129 (27%) | 1,035 (24%) | 1,066 (26%) | 1,098 (25%) |
| Sibling | 682 (16%) | 919 (21%) | 752 (18%) | 849 (19%) |
| Parent/parent-in-law | 439 (10%) | 509 (12%) | 475 (11%) | 473 (11%) |
| Grandparent | 165 (4%) | 101 (2%) | 140 (3%) | 126 (3%) |
| Grandchild | 390 (9%) | 357 (8%) | 363 (9%) | 384 (9%) |
| Other | 1,034 (25%) | 1,207 (28%) | 1,045 (25%) | 1,196 (27%) |
| Joined household, past 15 months | 398 (9%) | 144 (3%) | 279 (7%) | 263 (6%) |
| Has TB symptoms | 567 (14%) | 406 (9%) | 453 (11%) | 520 (12%) |
| If TB symptoms, on treatment | 31 (5%) | 36 (9%) | 40 (9%) | 27 (5%) |
| On ART |  |  |  |  |
| Yes | 354 (8%) | 142 (3%) | 206 (5%) | 290 (7%) |
| No | 3,783 (90%) | 4,194 (96%) | 3,861 (94%) | 4,116 (92%) |
| Don't know | 60 (1%) | 42 (1%) | 53 (1%) | 49 (1%) |

<sup>1</sup>Missing values: age (n=30), entered household in past 15 months (n=17), TB symptoms (n=39), ART (n=13)

A total of 974 (94%) and 977 (95%) of households randomised to the intervention and SOC arms, respectively, took part in final outcome assessments, with vital status information available for 4,242 (95%) and 3,961 (96%) household contacts captured in baseline censuses. In households with outcome assessments, an additional 119 (intervention) and 171 (SOC) individuals moved into the household after the baseline census. Supplementary analysis was therefore based on a total of 4,361 household contacts in the intervention arm and 4,132 in the SOC arms, of whom 3,301 (76%), and 2,723 (66%) respectively were interviewed directly at the 15 month visit and had both TB and HIV outcomes ascertained.

Of households randomised to the intervention arm, 923/1032 (89%) received the intervention, 516 (96%) in Mangaung and 407 (82%) in Capricorn. In the 923 households where the intervention was provided, a total of 2,993 household contacts consented, and then received the intervention (median 3 per household, IQR: 2-4). The prevalence of microbiologically-confirmed TB among intervention arm household contacts who provided sputum was 3% (69/2166). Overall, 13% (368/2752) had positive TST results; 763 initiated TPT.

The primary outcome, incident TB or death among household contacts present at baseline enumeration, was similar between the household intervention (93/3230, 3%) and SOC arms (80/2600, 3%, hazard ratio [HR]: 0.90, 95% CI: 0.66-1.24, p=0.54, Table 3). There was some evidence that the proportional hazards assumption was violated, with cumulative hazard curves crossing at approximately the target follow-up time of 15 months (Figure 2). Allowing the hazard ratio to vary, there was no effect of the intervention either in the first 15 months of follow-up or had their outcome visit beyond 15 months (HR 1.01, 95% CI: 0.72-1.42, p=0.95 and 0.45, 95% CI: 0.19-1.05, p=0.07, respectively).

**Table 3:** Effect of intervention versus standard of care on trial outcomes, among household contacts who were present at baseline list of household contacts.

| Outcome | Standard of care | Household Intervention |  |
| --- | --- | --- | --- |
| **Primary outcome** |  |  | **Hazard ratio (95% CI)** |
| Contacts diagnosed with TB | 31/2551 (1.2%) | 51/3188 (1.6%) | 1.33 (0.83, 2.16) |
| Contact deaths | 49/3961 (1.2%) | 42/4242 (1.0%) | 0.72 (0.47, 1.10) |
| TB or death | 80/2600 (3.1%) | 93/3230 (2.9%) | 0.90 (0.66, 1.24) |
| **Secondary outcomes** |  |  | **Odds ratio (95% CI)** |
| Prevalence of TST positivity ($\geq 10$mm) among children $\leq 14$ years | 15/800 (1.9%) | 38/845 (4.5%) | 2.25 (1.07, 4.72) |
| Prevalence of undiagnosed or untreated HIV infection | 32/2543 (1.3%) | 41/3185 (1.3%) | 1.02 (0.64, 1.64) |
| 444 Hazard ratios and odds ratios were calculated with the standard of care arm as the reference group; intracluster<br>445 correlation coefficients (ICC) were TB (0.04), death (0.004), TB or death (0.02), TST positivity (0.86),<br>446 undiagnosed/untreated HIV (0.08)<br>447 |  |  |  |

**Figure 2:**
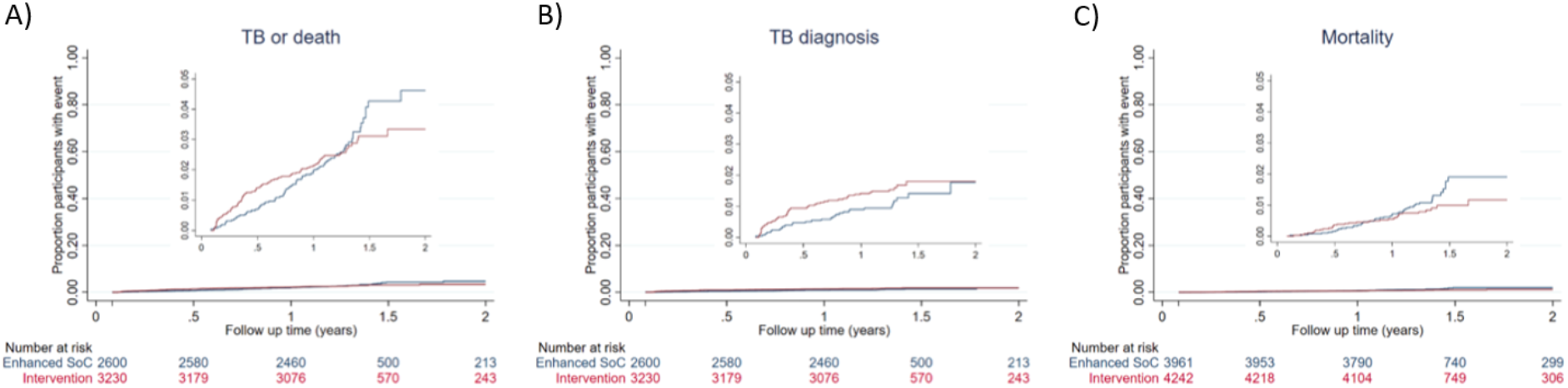
Cumulative hazard of incident TB or death among household contacts of TB patients by trial arm. A: Primary trial outcome of incident TB diagnosis or death between months 1 and 15. Inset: y-axis truncated to show range of data; B) Incident TB diagnosis between months 1 and 15. Inset: y-axis truncated to show range of data; C) Mortality between months 1 and 15. Inset: y-axis truncated to show range of data.

Protocol-specified sensitivity analyses for the primary outcome, including those who had entered the household after the baseline census, and based on only bacteriologically-confirmed cases of TB, showed similar results (S4 Table). In protocol-specified subgroup analysis (S5 Table), there was no difference in the composite primary outcome of incident TB or death at either trial site (Mangaung, HR 1.26, 95% CI: 0.81-1.97; Capricorn, HR: 0.63, 95% CI: 0.40-1.00), but death was lower among household contacts in Capricorn in the intervention arm compared to the SOC arm (HR: 0.56, 95% CI: 0.32-0.97).

In the intervention arm, a total of 51/3188 (2%) individuals without TB in the baseline census had an episode of incident TB, compared to 31/2551 (1%) in the SOC arm. Approximately half of the individuals diagnosed with incident TB had biological confirmation (45% in the intervention arm, 52% in the SOC arm) and 3 (4%) incident TB diagnoses were diagnosed at the final outcome visit (2 in intervention arm, 1 in SOC arm). Incidence of TB was 1.24 and 0.92 per 100 person-years among intervention and SOC household contacts, respectively (HR: 1.33, 95% CI: 0.83-2.16, p=0.24).

Overall, 69 participants were diagnosed with TB through trial screening: 24 by Xpert alone, 22 by culture alone, 9 by smear alone and 14 by more than one test. Of 69 diagnosed with TB, by the three month visit 37 (54%) were successfully referred for, and initiated TB treatment. The median time between date of sample being taken and patient initiating TB treatment was 3 days (IQR: 0-13), and was somewhat higher in the intervention arm compared to the control arm (median: 4 days, IQR: 0-28 versus median 3 days, IQR: 0-4, p=0.04).

A total of 91 deaths were ascertained among household contacts in the baseline census population, 42/4,242 (1%) in the intervention group and 49/3,961 (1%) in the SOC group. Incidence of mortality was 0.68 and 0.94 per 100 person-years among intervention arm and SOC household contacts, respectively (HR: 0.72, 95% CI: 0.47-1.10, p=0.13).

New HIV diagnoses at month-15 occurred with similar frequency in both arms (intervention arm 11/3185, 0.4% versus standard arm 22/2543, 0.9%). At the month-15 visit 31 HIV-positive participants in the intervention arm and 11 participants in the SoC arm were not taking ART. Thus, the prevalence of undiagnosed or untreated HIV at the final visit was comparable between trial arms (both 1%, OR 1.02, 95% CI: 0.64-1.64, p=0.92).

A total of 2,271 household contacts seen in person were aged 14 years or younger at the time of the final visit. Of these, 1,664 (73%) had TST placed (857 in the intervention arm, 807 in the SOC arm), and 1,645 had the result recorded with 38 (5%) in the intervention arm and 15 (2%) in the SOC arm testing positive (odds ratio: 2.25, 95% CI: 1.07-4.72, p=0.03). In protocol-specified subgroup analysis (S6 Table), TST positivity was higher in the intervention arm compared to the SoC arm among participants aged ≥5 years, but not among participants aged <5 years. At 15-months, of those assessed, 73% of household contacts in the intervention arm reported having taken 6-months of TPT, with 14% taking 1 month or less.

## Discussion

In this trial, a strategy of household contact tracing and intensive screening for TB and HIV did not affect the composite outcome of incident TB or death and was equivalent to providing clinic referral letters to TB index patients. Moreover, we found no difference in prevalence of undiagnosed HIV between arms. A greater proportion of children in the intensive arm had latent TB infection by TST testing compared to the SOC arm. Overall household tracing and intensive investigation of household contacts for TB and HIV does not offer sustained benefit beyond the initial screening episode, despite relatively high rates of detection of prevelant undiagnosed HIV and TB.

Our trial differs from previous randomised trials of household contact tracing for TB. In the ZAMSTAR Study, conducted in South Africa and Zambia, household contacts received TB symptom screening, followed by sputum smear microscopy if symptomatic, and HIV testing and TPT [11]. There was weak evidence that household interventions reduced TB prevalence and childhood TB transmission. In Uganda, intervention households received HIV testing and linkage to care, and TB symptom screen followed by smear microscopy or Xpert, and with SMS-supported linkage to care [12]. Completion of TB investigation and yield of TB diagnosis were not different between trial arms. In contrast, in a cluster-randomised trial in Vietnam household contacts of TB patients were invited for clinic-based screening, comprising symptom assessment and chest radiography, followed by smear microscopy and culture if positive [10]. In that trial, there was a 2.5-fold increase in TB treatment registrations in the household contact screening arm compared to the passive case detection arm. In a cluster-randomised trial in Rio de Janeiro, Brazil, supplementing the WHO DOTS strategy with more intensive interventions for household contacts (symptom screening, chest X-ray, TST) resulted in reductions in TB case notification rates [13].

Whereas these previous trials limited microbiological investigation to household contacts with a positive symptom screen or abnormal chest x-ray, we evaluated the provision of microbiological testing on household contacts who could provide a sample, prompted by evidence that a substantial fraction of community members with microbiologically-confirmed TB have minimal or no symptoms [14,15]. We successfully obtained sputum samples from 92% of intervention arm adults ≥15 years. Compared to previous studies, which mostly used smear microscopy, we used the more sensitive Xpert platform with MGIT culture for sputum testing, and made a home visit to intervention arm households to prompt linkage to care [24,25]. Despite this, a high percentage of household contacts with microbiologically-confirmed TB did not initiate TB treatment, emphasising that new approaches to improving linkage are needed. Our HIV testing strategy intended to identify the anticipated small proportion of people with undiagnosed HIV, and link promptly to ART initiation and TPT, thereby reducing the risk of incident TB disease and death.

Despite achieving high intervention coverage and follow-up, we saw no difference in TB-free survival between arms. There are a number of possible explanations for this. We anticipated that a letter prompting clinic-based screening for household contacts in the SOC arm would be insufficient to achieve satisfactory levels of screening completion and treatment linkage [26]. However, the percentage of household contacts initiating TB treatment was only slightly higher in the intervention arm (1.6%) compared to the SOC arm (1.2%), perhaps reflecting high motivation for TB screening among household contacts receiving letters. It is also possible that in intervention households, TB transmission had already occurred at the time of the intervention. Finally, as the majority of TB transmission is thought to occur outside households,[27] high forces of infection within South African communities may overcome the benefits of interventions targetting households. Indeed, in Capricorn, a relatively low annual TB incidence area (for South Africa), there was a suggestion that the intervention was effective in reducing mortality.

Our findings suggest that household contact tracing with home visits and intensive screening for TB and HIV is unlikely to be considered for implementation by National TB Programmes in low-resourced, high TB burden settings. Although household contact tracing of index TB patients is widely recommended, implementation is often poor due to the substantial resource requirements. Cost studies will be reported separately, but we anticipate resource implications of household visits to be substantial. Conversely, the strategy of providing referral letters for household contacts to take to their local clinics to prompt facility attendance for TB/HIV screening and care is affordable and implementable at scale, but requires further implementation research and evaluation.

We found that, in the intervention arm, prevalence of latent TB (defined by TST) was 13%, comparable to previous household contact tracing studies from the region [26], and with strong age- and site-specific dependency [28]. At 15-months, TST positivity in children was higher in the intervention arm than the SOC arm. One possible explanation is differential rates of acceptance of TST between the intervention and SOC arms by site: in the intervention arm in Mangaung, completion of TST was 77% compared to 55% in Capricorn; in the SOC arm, completion was 84% at both sites. We did not record data on reasons for refusal of the TST but it may be that those with a strong response previously were reluctant to be retested: in Capricorn, children who had baseline TST done were less likely to have the month-15 TST done (baseline TST done: 214/485, 44% vs. baseline TST not done: 167/205, 81%).

The study had several limitations. The planned sample size was reduced due to budget constraints; follow-up time in 14% of households was reduced in anticipation of South African COVID-19 pandemic-related lockdown (S7 Checklist) [29]. The study was at a time when South African preventive treatment guidelines were in flux, initially requiring a positive TST to initiate TPT and with restrictions on people who could receive TPT. This likely reduced the proportion of HIV-negative individuals older than five years who continued TPT beyond the initial study-dispensed month. Not all households allocated to the intervention arm received interventions, mainly due to difficulties locating households.

In conclusion, an intensive household contact tracing and TB/HIV screening intervention did not reduce incident TB or death when compared to a referral letter intervention. TB programme managers and policy makers should carefully consider benefits and costs before implementing similar household contact tracing and TB screening interventions. The provision of referral letters to index patients at the time of their TB diagnosis should be the preferred strategy to link household contacts to care in low-resourced high-TB burden settings.

## Supporting information

S1 Protocol

S2 Questionnaires

S3 Checklist

S7 Checklist

## Data Availability

Data will be made available online on manuscript publication.

## Acknowledgements

We acknowledge the expert oversight and advice from the members of the Data and Safety Monitoring Board (Derek Sloan, Yunus Moosa, and Locadiah Kuwanda).

## Supplemental material

S1 Protocol: Study protocol (version 5)

S2 Questionnaires: Study questionnaires

S3 Checklist: CONSORT extension for cluster randomised trials checklist

S4 Table: Effect of intervention versus standard of care on primary trial outcome, sensitivity analyses

S5 Table: Subgroup analyses of primary and secondary outcomes by study site

S6 Table: Subgroup analyses of prevalence of TST positivity, by age of child

S7 Checklist: CONSERVE-CONSORT Checklist

