## Supplementary material for "Household contact tracing with intensified tuberculosis and HIV screening in South Africa: a cluster randomised trial": S1 Protocol

**A household cluster randomised trial of active case finding for HIV and TB, preventive treatment against TB, and ART initiation to prevent TB disease and transmission.**
(The HomeACF Study)

Protocol
Version 5.0	17 August 2018

**Principal investigators:**
Dr Neil A Martinson
CEO, Perinatal HIV Research Unit
A Division of the Wits Health Consortium (Pty) Ltd
PO Box 114, Diepkloof, 1864
South Africa


Dr Peter MacPherson
Academic Clinical Lecturer
Department of Public Health and Policy
The Farr Institute@HeRC
Waterhouse Building (2^nd^ Floor, Block F)
1-5 Brownlow Street
Liverpool L69 3GL, United Kingdom


### Study Team

| **Principal Investigators** |  |
| --- | --- |
| Dr Neil A Martinson  | Perinatal HIV Research Unit  Diepkloof, South Africa |
| Dr Peter MacPherson MBChB PhD  | Department of Public Health and Policy  University of Liverpool, UK  Honorary Clinical Lecturer  Department of Clinical Sciences  Liverpool School of Tropical Medicine, UK |
| **Co-investigators** |  |
| Prof Ebrahim Variava  | Klerksdorp Tshepong Hospital Complex and University of the Witwatersrand |
| Dr Sanjay Lala  | Department of Paediatrics Chris Hani Baragwanath Academic Hospital and University of the Witwatersrand |
| Prof Andrew Ratsela  | Department of Medicine, Polokwane Hospital and University of Limpopo |
| Ms Katlego Mothlaoleng  | Project Director PHRU Matlosana |
| Dr L Lebina  | Project Director PHRU |
| **Statistician** |  |
| Dr Emily Webb  | London School of Hygiene and Tropical Medicine  London, UK |
| **Health Economist** |  |
| Dr Anthony Kinghorn  | Perinatal HIV Research Unit  Diepkloof, South Africa |
| **Epidemiology Consultant**  Dr Jonathan Golub  | Johns Hopkins University School Medicine, Baltimore |
| (Not involved in management of the study) |  |

### Executive Summary

Tuberculosis transmission occurs predominantly indoors, often at home. We propose this trial to assess whether an improved and intensified household intervention for household contacts of TB cases prevents TB infection and disease in two different TB burden settings.

The recent ZAMSTAR study suggests that household contact tracing strategies for TB prevention may be more effective in preventing future TB infections and cases than untargeted community-based case finding approaches, which are also resource intensive and difficult to sustain. Since ZAMSTAR was completed there have been major strides in the diagnosis and prevention of TB – particularly in high HIV prevalence settings. Additionally, we have previously shown that household contacts of TB cases had substantially higher rates of undiagnosed TB compared to randomly selected households without an index TB case, and that a substantial proportion of culture-positive households contacts are asymptomatic. Moreover, we and others, have shown that using children as sentinel cases to direct active case finding also has a high yield of undiagnosed TB in household contacts. Finally, it appears that in high HIV prevalence areas, rates of TB in household contacts are high irrespective of whether contacts live in a TB “hot spot” or a far lower annual TB incidence setting.

We therefore propose a cluster randomised trial in three health districts in South Africa to investigate the effect of an intensified active case finding strategy with immediate initiation and of treatment and linkages to care for household contacts of index cases of TB diagnosed at two sites with different TB burdens. The three selected health districts are Matlosana, Mangaung and Capricorn, with annual TB incidence of 937/100,000; 686/100,000and 371/100,00 respectively. One thousand two hundred hospital- and/or clinic-diagnosed index TB cases of all ages will be recruited at each site. Their households will be randomly allocated to receive either an enhanced standard of care contact tracing strategy using referral letters with follow up phone calls, or an intensified household contact tracing and treatment support (IHCS) intervention comprising of:

1. Sputum Xpert and TB liquid culture for all household contacts capable of providing a sputum sample, irrespective of TB symptoms, with supported initiation and linkage to TB treatment in those with TB disease;
2. HIV testing with prompt initiation of both antiretrovirals and TB preventive treatment for HIV-infected contacts according to current South African guidelines,
3. Preventive therapy for tuberculin skin test-positive, HIV-negative **and** HIV-infected household contacts without active TB disease;
4. A three-month supportive follow-up visit to confirm treatment receipt, and assist those who have either not accessed care and treatment; or who have experienced problems with their treatment; or who have developed symptoms of TB in the interim.

**Objectives**:

Primary trial outcomes are:

1. Comparison between trial groups of TB FREE SURVIVAL from two months after randomisation – to the final ascertainment visit among all household members identified at baseline – a total of 13 months. We will measure this in two ways; firstly, including all cases of where TB was diagnosed or multi drug TB treatment started, irrespective of the diagnostic method used and secondly, including only those with a hard copy of a laboratory confirmation of TB.

*Secondary trial outcomes*

1. Comparison between trial groups of the proportion of TB cases with diagnostic delay (defined as number of days between onset of symptoms and diagnosis of anti-tuberculosis treatment) among household members identified at baseline and diagnosed with TB between baseline and month 15
2. Comparison between trial groups of the prevalence of TB infection measured by tuberculin skin test reactivity >10mm at month 15 among household children identified at baseline and aged under 14 years of age.
3. Comparison between trial groups of the prevalence (reported as a percentage of those tested) of previously undiagnosed and untreated HIV infection among household members identified at baseline and again at month 15.
4. Comparison between trial groups of the cumulative incidence of all-cause mortality among all household members identified at baseline between baseline and month 15.
5. Comparison between trial groups of the cumulative incidence of TB- and HIV-related non-traumatic mortality among all household members identified at baseline between baseline and month 15.
6. Estimation of the cost-effectiveness of the intensified housed TB and HIV case-finding and prevention activities compared to the enhanced standard of care from a societal perspective. This will include comparisons of household costs and of quality of life of household members in the IHF and enhanced standard of care arms.

*Exploratory outcomes*

1. Cohort analysis of the incidence of bacteriologically-confirmed, and all-forms of tuberculosis among intervention group household members receiving and not receiving isoniazid preventive therapy, stratified by HIV serostatus.
2. Cohort analysis of the proportion of household members in the intervention group with previously undiagnosed HIV infection identified at baseline who successfully initiate antiretroviral therapy by month 15.
3. Estimation of the fraction of TB transmission to household members attributable to household exposure to TB index cases by analysis of whole genome sequences of sputum TB isolates (*Subject to further funding being secured)*
4. To quantify catastrophic costs linked to a new TB and/or HIV diagnosis or death due to TB and/or HIV of a household member (estimated at baseline prior to the intervention being implemented).

Developmental Outcome: This project will support and train the newly established Department of Internal Medicine at the University of Limpopo Medical School in Polokwane to conduct clinical research, as we will include both the Capricorn Health District in Limpopo Province and the Matlosana sub-district in North West Province. In addition, we will ensure that at least three South African masters students and at least one PhD student use data from this study for their research projects.

Background

*TB Epidemiology in high HIV prevalence settings*

Tuberculosis remains a public health priority, and is now the leading cause of death from an infectious disease worldwide. In sub-Saharan Africa, TB incidence rates initially skyrocketed, driven principally by extremely high prevalence rates of HIV infection. South Africa is the country with the largest number of people living with HIV worldwide, and has experienced extremely high rates of TB incidence, prevalence and mortality.

The new End-TB Strategy calls for bold action to eliminate tuberculosis as a public health issue by 2035. Central to the End-TB strategy is a recognition that intensified action, driven by innovate research, is required to find and treat all cases of tuberculosis, and to identify high-impact strategies to limit transmission to vulnerable individuals. Moreover, high out of pocket costs of care-seeking and treatment have proved to be catastrophic for many people living in poverty. Interventions that can effectively make TB treatment equitable, affordable and accessible are required.

*Intensified case-finding approaches for tuberculosis need to be evaulated*

Contact tracing of index TB cases (sometimes known as “sentinel” or “source” cases) has been advocated as a key part of TB control for many years (Grzybowski, 1975; World Health Organization, 2012; Rieder, 2003) because it facilitates early identification of symptomatic and infectious linked cases, and allows individuals with TB infection to receive preventive treatment. Although, World Health Organization (WHO) and National Guidelines in South Africa recommend household contact tracing, the 2012 WHO guidelines have not been prioritized or widely implemented in South Africa (or indeed many other high TB burden countries) because of the current very low quality of available evidence for effectiveness, and the requirement to reallocate resources in settings that are struggling to identify and treat TB patients presenting with symptoms (Claassens, 2013).

Recently, the groundbreaking ZAMSTAR community randomized trial in Zambia and South Africa (Ayles, 2013) has shown that a household contact tracing intervention was superior to an untargeted community TB case finding intervention, albeit with borderline significant benefit on two primary community-wide outcome measures: reduction of new TB infections in young children; and reduction of prevalence of TB in the communities which received contact tracing. The ZAMSTAR household contact tracing intervention comprised of: Identification of target households through adult index TB cases; symptom screening of household contacts to identify individuals with presumptive TB for further investigation; screening of symptomatic individuals using sputum smear microscopy. To date, the ZAMSTAR study team has not reported on process and outcome indicators of the household intervention, such as: newly diagnosed with TB and/or HIV who accessed the care and prevention services; or the number of times the entire household was offered TB symptom screening; or the proportion of patients who should have received TB preventive treatment, and/or ART, who actually received it. Additionally, the symptom-based, sputum smear screening strategy used in ZAMSTAR to identify and investigate TB suspects likely missed household contacts with prevalent infectious TB, thereby limiting the effectiveness household intervention.

Our studies in households of adult TB cases in two different South African settings where HIV prevalence is high, suggests that a substantial proportion of household contacts who have sputum cultures positive for *M tuberculosis* do not report symptoms and, under the ZAMSTAR strategy, would ***not*** be identified by symptom screening (Shapiro, 2013). Moreover, unpublished data from the Vhembe health district in Limpopo (a South African TB “cold spot”) showed that household contact tracing identified similar rates of undiagnosed TB in household contacts as in the TB hotspot of Matlosana, and also, many of them were asymptomatic. (Little K, et al; submitted).

Mathematical modelling work emphasizes that failure to detect asymptomatic or subclinical TB limits the impact on community TB transmission and that earlier identification and prompt treatment results in improved population-level TB control (Dowdy, 2013). Moreover, sputum smear has insufficient sensitivity when used in contact tracing, where a large proportion of contacts are HIV infected, and active TB is identified earlier and is likely to be paucibacillary.

In the intervening period since the final ZAMSTAR prevalence survey in 2010, there have been considerable advances in TB diagnosis, and prevention. Routine TB diagnostics have improved markedly with the widespread scale-up of the Xpert MTB/Rif molecular TB diagnostic platform (Boehme CC, 2010) (and more recently with a newer, apparently more sensitive version the Xpert Ultra not yet available for use). Xpert MTB/Rif reduces diagnostic delay and results in earlier TB treatment initiation (Theron G, 2014); TB preventive treatment guidelines for HIV-infected individuals have evolved to recommend long-term, more effective regimens (Samandari 2011, Martinson 2011), that when co-administered with antiretroviral therapy (ART), are safe and reduce mortality and TB disease (TEMPRANO ANRS 12136 Study Group 2015, Rangaka 2014). The CD4 threshold for initiating ART has been raised to 500 cells/mm3 and likely will be redundant when the test-and-treat strategy is implemented; and first line ART regimens have been simplified to single daily dosing with fixed-dose combination tablets. Moreover, children diagnosed with TB are increasingly being recognized as important sentinel cases for rapid identification of an infectious household TB case (Puryear,2013; Lala, 2014). Finally our data shows that HIV-seronegative household contacts in a high burden setting in the year after a household contact tracing intervention have annual TB incidence of 700/100,000 (van Schalkwyk, 2014) – suggesting preventive treatment against TB should be offered to HIV seronegative adults and older children in contact with a case, although this is not currently part of guidelines for preventive treatment in South Africa.

**We therefore hypothesise that a household intervention that actively includes these recent improvements will improve TB free survival** **and decrease the prevalence of new TB infections in children**.

### Overall aim

The overall aim of the study is to investigate the effect on household contacts of a TB case of an intensified household contact tracing and treatment support (IHCS) for household contacts of index adult and paediatric cases of TB diagnosed in clinics and hospitals in two sites with markedly different TB burdens.

### Objectives

**Primary objectives:**

1. To assess the impact of the intensified household case-finding strategy on:
   1.1. TB free survival among all household contacts
   1.2. Prevalence of TB infection in household contacts under 14 years of age.
2. To assess the cost effectiveness of IHCS compared to the enhanced standard of care.

**Secondary objectives:**

1. To assess the effectiveness of isoniazid preventive treatment on new cases of TB in HIV-seronegative individuals who are tuberculin skin test positive.
2. To compare the yield and costs of IHCS versus standard of care in identifying undiagnosed cases of TB and undiagnosed or untreated HIV
3. To compare rates of linkage to TB treatment between participants identified with TB through IHCS and standard of care
4. To explore perceptions and acceptability of doing home visits with household members

### Methods

#### Study design

Household cluster randomized trial of an intensified household contact tracing and treatment support (IHCS) intervention for contacts of both adult and paediatric TB cases.

#### Study setting

We will recruit index cases at two South African sites with large differences in annual TB incidence and HIV prevalence Mangaung Municipality (2011 population: 747 431) and the Capricorn Health District in Limpopo (2011 population: 1,293,000) (Stats SA Census 2011 Municipal Fact Sheet) – Figure 1). The antenatal HIV seroprevalence and the annual TB incidence for 2012 in Mangaung and Capricorn Health Districts were 36% and 937/100,000; 30.3% and 686/100,000 and 14.6% and 371/100,000 respectively (Dept of Health, 2013; Health Systems Trust 2013). These two settings will provide valuable information on the intervention when implemented in a TB “hot spot” compared to one with a lower annual TB burden.

Figure 1: Study sites

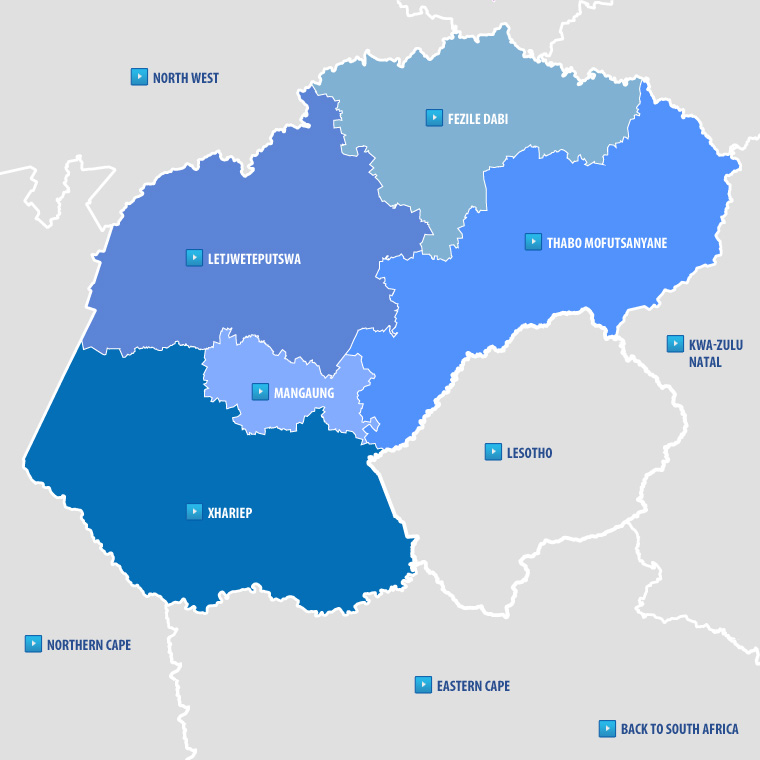

Figure 2. Matlosana municipality and Capricorn district sites

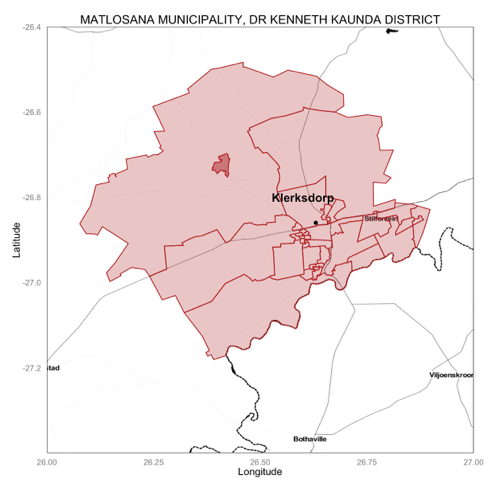

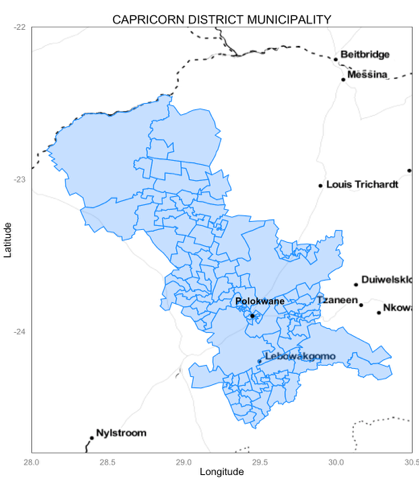

#### Index cases

Using routine reporting of TB cases in health facilities in the two study sites, and lists of specimens positive for TB generated by routine public sector laboratories in the two study sites, the recruiting teams will identify potentially eligible patients in clinics and hospitals. Once a potential index patient has been identified, if it appears they currently reside permanently within the boundaries of one of the study sites, they will be approached and invited to participate in the the study. Following the informed consent process, we will recruit consecutive index TB patients (those 7 years and older with pulmonary TB, and children under 7 years with TB of any organ) diagnosed with TB and currently residing in the catchment health facilities in each of the study sites. If older index cases (7 years and older) are extremely ill and are unable to provide their own assent or informed consent, we will obtain proxy consent from next-of-kin. Moreover, using sensitive approaches we will also attempt to obtain consent from next of kin of potential index cases who died soon after their diagnosis of TB.

**Inclusion Criteria for Index Cases**

1. Any patient (living or recently deceased) diagnosed with TB at any health facility in the study districts in the six weeks prior to recruitment.
   a. For index cases with TB older than 7 years of age, laboratory confirmation of **pulmonary TB disease** is required, evidenced by hard copy lab report of at least one of: a smear positive result for acid fast bacilli (graded as +, ++, or +++); an Xpert MTB/Rif test or other WHO recommended molecular assay positive for *M. tuberculosis*; a mycobacterial culture positively identified as *M tuberculosis*; or a histology/cytology report from a biopsy that includes lung tissue that is suggestive of TB disease. LF-LAM may be used to assist in the diagnosis of TB in HIV positive patients with signs and symptoms of TB (pulmonary and/or extrapulmonary) who have a CD4 cell count less than or equal to 100 cells/μL, or HIV positive patients who are seriously ill regardless of CD4 count or with unknown CD4 count. Evidence of a hard copy of recorded LF-LAM result is required.
a. For index cases 7 years and younger, the diagnosis of **TB of any organ** will be evidenced by – at a minimum- firstly: a recorded diagnosis of TB and its anatomical site in any clinic or hospital record; and secondly the prescription of multidrug TB treatment by a doctor for a child with signs and symptoms suggestive of TB. The presence of signs or symptoms suggestive of TB may be obtained either from the clinical record or from the caregiver.
2. Permanent resident in a household with at least one other person in either the Capricorn Health District or the Matlosana Municipality for a total of at least three months in the six months prior to recruitment.
3. Either the participant, the parent or next of kin of a participant declare they have no plans for relocation of their household to another municipality or health district during the course of follow up.

**Exclusion Criteria for Index Cases**

1. The research team is unable to obtain informed consent or assent for the following reasons:
   a. Participant is unwilling to provide informed consent or assent;
   b. The research team is unable to obtain informed consent from the parents of participants younger than 18 years;
   c. The research team is unable to obtain consent for participation from the close next-of-kin of adult patients who are too ill to provide their own consent, or have died.
2. Patients with TB who appeared to have developed or contracted TB or while either institutionalized or incarcerated. Similarly, no person incarcerated or institutionalized at the time of diagnosis will be included in the study.
3. If the study team is unable to locate the household of the index case, or if there is no other household member/s able to be recruited at the household, the participant will be withdrawn from the study.

#### Index case identification and consent procedures

A team of enrolled nurses will recruit index cases. They will be equipped and trained to use N95 or other appropriate respiratory protection and infection control measures and also in Good Clinical Practice. They will visit hospitals, clinics and TB laboratories in the catchment areas of the study on repetitive, frequent cycles, to identify potential index cases diagnosed in the past six weeks. Index cases and the parents of index cases younger than 18 will be approached to obtain consent for their own participation in the case of index cases; and in the case of parents of an index case, consent for participation of their child in research Using sensitive approaches, we will attempt to obtain consent from the close next-of-kin of patients with TB who died in hospital/ or at home. Deceased patients who were diagnosed with TB will only be enrolled into the study if date of death is no more than 6 weeks from date of enrolment. If a potential index case of any age is too unwell to comprehend the informed consent process, the research team will approach close next-of-kin of the patient with TB and ascertain if they would give consent for the participation of the index case and his/her household in the study. For this study close next-of-kin are: current spouse living together for past year in the same household, sibling older than 25 years of age, biological parent or biological grandparent – the latter if parents are not available or have passed away.

Adults sputum positive for acid fast bacilli, or with a cycle threshold (Ct) on Xpert MTB/Rif of ≤24, and those with drug resistant TB will take precedence for recruitment, in the event the study capacity to recruit eligible households is exceeded.

Enrolled nurses will provide prospective participants with verbal and written information about the study, emphasising the procedures involved in the study, especially that the study involves a study visit to the household. They will be informed that household members will be asked questions about the health, testing and treatment, and also about other issues such as costs of health care and illness. Following provision of information and answering any questions, individuals will be invited to provide informed consent to participate in the language of their choice, with consent recorded by signature on a study consent form. Parental consent to participate in research will be obtained for children <18 and assent will be obtained from children 7-14 years. Individuals who are illiterate, or unable to write, will be invited to provide a thumbprint confirmation of consent to participant, accompanied by a witness signature.

Potential participants, their parents, or their next of kin will be informed that non-participation will have no detrimental effect on the care they will continue to receive and that no incentives (financial or otherwise) will be provided to any study subject.

#### Procedures for all index case TB participants

After obtaining informed consent, enrolled nurses will administer a baseline index case demographic and clinical questionnaire (including presence and duration of symptoms, comorbid conditions, treatment received and laboratory confirmation of TB). Additionally, the study staff will obtain from each index case a handwritten household census comprising the first two letters of the first name and first two letters of the last name and ages and genders of all of their household members.

Index TB cases will continue to receive TB treatment and HIV testing and care through routine clinical services according to South African National Guidelines. Results of routine TB and HIV investigations completed by the health facility (sputum smear, sputum TB culture, Xpert MTB/Rif, HIV-test, CD4 count, HIV viral load), TB category (pulmonary, extra-pulmonary); and treatment initiation dates will be extracted from clinical records and recorded by study staff using data extraction forms).

#### Randomisation, allocation and blinding

The unit of randomization will be households of recruited index TB cases. The dwelling within which the household members live is defined as all rooms under a contiguous roofed area linked by doorways or windows through which air can pass. For this study, household members will be all individuals who shared airspace for lengthy periods – either slept overnight at least once, or shared at least two meals in the same household as the index case in the 14 days prior to the index case’s diagnosis of TB.

Randomization will be conducted immediately after eligibility criteria for the index case have been documented. Index cases and their households will be randomly allocated in blocks of multiples of seven to either the intervention or enhanced standard of care (SoC) group using computer-generated random numbers, stratified by district and by age. For all non-deceased patients, randomisation will take place in or adjacent to the health facility where the index case was either recruited or will be followed up for their routine clinical care. No randomization of currently living index cases should take place in the household where the index case lives.

A study staff member will call a dedicated study telephone number – staffed by someone not linked to any other aspect of the conduct of the study. This person will open consecutive envelopes for each district, containing the randomization schedule in the order of the telephone calls received from each district, and will sign and date the envelope and the contents, record and convey the arm to which each index patient’s household has been allocated as well as the study identification number, study site, date of randomization, age and initials of the index case. The study ID and the allocated arm will be immediately sent by SMS to that site’s study coordinator. An image of that participants and their household’s randomization sheet with its attendant now-opened envelope will be both texted and emailed to the study site and printed and captured in the site files. The original will be retained by the person opening the randomization envelopes.

This is an open study and we cannot blind index cases, their households, or study field research teams to allocation. However, investigator blinding will be maintained until final analysis – no interim analyses which reveal results of the study or provide an indication by group lab results or other outcomes will be provided to site staff or an investigator (apart from the trial statistician). The trial will be unblinded by the trial statistician only to the closed part of the data safety management board (DSMB) meetings, on completion of all trial activities and data cleaning, or if there is agreement to prematurely halt the study because of safety, or other participant or health service concerns, futility, or because the trial overall objective has been met prior to full recruitment or follow up.

### Trial Procedures

**Intervention group (**intensified household contact tracing and treatment support [IHCS]):

*Intensified contact tracing & baseline assessment*
A team comprised of an enrolled nurse and two counsellors will conduct household visits with supervision by a research nurse. An experienced study coordinator at each site will take overall day-to-day management of recruitment and follow up at each site.

**The initial household contact** will be conducted within 14 days of recruitment of each index case allocated to the intervention group. Following provision of information and permission from household head, a brief household survey will be done to assess household poverty status and dwelling characteristics, including tallying windows, external doors and rooms.

During the first household contacts which we will limit to three visits at different times of the day and week to accrue as many household members as possible. Before any individual level data is collected, Research Assistants will obtain individual written informed consent to participate from each household member over 14 years; parental consent for children <18 years to participate in research, and assent from those aged 7 years to 14 years old. We will administer a short questionnaire to each recruited household member that includes: demographic details including relationship and airspace sharing with the recruited index case; risk factors for TB; presence of TB symptoms; and history of previous and current HIV and TB treatment; current quality of life; and employment status and income

In addition, we will question the household head/s about the cost of the index case TB and diagnosis has had on the household. We will attempt to ascertain costs, from the patient household perspective, of a TB diagnosis. If the index case has passed away, the costs related to the funeral and final burial of the body will be ascertained. We will also ascertain the contribution of the ill or deceased person to the total household income. This will be done in a sensitive manner, with interviewers having been trained in grief counselling and aware of changes in emotion of the interviewee, being able to terminate the interview if required. Participants either requesting additional assistance or being seen to require additional assistance will be referred to appropriate care or social assistance.

**Sputum Collection**

To avoid bias related to collecting better quality sputum samples from symptomatic patients, we will collect one spot sputum sample from all household members capable of producing a sputum sample, before we ask about TB symptoms. Prior to producing a sputum specimen household members will be asked to drink a cup of water and rinse out their mouths, then take four deep breaths prior to giving a good cough effort. If small volumes of sputum are obtained, we will allow up to three hours for the collection of a spot sputum specimen. This specimen will be subjected to Xpert MTB/Rif (or the at time-of-study current version of the usual first line TB lab test in the public sector) and liquid mycobacterial culture at public sector laboratories. Single sputum specimens will be at least 3ml and at the lab will be mixed with decontaminant as per standard operating procedures in the two public sector laboratories we will send specimens to - then split into two: Xpert MTB Rif (with subsequent addition of buffer) and immediate liquid culture in the MGIT automated system as per current protocols in public sector mycobacterial laboratories. We will record the cycle threshold number of the Xpert and the time-to-positive in liquid culture. Sputum samples will be immediately stored and transported in coolboxes at 4ºC to public sector laboratories for testing. Anyone who tests positive for *M tuberculosis* will be immediately referred to their chosen public sector clinic for TB treatment initiation with a clear referral letter. For those who are Xpert MTB/Rif negative, HIV-negative and over 5 years of age a tuberculin skin test will be performed with the needle of the syringe parallel to the long axis of the arm at this initial visit and read 48 – 72 hours later using the ball-point pen to mark the widest transverse diameter measured with the reverse calliper method in millimetres. No induration will be documented as 0mm.

**HIV testing**

All household members will be offered HIV testing after counselling, in the first instance using a serial rapid diagnostic test algorithm, the kits for which will be provided provided by the Provincial Departments of Health. Research Assistants trained and accredited in HIV testing and counselling. In line with recommendations in the World Health Organization’s HIV Testing Services Guidelines 2015, a negative A1 test result will diagnose participants as HIV-negative. Positive results to both Tests A1 and the confirmatory A2 test will diagnose participants as HIV-positive. Where results to Test A1 and A2 are discrepant, both tests will be repeated, and followed by a tie-breaker using a laboratory-based 4^th^ Generation ELISA (anticipated in <1 per 1000 participants). Couples testing will be provided if requested.

Children under 18 months of age will have a dried blood spot sample taken for HIV PCR testing as per South African National Guidelines, with testing performed at the National Institute for Communicable Diseases. Results-based post-test counselling will be offered individually in a private setting out-of-earshot and out of sight of other household members.

Participants declining HIV testing may be offered alternative laboratory-based HIV testing on either a blood specimen or an oral fluid specimen using OraQuick, OraSure Technologies –depending on their availability, with communication of results within one week, but if still reluctant to be HIV tested, will be asked to provide an anonymous oral fluid sample for HIV testing for study purposes. Those who request anonymous testing without receipt of results and those who refuse HIV testing will be encouraged to go to a HIV testing facility for testing.

All participants who test HIV positive will be referred to their nearest clinic for initiation of ART. We will draw blood specimen for CD4 count estimates -if still required by National Guidelines as these are expected to change to a test and treat approach in the near future – as soon as the participant is diagnosed with HIV. Participants who test HIV positive will receive supported access to HIV treatment, prearranged by the study at their nearest HIV care clinic.

**Other measures**

Weight and height will be measured in all participants, for children under one year, we will measure their length while recumbent.

Blood pressure will be measured in all adults over 18 years, using standardised method, at least three times with the patient seated using an electronic calibrated device with an interval of at least three minutes between each reading, each reading will be recorded with the time that it was measured. This measure will be repeated at the follow up household visit if the blood pressure is elevated. Any person requiring antihypertensive treatment according to current South African guidelines will be referred to initiation of therapy.

Random finger prick glucose level will be measured in all participants over 30 years of age. Those with measures >10 mmol/l will have an HbA1C taken and referred for treatment if elevated.

**Follow-up household visit (1week after baseline household visit)**
Xpert MTB results and results of laboratory HIV testing (and CD4 counts and HbA1C as appropriate) will be communicated to participants by study staff including registered nurses at a follow-up household visit approximately one-week after the baseline assessment is completed. Participants with microbiologically-confirmed tuberculosis or diagnosed with HIV will be supported to attend their nearest local primary care clinic to register for ongoing care and treatment.

Participants who are older than 5 years of age or test HIV-negative and whose sputum tests are negative for TB and whose tuberculin skin test is negative will be dispensed the first month of IPT by study nurse and then referred for ongoing preventive treatment at their local clinics with a clear referral letter that includes all results. Additionally all those participants who are less than 5 years of age or are HIV-positive and sputum tests are negative for TB will be dispensed the first month of isoniazid preventive therapy by study nurses, Linkage to care will be confirmed by collection of referral cards at study facilities by study staff. Home tracing to encourage re-entry to care will be undertaken on three occasions for participants who do not attend clinics within 2 weeks of referral.

Should any liquid TB culture be resulted positive after this visit, a visit will be made to the few anticipated households from where this result was taken to inform the patient and refer him/her for TB treatment.

| **Age of household member** | **HIV status** | **TST Required** | **Active TB disease** | **TB HH contact** | **IPT initiated** | **Study procedure** |
| --- | --- | --- | --- | --- | --- | --- |
| Child < 5 years | POSITIVE on ART or not | NO | NO | YES | YES | Initiate 1 month IPT |
| Child ≥5 years, adult or adolescent | POSITIVE on ART or not | NO | NO | YES | YES | Initiate 1 month IPT |
| Child < 5 years | NEG | NO | NO | YES | YES | Initiate 1 month IPT |
| Child ≥5 years, adult or adolescent | NEG | YES | NO | YES | YES | Initiate 1 month IPT |

Table 1. IPT initiation for household contacts without TB symptoms

*Final intervention household visit (3 months after baseline household visit)*
All intervention households will be visited once at approximately three months after the initial assessment. This visit will be offered because our previous data suggests that a significant proportion of household members with either TB, or HIV, diagnosed at the baseline household visit, do not access care or treatment. Patients not accessing TB treatment, ART or IPT care will be reassessed and assisted to start appropriate treatment, with study staff attempting to overcome access barriers identified by this group. In addition, we will reassess whether those with hypertension or those diagnosed with type 2 diabetes mellitus were started on appropriate therapy and are at this visit controlled or have a plan for future care. Household members requiring other assistance or care will receive supported access to their local health facility or to the Department of Social Development to access social grants. Moreover, at this visit a TB symptom screen on all available household members not on TB treatment will be done, with investigation of those with symptoms, using Xpert and TB culture. We will not visit the household again to interview household members not present at this single final intervention visit.

All visits will be recorded including the number and level of staff members and the purpose and duration of the visit.

**Non-intervention group (Enhanced standard of care)**
After index cases have been allocated to the non-intervention group at their health facility,, a careful census of the demographic characteristics of each household member will be recorded from the index case. A single contact will be made with the non-intervention index cases at the time of randomisation. Each recruited index case allocated to the non-intervention group will be provided with a referral letter for every household member they identify, and will be requested to give the pre-printed referral letter to each person in their household. The referral letter will contain information about tuberculosis and HIV and will provide recommendation to household members, that they should be screened for TB and HIV, with details of local health facilities where screening and further care (if required) may be accessed. Additionally, the referral letter will contain guidance for health providers, including recommendations that TB screening, HIV testing and further care and prevention services (including ART and TB preventive therapy as required) should be offered as per National guidelines because the individual was exposed to a likely infectious case of TB.

After approximately a week, Study Research Assistants will make a follow up call to the index case or the caregiver of the index case to assess if the referral letters were given to household members and to confirm that any ill household members have accessed their local health facility. If they have been unable to do this, advice on locations of clinics, and other assistance will be provided telephonically. No further contact will be had with non-intervention index cases of households till the final outcome visit.

**All households: final outcome assessment visit**
We will conduct an outcome household visit to all households, irrespective of their study allocation at 15 months after the recruitment of the index case and initial first visit to the household and 12 months after the second follow up visit. Although the study is unblinded, we will attempt to limit research field staff’s knowledge of initial allocation; and in training for and the conduct of the final outcome assessment we will not refer to households as intervention or non-intervention households.

Finding households of index cases can be time consuming, and ideally non-intervention households should be as similar to intervention households as possible. To try and ensure that non-intervention households are not visited by teams who have been searching for a household for longer periods than intervention households where locations are known, 2-6 months prior to the final outcome visit – if resources permit - we will send a study staff team member to locate the non-intervention houses and reintroduce the study to at least one household member. Apart from confirming the address using the what3word App, no data will be recorded.

At the final outcome visit, Research Assistants will revisit all households 1) trace all household participants, ascertain vital status and brief verbal autopsies for those who have died; 2) record episodes of TB and HIV diagnosis and when they occurred, treatment and other hospitalizations from verbal report and inspection of patient-held records; and 3) investigate participants with symptoms of TB (any of: cough, fever, weight loss, night sweats) by collecting sputum for smear microscopy, sputum culture and Xpert; and 4) encourage and offer repeat HIV testing to participants negative or not previously tested at baseline; and 5) conduct a prevalence survey for latent TB infection by testing all children under 14 years old with the tuberculin skin test, read at 48-72 hours later as described previously; 6) a livestock survey will also be done on a sub-sample of 300 households (150 households per site) assessing exposure to domestic pets and livestock and consumption of raw milk.

**In Depth Interviews**

In-depth individual interviews will also be conducted using a semi-structured interview guide. We will conduct interviews with 30 randomly selected households in the intervention arm during the study split by site, and 30 different randomly selected households after all follow-ups have been completed, split by site. We will also do interviews with 30 randomly selected non-intervention households during the study. In total, we aim to conduct 90 interviews across both arms. Interviews will explore household members’ perceptions and acceptability of being visited at home by healthcare personnel and attempt to ascertain in the intervention arm, how they were perceived and if any suggestions in the process of visiting and follow up could be made.

The interviews will be conducted with available household members who are over >18 years by trained study personnel for approximately 30 minutes, in the participant’s language of choice, taped and transcribed. The interviews will be conducted one on one so as to avoid having individual responses influenced by other household members

#### Outcomes

*Primary trial outcomes*

2. Comparison between trial groups of TB FREE SURVIVAL from two months after randomisation – to the final ascertainment visit among all household members identified at baseline – a total of 13 months. We will measure this in two ways; firstly, including TB irrespective of the diagnostic method and secondly, including only bacteriologically-confirmed cases of TB.

*Secondary trial outcomes*

7. Comparison between trial groups of the incidence of all-forms of TB over months 2-15 among household members identified at baseline
8. Comparison between trial groups of the proportion of TB cases with diagnostic delay (defined as number of days between onset of symptoms and diagnosis of anti-tuberculosis treatment) among household members identified at baseline and diagnosed with TB between baseline and month 15
9. Comparison between trial groups of the prevalence of TB infection measured by tuberculin skin test reactivity >10mm at month 15 among household children identified at baseline and aged under 14 years of age. Analysis will be done overall and stratified by age.
10. Comparison between trial groups of the prevalence (reported as a percentage of those tested) of previously undiagnosed and untreated HIV infection among household members identified at baseline and again at month 15.
11. Comparison between trial groups of the cumulative incidence of all-cause mortality among all household members identified at baseline between baseline and month 15.
12. Comparison between trial groups of the cumulative incidence of TB- and HIV-related non-traumatic mortality among all household members identified at baseline between baseline and month 15.
13. Estimation of the cost-effectiveness of the intensified housed TB and HIV case-finding and prevention activities compared to the enhanced standard of care from a societal perspective.

*Exploratory outcomes*

5. Cohort analysis of the incidence of bacteriologically-confirmed, and all-forms of tuberculosis among intervention group household members receiving and not receiving isoniazid preventive therapy stratified by HIV serostatus.
6. Cohort analysis of the proportion of household members in the intervention group with previously undiagnosed HIV infection identified at baseline who successfully initiate antiretroviral therapy by month 15.
7. Estimation of the fraction of TB transmission to household members attributable to household exposure to TB index cases by analysis of whole genome sequences of sputum TB isolates (*Subject to further funding being secured)*

#### Health economics and costing procedures

All costing and economic evaluations will be primarily from the perspective of the health care system to enhance comparability with other studies. However, estimates of household costs of illness and deaths will also be included in certain cost effectiveness measures to reflect a societal perspective. The analysis will also explore the magnitude of household costs, potential for catastrophic costs of HIV and TB, and implications for universal access.

We will use a direct costing approach to assess incremental costs of the ICHF intervention in achieving the primary objective. Costs of diagnostic tests will be obtained from actual expenditures and price lists. Staff costs will be based on public sector remuneration scales for relevant staff, combined with observations, recording time use and services delivered, both to distinguish services from each other and to quantify, cost and exclude research specific activities. Travel costs will be based on logbooks and similar cost allocation approaches. Where any other substantial costs are identified and cannot be directly costed, e.g. management overheads, these will be estimated using step down methods. Household members will be asked at interview about direct and indirect costs (e.g. lost income) of illness and accessing healthcare, and about effect on various household asset holdings (appliances, cellphones etc.) to explore dissaving and potential for catastrophic costs.

The principal cost effectiveness indicators will be incremental costs per person diagnosed with HIV and/or TB, and importantly cost per person linked to HIV and/or TB care through the ACF intervention in each setting, and cost per incident TB case and death avoided, as these could inform immediate, large-scale policy choices. Further indicators such as estimated incremental cost per life year saved or per TB treatment completion will be generated if the frequency and profile of reported clinical outcomes support this. In addition, we will measure health-related quality of life of household members using the EuroQol EQ-5D adjusted for Southern Africa (Maheswaran et al. 2016)

Records of health service utilization (in-patient days and outpatient visits) for each group will be combined with unit costs for each type of care derived from step down costings in relevant facilities to obtain indications of possible cost savings and net costs. Sensitivity analyses will be undertaken to assess strength and robustness of cost effectiveness findings, and aggregated and disaggregated measures of costs will be reported to aid comparisons with other interventions and settings. Incremental cost effectiveness ratios will be compared to South African benchmarks and other measures (e.g. per capita GDP) to facilitate interpretation for policymakers.

### Study Definitions

**Households:** Physical dwelling structures comprising of all rooms under a contiguous roofed area linked by doorways or windows through which air can pass or potentially can pass.

**Household members:** All individuals who shared household airspace for lengthy periods – either slept overnight at least once, or shared at least two meals in the same household as the index case - in the 14 days prior to the index case’s diagnosis of TB

**Incident bacteriologically-confirmed TB** will be defined by a household contact who has documented evidence of at least one of any of the following: sputum smear positive for acid-fast bacilli; or culture with growth of Mycobacterium tuberculosis confirmed on speciation; or positive Xpert MTB/Rif result; or histology or cytology suggestive of TB, regardless of whether TB treatment was initiated. We will restrict incident cases to those diagnosed on a specimen taken at least 2 months after the initial baseline household contact.

**Incident bacteriologically unconfirmed TB** will be defined by a household contact without bacteriologically-confirmed TB but who has at least one of the following: TB treatment registers, or patient held TB treatment cards indicate TB treatment was initiated; or either died or was discharged from hospital with a diagnosis of TB without laboratory confirmation.

**All forms of TB** will be defined in individuals with either bacteriologically-confirmed, or bacteriologically-unconfirmed TB.

**Prevalent *Mycobacterium tuberculosis infection*** will be defined by the presence of skin induration reaction measured to be >10mm using technique recommended by the International Union Against Tuberculosis and Lung Disease at 48-72 hours after subdermal injection of 2 TU in 0.1 ml PPD RT23/Tween 80 in children younger than 7 years of age.

**Prevalent previously undiagnosed HIV infection** will be defined by household members who, at month 15 assessment, test positive for HIV using a serial rapid diagnostic test algorithm infection, having reported never previously tested positive for HIV.

**Prevalent previously undiagnosed and untreated HIV infection** will be defined by household members who, at month 15 assessment, test positive for HIV using a serial rapid diagnostic test algorithm infection and report not currently taking antiretroviral therapy.

### Adverse events

We anticipate only a very small number of adverse events (SAEs), but case definitions, standardised opportuning procedures and a reporting protocol will be in place, and any event deemed to be a serious adverse event will be reported to all IRBs within 7 days of reporting. Additionally, all SAEs will be reviewed by the DSMB.

Events deemed to be serious adverse events and systematically recorded and reported, and will include:

- Misclassification or misinterpretation of HIV or TB results leading to a participant starting treatment in error, or failing to start treating
- Adverse reaction to tuberculosis skin testing, such as ulceration, gangrene or loss of limb, disseminated infection, or death
- Participant withdrawal due to reported stigma, discrimination or breach in confidentiality
- Needlestick injuries to participants or staff
- Episodes of violence or discrimination reported by participants or other household members, or other parties, that is attributed to study contacts or interventions.

### Statistical Methods

##### Sample size

Assuming a mean household size of 5.5 and household TB incidence the mean of that reported for each district, a sample size of 1,200 index cases in each district (2,400) in total, will provide 80% power at an alpha of 0.05 for the study to detect a 30% overall difference in TB free survival between groups with intracluster correlation coefficient of k=0.3. Routine TB data from both sites (Capricorn and Matlosana) suggest that they diagnose and treat at least double this number per annum.

##### Statistical analysis

All statistical analysis will be conducted using R (The R Foundation for Statistical Computing, Vienna). All study data cleaning and analysis procedures will be fully documented in reproducible formats. Final, anonymised study datasets will be made available though an open access data repository based at the University of Liverpool UK to facilitate replication of analysis, and secondary analysis.

All analysis will be done on an intention to treat basis, with denominators comprising of households and household members randomly allocated to trial groups.

We will compare baseline household-, and index case-, and household members-level socio-demographic and clinical characteristics between allocated trial groups using absolute numbers and proportions for categorical variables, and means and standard errors, or medians and interqualitile ranges for continuous variables.

The following situations will be considered unbalance in baseline covariates between the two intervention groups:

- More than 5% difference between the two arms in: % male and proportion of index cases with bacteriologically-confirmed pulmonary tuberculosis.
- More than 5 years difference in mean age.
- More than 1 person difference in mean number of household members

If there is no unbalance in covariates, study conclusions will be based on the unadjusted analysis described below. Otherwise, study conclusions will be based on analysis adjusted for imbalanced covariates as described below.

The primary trial outcome (*TB free survival – both bacteriologically-confirmed TB, and all forms of TB - among household contacts identified at baseline between months 2 and 15*) will be estimated by a rate ratio and 95% confidence interval, with a 2-sided p-value<0.05 considered a statistically significant difference.

Because individuals will be clustered at the household level, we will use negative binomial generalised linear models with generalised estimating equations, accounting for clustered data with robust standard errors. We will assume an exchangeable correlation structure, whereby a correlation is taken to be constant between any two cluster members. As a sensitivity analysis, we will test the independent correlation structure. We will use person-years as an offset to the models to account for different amounts of time participants will be resident at the house. Where imbalance exists between trial groups (as defined above), analysis of outcomes will be adjusted for covariates that demonstrate important differences.

Analysis of secondary outcomes and exploratory outcomes will follow a similar approach, excepting where binomial outcomes are assessed (e.g. proportion of child household members with *MTB* infection, proportion of household members with undiagnosed or untreated HIV), in which case we will use log-binomial models, accounting for clustering with generalised estimating equations and robust standard errors.

The effectiveness of the intervention will be defined in terms of the primary outcome. A societal perspective will be taken for cost effectiveness: system and patient costs will be captured and analysed both jointly and separately to assess to whom benefits primarily accrue (see Economic Statistical Analysis Plan for full details).

6.	Analysis populations

Outcomes will be assessed in two analysis populations: A) all household members who were listed at baseline for whom information is available at final study visit (“baseline cohort”); B) all household members for whom information is available at final study visit irrespective of presence at baseline (“final visit cohort”). Cohort A will be the primary analysis population. Results from cohort B will be used to investigate whether the effect of the intervention extends beyond those exposed to the intervention.

All analyses will be done on an intention-to-treat basis, i.e. data from household members included in populations A and B described above, will be included in the analysis, regardless of whether they underwent all intervention procedures in their trial arm. Household members will be defined as all individuals who either slept overnight at least once, or shared at least two meals in the same household as the index case in the 14 days prior to the index case’s diagnosis.

Additionally, for secondary outcome 1, the analysis population will be restricted to those individuals aged less than 14 years at the time of outcome assessment at 15 months and for secondary outcome 2, the analysis population will be restricted to those individuals with a bacteriologically-confirmed TB diagnosis during the follow-up period.7.	Subgroup analysis

Subgroup analysis by study site (Capricorn District and Botshabelo District) will be conducted for all outcomes, in order to assess whether the effect of the intervention differs between a high (Botshabelo) and lower (Capricorn) TB burden setting.

8.	Recruitment and trial profile

A trial profile figure will be produced illustrating the following, by trial arm:

•	Number of index cases screened for enrolment

•	Number of index cases excluded, with reasons

•	Number of index cases enrolled and randomised

•	Number of household members identified at baseline

•	Of those household members identified at baseline, number whose vital status can be ascertained at 15 months

•	Of those household members identified at baseline, number for whom data on TB diagnosis is available at 15 months

9.	Baseline characteristics and descriptive analyses

Characteristics of index cases, their households and household members identified at baseline will be tabulated and summarised by trial arm. Absolute number and proportions will be used for categorical variables, and means and standard errors, or medians and interquartile ranges will be used for continuous variables.

Characteristics to be summarised will include

Index case characteristics:

•	Age

•	Sex

•	Source of index case

•	Vital status

•	Presence and duration of TB symptoms

•	Treatment received

•	Laboratory confirmation of TB

•	Co-morbidities including HIV

•	Employment status

•	Income

Household characteristics:

•	Number of household members

•	Number of rooms, windows, external doors

•	Household crowding

•	Household construction materials

Household member characteristics

•	Relationship with recruited index case

•	Airspace sharing with recruited index case

•	Presence and duration of TB symptoms

•	History of previous and current HIV treatment

•	History of previous and current TB treatment

•	Employment status

•	Income

10.	Statistical analysis of outcomes

Statistical analysis will be conducted using R (The R Foundation for Statistical Computing, Vienna, Austria) or Stata (StatCorp, College Station, Texas, USA). All analyses will employ statistical methods that allow for within-cluster (household) correlations. For all analyses, a 2-sided p-value<0.05 will be considered a statistically significant difference. No adjustment will be made for multiple testing.

Although baseline imbalance between the two intervention groups is unlikely due to the large sample size of this cluster randomised trial, the following situations will be considered as demonstrating imbalance in baseline covariates:

•	More than 5% difference between the two arms in: % male and proportion of index cases with bacteriologically-confirmed pulmonary tuberculosis.

•	More than 5 years difference in mean age.

•	More than 1 person difference in mean number of household members

If there is no unbalance in covariates, study conclusions will be based on the unadjusted analysis described below. Otherwise, study conclusions will be based on analysis adjusted for imbalanced covariates as described below.

For the primary trial outcome (TB free survival), Poisson regression with random effects to account for clustering at the household level, will be used to estimate a rate ratio, 95% confidence interval and p-value for the effect of the intervention. To allow for the stratified randomisation by district, a fixed effect term for district will be included in the regression model. Where imbalance exists between trial groups (as defined above), these covariates will also be included in the regression model.

Analysis of secondary- and exploratory outcomes will follow a similar approach, except where binomial outcomes are assessed (e.g. proportion of child household members with MTB infection; proportion of household members with undiagnosed or untreated HIV), in which case we will use logistic regression with random effects to account for household clustering. For secondary outcome 5 (community HIV load), we will calculate this as the fraction of HIV-positive individuals with a measured viral load who have detectable viraemia multiplied by the number of individuals testing HIV-positive and divided by the total population with a measured HIV status.

Subgroup analysis will be done by examining stratum-specific (district-specific) rate ratios (or odds ratios for binary outcomes) and by fitting terms for the interaction between trial arm and district in random effects regression models.

Thematic content analysis with an inductive approach will be used to analyse transcripts for obvious and hidden content.

### Study Administration and Monitoring

**IRB Approval**
Ethical approval will be sought from Institutional Review Board of the University of the Witwatersrand, South Africa, the University of Liverpool, UK, and the London School of Hygiene and Tropical Medicine, UK.

**Trial Registration**
The trial protocol (and all subsequent amendments) will be registered on the clinicaltrials.gov website in the South African equivalent prior to commencement of study activities.

**Trial Steering Committee**
We will establish a Trial Steering Committee, comprising of the Trial Principal Investigators, Trial Statistician and Project Manager who will assume responsibility for the trial management and conduct.

**Data and Safety Monitoring Board**
We will establish a Data and Safety and Monitoring Board (DSMB) comprising of external experts, including a statistician experience in cluster-randomised controlled trials in sub-Saharan Africa, a clinical scientist experienced in TB management, and a lay member. The DSMB will meet four times: once prior to commencement of study activities; twice during the 15 month trial activity period, and once following completion of analysis. At each meeting, the DSMB will review a Trial progress report describing rates of recruitment and uptake of trial interventions, outcomes, and serious adverse events recorded. Because of the short duration of study activities, and the low potential for harm, interim analysis will not be performed.

**Good Clinical Practice**
All study staff members will complete study specific clinical trial research training provided by Perinatal HIV Research Unit, and will be required to have certification to demonstrate proficiency to Good Clinical Practice (GCP) Standards.

### Available Infrastructure

***PHRU: Matlosana:*** The North West Department of Health has provided office and clinic space to PHRU, at the Tshepong Hospital in Klerksdorp. Classified as a regional hospital, its immediate catchment area is Matlosana but also takes referrals from further afield. There are four towns and their adjacent townships in Matlosana, with a total population of about 750,000. The Matlosana health sub-district treats about 5000 TB patients each year, diagnosed in its 16 clinics and the hospital. HIV coinfection in adults is 85%, almost 90% of women diagnosed with TB being co-infected with HIV. Adult medicine admits about 40 patients per day, approximately half with infected with HIV and approximately one third of all admissions will either be receiving treatment or diagnosed with TB. PHRU established a research presence seven years ago, and now employs over 130 staff conducting research into several important TB-related areas. We currently recruit MDR patients to clinical trials of novel TB agents at the the purpose-built, drug resistant in-patient TB wards, capable of treating 100 in-patient MDR, and 20 XDR TB cases.

The hospital has a well-equipped accredited NHLS laboratory, capable of doing Xpert MTB/Rif and liquid culture. PHRU has daily courier service to research laboratories in Johannesburg. The Matlosana sub-district diagnoses PHRU has a research presence in all clinics of the Matlosana health sub-district, and in the hospital, there is an active surveillance program identifying all cases of TB either diagnosed through positive laboratory tests or started on empiric TB treatment. It will be relatively easy to include in this study only TB cases identified in the Jouberton township immediately adjacent to the Tshepong Hospital.

In the **Polokwane Municipality in Capricorn Health District, Limpopo Province,** we will establish a study office in the Department of Medicine, University of Limpopo, under the leadership of Prof Ratsele. The populations size is larger and demographics are similar to those of Matlosana, but the epidemiology of HIV and TB is very different. As PHRU has done in Klerksdorp, we will support and develop the creation of a research facility in Polokwane. Not only will we ensure that both the study staff and those in the Department of medicine receive research and GCP training, but also will assist Prof Ratsele in applying for other grants, research projects and turning research findings into publications. We have already conducted a pilot study of household contact tracing, recruiting 130 index cases in 23 clinics around Thouyandou, in Limpopo, are implementing a cluster randomized trial in the area of Vhembe around Makhado, and are conducting further research in household contact tracing with Dr David Dowdy in the Vhembe and Waterberg Health Districts in Limpopo. This study will include the local teaching hospital as an active partner.

### Ethical Considerations

This trial offers a number of important health benefits to participants in the intervention group, including early diagnosis and supported access to TB and HIV care, which are known to be problematic and expensive for both the health system and individuals. We feel there is insufficient data to support recommendations for household contact tracing. Suggesting this trial and the inclusion of an enhanced standard of care group is required. Additionally, individuals allocated to the control group with receive support to access to care beyond that usually provided through the routine health system. Moreover, the trial will provide important information to assist the development of improved HIV and TB services in local communities and in South Africa.

After a thorough assessment of risks, the investigators consider the risk of unintentional harm to participants is minimal, and outweighed by the positive benefits. However, there are a small number of ethical considerations that we will take care to minimise.

Research participants will be clearly informed that participation in the study is entirely voluntary and will be assured that their clinical care will not in any way be affected by their decision to participate or not participate. Participants will be free to withdraw from the study at any time they choose and if they do so, will be referred to the nearest primary health care centre for ongoing care. No financial incentives will be offered for participation as this may influence care-seeking behaviour. There will be no financial costs to the participant for taking part.

Household tracing has the potential to inadvertently disclose confidential clinical information such as TB or HIV status and perpetrate stigma and discrimination, although previous cluster randomised trials in sub-Saharan Africa have demonstrated that these potential risks are largely unfounded, with trial withdrawal due to stigma or discrimination extremely uncommon. Disclosures of HIV status and TB status have been clearly shown to be beneficial to household members and partners. Nevertheless, the trial team will take extensive steps to ensure that recruited index cases are fully aware of the implications of participation in terms of household tracing and follow-up, and will be supported to disclose results to members of their household by trained research assistants and study nurses.

HIV and TB testing will use South African National guideline procedures, which have been well-validated in terms of diagnostic accuracy and have extremely low potential for misdiagnosis.

### Dissemination

Following unblinding and final analysis, study results will be formally presented to the District and Provincial health departments.

Study results will be submitted for publication in a peer-reviewed journal with an emphasis of public health and tuberculosis soon after completion of analysis. Results will also be presented at an international scientific conference to facilitate dissemination.
