## Supplementary material for "Household contact tracing with intensified tuberculosis and HIV screening in South Africa: a cluster randomised trial": S2 Questionnaires

### Index Case Report Form C

| | |
|---|---|
| Study ID | ______________________________ |
| Study site: | ◯ Mangaung<br>◯ Capricorn |
| Name of area | ______________________________ |
| C01. Form completed by | ◯ Index case<br>◯ Next-of-kin<br>◯ Parent/Legal guardian |

#### Demographics

| | |
|---|---|
| C02. Date | ______________________________ |
| C03. Site where participant was first recruited or diagnosed | ◯ Clinic<br>◯ Hospital out-patient<br>◯ Hospital in-patient<br>◯ TB Laboratory |
| C04. Date of Admission | ______________________________ |
| C05. Date of Discharge | ______________________________ |
| C06. Deceased? | ◯ Yes<br>◯ No |
| C07. Date of death | ______________________________ |
| C08. Name of clinic or hospital | ______________________________ |
| C09. Age of index case in: | ◯ Years<br>◯ Months |
| C10. __ __ years | ______________________________ |
| C11. __ __ months | ______________________________ |
| C12. Gender of index case | ◯ Male<br>◯ Female |

| | |
|---|---|
| C13. Employment status | ◯ Currently employed<br>◯ Not employed<br>◯ Student<br>◯ Other |
| C13.a. Describe Other | __________________________________ |
| C14. Employment industry | ◯ Agriculture<br>◯ Mining<br>◯ Construction<br>◯ Manufacturing<br>◯ Transportation<br>◯ Hospitality<br>◯ Unemployed<br>◯ Student<br>◯ Other |
| C14.a. Describe Other | __________________________________ |
| C15. Index case monthly income type | ◯ Salary<br>◯ Wage<br>◯ Grant<br>◯ No income |
| C16. Index case monthly income amount | ◯ R0<br>◯ R1-R999<br>◯ R1000-R4999<br>◯ R5000-R9999<br>◯ R10000+ |

**Basis of diagnosing TB (tick all that apply)**

| | |
|---|---|
| C17. Sputum Xpert | ◯ Positive ◯ Negative<br>◯ Not done |
| C18. Sputum smear | ◯ Positive ◯ Negative<br>◯ Not done |
| C19. Sputum culture | ◯ Positive ◯ Negative<br>◯ Not done |
| C20. Chest X-Ray | ◯ Positive ◯ Negative<br>◯ Not done |
| C21. (Children only) | ◯ Clinical diagnosis |
| C78. LF-LAM | ◯ Positive<br>◯ Negative<br>◯ Invalid<br>◯ Not done |
| C22. How sick was the index case at recruitment?<br><br>(Karnofsky score) __ __% | __________________________________ |

C23. Weight __ __ __kg

______________________________

C24. Height __ __ __cm

______________________________

C25. Blood pressure __ __ __/__ __ __

______________________________
(Sys/Dia)

C26. MUAC __ __mm

______________________________

C27. Fingerprick HGT __ __.__mmol/L

______________________________
(Changed to 25yrs as of 20 July 2017)

C28. Did you draw blood for HbA1C?

◯ Yes
◯ No

C29. HbA1C result

______________________________

#### General Health

C30. Does the index case currently smoke tobacco?

◯ Yes (current smoker)
◯ No (but smoked previously)
◯ Never smoked

C31. Does/did the index case smoke tobacco:

◯ Daily
◯ Weekly
◯ Monthly
◯ Every few months
◯ Only when drinking alcohol
◯ Never smoked

C32. How many cigarettes does/did the index case smoke per day? __ __

______________________________

C33. How old was the index case when he/she started smoking tobacco? __ __

______________________________

C34. Does/did the index case drink alcohol?

◯ Yes
◯ No

C35. How many units of alcohol does/did the index case drink?

__ __ __ units/week

______________________________
(If NONE record 000. If NOT KNOWN record 999)

C36. Does the index case drink traditional beer?

◯ Yes
◯ No

C37. HIV sero-status of index case?

◯ Positive
◯ Negative
◯ Unknown

C38. Most recent CD4 count __ __ __ __

(9999 if CD4 count never done)

C39. Date taken

C40. Is/was the index case taking ART for the treatment of HIV?

(If deceased were they taking ART before they died?)

◯ Yes
◯ No

C41. Date started ARV's

C42. Has the index case been referred for ARV initiation?

◯ Yes
◯ No
◯ Already taking ART

C43. Has the index case tested for HIV before?

◯ Yes
◯ No

C44. When was the last HIV test done?

◯ In the last year
◯ Over 1 year ago
◯ Never tested for HIV

**Symptom Screen**

C45. Cough (days) __ __ __

(If NO record 000. If NOT KNOWN record 999)

C46. Productive cough (days) __ __ __

(If NO record 000. If NOT KNOWN record 999)

C47. Blood stained or bloody cough (days) __ __ __

(If NO record 000. If NOT KNOWN record 999)

C48. Loss of weight (days) __ __ __

(If NO record 000. If NOT KNOWN record 999)

C49. Night sweats (days) __ __ __

(If NO record 000. If NOT KNOWN record 999)

C50. Fever (days) __ __ __

(If NO record 000. If NOT KNOWN record 999)

C51. Has index lived with any person who has had TB in the last 3 years?

◯ Yes
◯ No
◯ Don't know

| | |
|---|---|
| C52. Did this person with TB at home, take TB treatment? | ◯ Yes<br>◯ No<br>◯ Don't know |
| C53. Is this person who had TB deceased? | ◯ Yes<br>◯ No |

#### Co-Morbid Conditions in the Index Case

| | |
|---|---|
| C54. Diabetes | ◯ No Diabetes<br>◯ Diabetes - Insulin injection<br>◯ Diabetes - Oral treatment only<br>◯ Diabetes - Not treated |
| C55. Hypertension | ◯ No Hypertension<br>◯ Hypertension - Oral treatment only<br>◯ Hypertension - Not Treated |

#### TB Results
#### (Only include specimens that were taken two months before to two months after enrolment)

| | |
|---|---|
| C56. GeneXpert | ◯ M Tuberculosis detected<br>◯ M Tuberculosis not detected<br>◯ Not done |
| C56. Date taken (GeneXpert) | __________________________ |
| C57. GeneXpert Rif result | ◯ Rifampicin resistance detected<br>◯ Rifampicin resistance not detected<br>◯ Not done |
| C58. Sputum smear result | ◯ Negative/Not seen<br>◯ Scanty<br>◯ 1+<br>◯ 2+<br>◯ 3+<br>◯ Not done |
| C58. Date taken (Sputum smear) | __________________________ |
| C59. Sputum culture result | ◯ Positive for M tuberculosis<br>◯ Negative<br>◯ Contaminated or lab problem<br>◯ Not done |
| C59. Date taken (Sputum culture) | __________________________ |

#### Drug Sensitivity from any result in file or in the lab for this episode of TB

C60. Isoniazid
◯ Resistant
◯ Sensitive
◯ Not done

C61. Rifampicin
◯ Resistant
◯ Sensitive
◯ Not done

C62. If resistant to Isoniazid and Rifampicin, resistant to second-line drugs?
◯ Yes
◯ No

C63. Resistant to
__________________________________

C63. Resistant to
__________________________________

C63. Resistant to
__________________________________

C64. Other biological specimen
◯ Collected
◯ Not collected

C65. Specify specimen
__________________________________

C66. Date taken (other biological specimen)
__________________________________

C67. Result
◯ TB (probable/confirmed)
◯ No TB
◯ Unknown

C68. CXR
◯ Done
◯ Not done

C69. Result
☐ Cavities
☐ Bilateral disease

C70. Record anatomical site of TB
◯ Pulmonary (lungs/pleura)
◯ Extra-pulmonary

C71. Site(s)
__________________________________

C72. Has the index case previously been diagnosed with or treated for TB?
◯ Yes
◯ No

#### For THIS episode of TB in the Index Case

C73. TB treatment started?
◯ Yes
◯ No

C74. Start date
__________________________________

C75. Reason for not starting TB treatment
◯ Deceased
◯ Defaulted
◯ Other

C75. State Other
__________________________________

#### Form Completion

C76. Date form C completed
__________________________________

C77. Name of person completing this form
__________________________________

#### Audit Trail

Name of Data Entry Person
__________________________________

Date of Data Entry
__________________________________
(Kindly change the date every time a change is made to the data)

Form Version: 1.2 dated 22 October 2018
__________________________________

### Index Census For Household Members Form D

| | |
|---|---|
| D01. Completed by | ◯ Index case<br>◯ Next-of-kin<br>◯ Parent/Legal guardian |
| D02. Child headed household | ◯ Yes<br>◯ No |
| D03. Total number of household members __ __ | ______________________________ |
| D04 Name | ______________________________ |
| D05 Participant ID<br><br>(kindly enter the complete ID, inclusive of Study ID) | ______________________________<br>(x-xx-xxxx-xx format. Changed 19-10-2017) |
| D06 Age | ______________________________ |
| D06 Age in: | ◯ Years<br>◯ Months<br>◯ UNK |
| D07 Gender | ◯ Male<br>◯ Female |
| D08 Relationship to index | ◯ Husband/Wife/Partner<br>◯ Son/Daughter/Stepchild<br>◯ Brother/Sister<br>◯ Parent (mother/father)<br>◯ Parent-in-law<br>◯ Grandparent<br>◯ Grandchild<br>◯ Other |
| D08.1. Describe other | ______________________________ |
| D09 Confirm this person is alive | ◯ Yes<br>◯ No |
| D10 New household member Entered household in past 15 months | ◯ Yes<br>◯ No<br>◯ N/A |
| D10.1. Does this person have TB symptoms now? | ◯ Yes<br>◯ No<br>◯ N/A |
| D11 Cough __ __ | ______________________________ |

| | |
|---|---|
| D12 Weight loss __ __ | ______________________ |
| D13 Fever __ __ | ______________________ |
| D14 Night sweats __ __ | ______________________ |
| D15 Is this person taking TB treatment now? | ◯ Yes<br>◯ No<br>◯ Don't know |
| D16 Is this person taking ART? | ◯ Yes<br>◯ No<br>◯ Don't know<br>◯ N/A |
| D17. To which group was this person randomized to? | ◯ Household intervention strategy<br>◯ Letter strategy |
| D18. Randomization number __-__ __-R__ __ __ __-01 | ______________________ |
| D19. How many letters were given to household? __ __ | ______________________ |

**Form Completion**

| | |
|---|---|
| D20. Date form D completed | ______________________ |
| D21. Time completed | ______________________ |
| D22. Name of person completing this form | ______________________ |
| Some required questions have been skipped!<br>Kindly review questions<br>D03, D18 & D19 before proceeding.... | |

**Audit Trail**

| | |
|---|---|
| Name of Data Entry Person | ______________________ |
| Date of Data Entry | ______________________<br>(Kindly change the date every time a change is made to the data) |

### Household Census At Baseline Form G

| | |
|---|---|
| G01. Housing type | ◯ Brick/Concrete house<br>◯ Apartment<br>◯ Traditional hut/Dwelling<br>◯ Garage<br>◯ 1 room<br>◯ Shack<br>◯ Other |
| Describe other | __________________ |
| G02. How many dining rooms? | __________________ |
| G03. How many kitchens? | __________________ |
| G04. How many bed rooms? | __________________ |
| G05. How many living rooms? | __________________ |
| G06. How many bathrooms/toilet? | __________________ |
| G07. How many other rooms? [Type 1] | __________________ |
| G08. How many other rooms? [Type 2] | __________________ |
| G09. Total rooms in household | __________________ |
| G10. How many windows are there in the household? | __________________ |
| G11. How many doors are there in the household? | __________________ |
| G12. What are your main sources of energy for cooking? [Primary] | ☐ Electricity<br>☐ Gas<br>☐ Wood<br>☐ Coal<br>☐ Paraffin<br>☐ Candles<br>☐ Other |

| | |
|---|---|
| G12. What are your main sources of energy for cooking? [Secondary] | ☐ Electricity<br>☐ Gas<br>☐ Wood<br>☐ Coal<br>☐ Paraffin<br>☐ Candles<br>☐ Other |
| G13. What are your main sources of energy for heating? [Primary] | ☐ Electricity<br>☐ Gas<br>☐ Wood<br>☐ Coal<br>☐ Paraffin<br>☐ Candles<br>☐ Other |
| G13. What are your main sources of energy for heating? [Secondary] | ☐ Electricity<br>☐ Gas<br>☐ Wood<br>☐ Coal<br>☐ Paraffin<br>☐ Candles<br>☐ Other |
| G14. What are your main sources of energy for lighting? [Primary] | ☐ Electricity<br>☐ Gas<br>☐ Wood<br>☐ Coal<br>☐ Paraffin<br>☐ Candles<br>☐ Other |
| G14. What are your main sources of energy for lighting? [Secondary] | ☐ Electricity<br>☐ Gas<br>☐ Wood<br>☐ Coal<br>☐ Paraffin<br>☐ Candles<br>☐ Other |
| G15. How many people smoke tobacco inside the household? __ __ __ | ______________________________<br>(If NONE record 000. If NOT KNOWN record 999) |
| G16. Access to piped water | ◯ Piped water inside the dwelling<br>◯ Piped water within 100m of household<br>◯ Piped water inaccessible<br>◯ Piped water inside the yard<br>◯ Piped water on a community stand < 100m of household |
| G17. Main source of water for drinking/cooking/cleaning | ◯ Government piped water<br>◯ Borehole<br>◯ Spring<br>◯ Dam/pool/stagnant water<br>◯ River or stream<br>◯ Other |
| Describe other | ______________________________ |

| | |
|---|---|
| G18. Toilet facilities | ◯ Flush toilet<br>◯ Chemical toilet<br>◯ Pit toilet<br>◯ Bucket toilet<br>◯ Other |
| Describe other | __________________________________ |

**Deceased**

| | |
|---|---|
| G19. Did anyone who lived in this house die in the past year? | ◯ Yes<br>◯ No |
| G20. How many people living in the household died in the last year? | __________________________________ |

**Record the 4 most recent deaths in the household, with the most recent FIRST**

| | |
|---|---|
| G21.1. Age at death [Death 1] | __________________________________ |
| G21.1. Gender [Death 1] | ◯ Male<br>◯ Female |
| G23.1. Died with TB? [Death 1] | ◯ Yes<br>◯ No<br>◯ Don't know |
| G24.1.Died whilst taking TB treatment? [Death 1] | ◯ Yes<br>◯ No<br>◯ Don't know |
| G25.1. Did they ever test HIV-positive? [Death 1] | ◯ Yes<br>◯ No<br>◯ Don't know |
| G26.1. Taking ART at death? [Death 1] | ◯ Yes<br>◯ No<br>◯ Don't know |

**Record the 4 most recent deaths in the household, with the most recent FIRST**

| | |
|---|---|
| G21.2. Age at death [Death 2] | __________________________________ |
| G22.2. Gender [Death 2] | ◯ Male<br>◯ Female |
| G23.2. Died with TB? [Death 2] | ◯ Yes<br>◯ No<br>◯ Don't know |
| G24.2.Died whilst taking TB treatment? [Death 2] | ◯ Yes<br>◯ No<br>◯ Don't know |

G25.2. Did they ever test HIV-positive? [Death 2]
◯ Yes
◯ No
◯ Don't know

G26.2. Taking ART at death? [Death 2]
◯ Yes
◯ No
◯ Don't know

**Record the 4 most recent deaths in the household, with the most recent FIRST**

G21.3. Age at death [Death 3]
__________________________________

G22.3. Gender [Death 3]
◯ Male
◯ Female

G23.3. Died with TB? [Death 3]
◯ Yes
◯ No
◯ Don't know

G24.3.Died whilst taking TB treatment? [Death 3]
◯ Yes
◯ No
◯ Don't know

G25.3. Did they ever test HIV-positive? [Death 3]
◯ Yes
◯ No
◯ Don't know

G26.3. Taking ART at death? [Death 3]
◯ Yes
◯ No
◯ Don't know

**Record the 4 most recent deaths in the household, with the most recent FIRST**

G21.4. Age at death [Death 4]
__________________________________

G22.4. Gender [Death 4]
◯ Male
◯ Female

G23.4. Died with TB? [Death 4]
◯ Yes
◯ No
◯ Don't know

G24.4.Died whilst taking TB treatment? [Death 4]
◯ Yes
◯ No
◯ Don't know

G25.4. Did they ever test HIV-positive? [Death 4]
◯ Yes
◯ No
◯ Don't know

G26.4. Taking ART at death? [Death 4]
◯ Yes
◯ No
◯ Don't know

#### Form Completion

G27. Date form G completed

__________________________________

G28. Name of person completing this form

__________________________________

Some required questions have been skipped!
Kindly review questions
G01 & G09 before proceeding....

#### Audit Trail

Name of Data Entry Person

__________________________________

Date of Data Entry

__________________________________
(Kindly change the date every time a change is made to the data)

### Baseline Household Member Form H

| | |
|---|---|
| Participant ID | ________________________ |
| Name | ________________________<br>(First 2 letters of First name and first 2 letters of Last name) |
| H01. Has household member consented to be in the study? | ◯ Yes<br>◯ No |
| H02. Date participant enrolled | ________________________ |
| Age of household member in: | ◯ Years<br>◯ Months |
| H03. __ __ years | ________________________ |
| H04. __ __ months | ________________________ |
| Age captured doesn't correlate to that captured in Form D!<br>Kindly review before proceeding... | |
| H05. Gender of index case | ◯ Male<br>◯ Female |
| H06. Relationship to Index case | ◯ Husband/wife/partner<br>◯ Son/daughter/stepchild<br>◯ Brother/sister<br>◯ Parent(mother/father)<br>◯ Parent-in-law<br>◯ Grandparent<br>◯ Grandchild<br>◯ Other |
| Describe other | ________________________ |
| H07. Employment status | ◯ Currently employed<br>◯ Not employed<br>◯ Student<br>◯ Other |

| | |
|---|---|
| H08. Employment industry | ◯ Agriculture<br>◯ Mining<br>◯ Construction<br>◯ Manufacturing<br>◯ Transportation<br>◯ Hospitality<br>◯ Unemployed<br>◯ Student<br>◯ Other |
| H09. Household member monthly income type | ◯ Salary<br>◯ Wage<br>◯ Grant<br>◯ No income |
| H10. Household member monthly income amount | ◯ R0<br>◯ R1-R999<br>◯ R1000-R4999<br>◯ R5000-R9999<br>◯ R10000+ |
| H11. Over the past 3 months have you shared AIR space with index case? | ◯ Yes<br>◯ No |
| H12. Over the past 3 months how much time did you spend with the index case? | ◯ Every now and then<br>◯ Part of the day<br>◯ Most of the day |
| H13. What activities do you do with the index case? | ☐ Meals<br>☐ Watch TV<br>☐ Sleep in the same room<br>☐ Sleep on the same bed<br>☐ Other |
| Describe other | __________________________________ |

#### Quality of Life

| | |
|---|---|
| H14. In general how do you rate your general health | ◯ Very good<br>◯ Good<br>◯ Neither good or bad<br>◯ Poor<br>◯ Very poor |
| H15. Do you have any difficulties doing the usual activities? | ◯ No problems<br>◯ Slight problems<br>◯ Moderate problems<br>◯ Severe problems<br>◯ Unable to do usual activities |

#### General Health

H16. Weight __ __ __.__kg

H17. Height __ __ __cm

H18. Blood pressure __ __ __/__ __ __

Kindly insert leading zero if your reading has only 2 digits.
e.g 120/080, 090/060 etc

H19. MUAC __ __mm

H20. Do you currently smoke tobacco?
- ◯ Yes (currently smoke)
- ◯ No (previously smoked)
- ◯ Never smoked

H21. How often did/do you smoke?
- ◯ Daily
- ◯ Weekly
- ◯ Monthly
- ◯ Every few months
- ◯ Only when drinking alcohol
- ◯ Never smoked

H22. On average, how many cigarettes did/do you smoke per day? __ __ __

(999, if NOT known)

H23. How old were you when you started smoking tobacco? __ __

(99, if NOT known)

H24. Do you drink alcohol now?
- ◯ Yes
- ◯ No

H25. On average, how many units of alcohol; do you drink per week? __ __ __

(999, if NOT known)

H26. Does the household member drink traditional beer?
- ◯ Yes
- ◯ No

H27. What is your HIV status? (before study testing)
- ◯ Positive
- ◯ Negative
- ◯ Not known

H28. Most recent CD4 count __ __ __ __ cells/mm3

(9999, if CD4 count NEVER done)

H29. Date taken

| | |
|---|---|
| H30. Are you taking ART as treatment for HIV? | ◯ Yes<br>◯ No |
| H31. Date ART started | __________________________ |
| H32. If ART not started, has the household member been referred for ARV initiation? | ◯ Yes<br>◯ No |
| H33. Has household member tested for HIV before? | ◯ Yes<br>◯ No |
| H34. When was your last HIV test done? | ◯ In the last year<br>◯ Over 1 year ago<br>◯ Never tested for HIV |

#### Symptom Screen

| | |
|---|---|
| H35. Cough (days) __ __ __ | __________________________<br>(If NO record 000. If NOT KNOWN record 999) |
| H36. Productive cough (days) __ __ __ | __________________________<br>(If NO record 000. If NOT KNOWN record 999) |
| H37. Blood stained or bloody cough (days) __ __ __ | __________________________<br>(If NO record 000. If NOT KNOWN record 999) |
| H38. Loss of weight (days) __ __ __ | __________________________<br>(If NO record 000. If NOT KNOWN record 999) |
| H39. Night sweats (days) __ __ __ | __________________________<br>(If NO record 000. If NOT KNOWN record 999) |
| H40. Fever (days) __ __ __ | __________________________<br>(If NO record 000. If NOT KNOWN record 999) |

#### Co-Morbid Conditions

| | |
|---|---|
| H41. Diabetes | ◯ No Diabetes<br>◯ Diabetes - Insulin injection<br>◯ Diabetes - Oral treatment only<br>◯ Diabetes - Not treated |
| H42. Hypertension | ◯ No Hypertension<br>◯ Hypertension - Oral treatment only<br>◯ Hypertension - Not Treated |

#### Investigations

H43. Sputum collected?
- ◯ Yes
- ◯ No

H43.1. Date of collection

__________________________________

H44. Time of collection

__________________________________

H44.1. AM/PM
- ◯ AM
- ◯ PM

H45. GeneXpert
- ◯ M Tuberculosis detected
- ◯ M Tuberculosis not detected
- ◯ Not done

H46. Date taken (GeneXpert)

__________________________________

H47. GeneXpert Rif result
- ◯ Rifampicin resistance detected
- ◯ Rifampicin resistance not detected
- ◯ Not done

H48. Sputum smear result
- ◯ Negative/Not seen
- ◯ Scanty
- ◯ 1+
- ◯ 2+
- ◯ 3+
- ◯ Not done

H49. Date taken (Sputum smear)

__________________________________

H50. Sputum culture result
- ◯ Positive for M tuberculosis
- ◯ Negative
- ◯ Contaminated or lab problem
- ◯ Not done

H51. Date taken (Sputum culture)

__________________________________

#### Drug Sensitivity from any result in file or in the lab for this episode of TB

H52. Isoniazid
- ◯ Resistant
- ◯ Sensitive
- ◯ Not done

H53. Rifampicin
- ◯ Resistant
- ◯ Sensitive
- ◯ Not done

H54 If resistant to Isoniazid and Rifampicin, resistant to second-line drugs?
- ◯ Yes
- ◯ No

| | |
|---|---|
| H55. Resistant to | __________________ |
| H55. Resistant to | __________________ |
| H55. Resistant to | __________________ |
| H56. Fingerprick HGT __ __.__mmol/L | __________________ |
| H57. Did you draw blood for HbA1C? | ◯ Yes<br>◯ No |
| H58. HbA1C result __.__% | __________________ |

**TST Testing**

| | |
|---|---|
| H59. TST placed | __________________ |
| H60. TST read | __________________ |
| H61. Induration? | ◯ Yes<br>◯ No |
| H62. Diameter __ __mm | __________________ |
| H63. Pre-existing liver disease? | ◯ Yes<br>◯ No |
| H64. History of severe alcohol use? | ◯ Yes<br>◯ No |
| H65. Symptoms of burning feet and weakness in feet and legs? | ◯ Yes<br>◯ No |
| H66. Was a prescription written? | ◯ Yes<br>◯ No |
| Reason, prescription not written | __________________ |
| H67. Was 1 month of treatment dispensed? | ◯ Yes<br>◯ No |
| Reason, treatment not dispensed | __________________ |
| H68. Date first dose started | __________________ |

| | |
|---|---|
| H69. HIV Rapid test done? | ◯ Yes<br>◯ No |
| H70. Result | ◯ Positive<br>◯ Negative<br>◯ Invalid |
| H71. Confirmatory test done? | ◯ Yes<br>◯ No |
| H72. Result | ◯ Positive<br>◯ Negative<br>◯ Invalid |
| H73. Was an ELISA done? | ◯ Yes<br>◯ No |
| H74. Result | ◯ Positive<br>◯ Negative<br>◯ Invalid |
| H75. Children under 18 months, had an HIV PCR? | ◯ Yes<br>◯ No |
| H76. Result | ◯ Positive<br>◯ Negative<br>◯ Invalid |
| H77. OraQuick? | ◯ Yes<br>◯ No |
| H78. Result | ◯ Positive<br>◯ Negative<br>◯ Invalid |
| H79. Blood drawn for ELISA? | ◯ Yes<br>◯ No |
| H80. Result | ◯ Positive<br>◯ Negative<br>◯ Invalid |
| H81. DNA PCR? | ◯ Yes<br>◯ No |
| H82. Result | ◯ Positive<br>◯ Negative<br>◯ Invalid |
| H83. Anonymous oral fluid? | ◯ Yes<br>◯ No |
| H84. Result | ◯ Positive<br>◯ Negative<br>◯ Invalid |

H85. Final HIV status today

◯ Positive
◯ Negative
◯ Inconclusive (Must retest in two weeks)

H86. Referred for ART?

◯ Yes
◯ No

H87. Blood drawn for CD4

______________________________

H89. CD4 result __ __ __ __ cells/mm3

______________________________

#### Form Completion

H90. Date form H completed

______________________________

H91. Name of person completing this form

______________________________

#### Audit Trail

Name of Data Entry Person

______________________________

Date of Data Entry

______________________________
(Kindly change the date every time a change is made to the data)

### Household Census At 3 Month Followup Form I

Participant ID
________________________________

Name
________________________________
(First 2 letters of First Name and first 2 letters of Last Name)

I01. Who provided information for the CRF?
◯ Index case
◯ Next-of-kin
◯ Parent/Legal guardian

I02. Child headed household
◯ Yes
◯ No

I03. Total number of household members __ __
________________________________

I04. Name
________________________________

I05. Study ID : [record_id]-
________________________________

I06. Age
________________________________

I06. Age in
◯ Years
◯ Months

I07. Gender
◯ Male
◯ Female

I08. Relationship to index
◯ Husband/Wife/Partner
◯ Son/Daughter/Stepchild
◯ Brother/Sister
◯ Parent (mother/father)
◯ Parent-in-law
◯ Grandparent
◯ Grandchild
◯ Other

I08.1. Describe other
________________________________

I09. Confirm this person is alive
◯ Yes
◯ No

I10. Left this household since baseline census?
◯ Yes
◯ No
◯ N/A

| | |
|---|---|
| I11. Does this person have TB symptoms now? | ◯ Yes<br>◯ No<br>◯ N/A |
| I12. Cough __ __ | __________________ |
| I13. Weight loss __ __ | __________________ |
| I14. Fever __ __ | __________________ |
| I15. Night sweats __ __ | __________________ |
| I16. Is this person taking TB treatment now? | ◯ Yes<br>◯ No<br>◯ N/A<br>◯ Don't know |
| I17. Is this person taking ART? | ◯ Yes<br>◯ No<br>◯ N/A<br>◯ Don't know |

#### New people living in the household

| | |
|---|---|
| I18.01. Name | __________________ |
| I19.01. Study ID : [record_id]- | __________________ |
| I20.01. Age | __________________ |
| I21.01. Gender | ◯ Male<br>◯ Female |
| I22.01. Relationship to index | ◯ Husband/Wife/Partner<br>◯ Son/Daughter/Stepchild<br>◯ Brother/Sister<br>◯ Parent (mother/father)<br>◯ Parent-in-law<br>◯ Grandparent<br>◯ Grandchild<br>◯ Other |
| I22.01.1. Describe other | __________________ |
| I23.01. Is this person is alive | ◯ Yes<br>◯ No |
| I24.01. New household member since baseline census? | ◯ Yes<br>◯ No |

| | |
|---|---|
| Does this person have TB symptoms now? | ◯ Yes<br>◯ No<br>◯ N/A |
| I25.01. Cough __ __ | ________________________ |
| I26.01. Weight loss __ __ | ________________________ |
| I27.01. Fever __ __ | ________________________ |
| I28.01. Night sweats __ __ | ________________________ |
| I29.01. Is this person taking TB treatment now? | ◯ Yes<br>◯ No<br>◯ N/A<br>◯ Don't know |
| I30.01. Is this person taking ART? | ◯ Yes<br>◯ No<br>◯ N/A<br>◯ Don't know |
| I18.02. Name | ________________________ |
| I19.02. Study ID : [record_id]- | ________________________ |
| I20.02. Age | ________________________ |
| I21.02. Gender | ◯ Male<br>◯ Female |
| I22.02. Relationship to index | ◯ Husband/Wife/Partner<br>◯ Son/Daughter/Stepchild<br>◯ Brother/Sister<br>◯ Parent (mother/father)<br>◯ Parent-in-law<br>◯ Grandparent<br>◯ Grandchild<br>◯ Other |
| I22.02.1. Describe other | ________________________ |
| I23.02. Is this person is alive | ◯ Yes<br>◯ No |
| I24.02. New household member since baseline census? | ◯ Yes<br>◯ No |

| | |
|---|---|
| Does this person have TB symptoms now? | ◯ Yes<br>◯ No<br>◯ N/A |
| I25.02. Cough __ __ | ______________________ |
| I26.02. Weight loss __ __ | ______________________ |
| I27.02. Fever __ __ | ______________________ |
| I28.02. Night sweats __ __ | ______________________ |
| I29.02. Is this person taking TB treatment now? | ◯ Yes<br>◯ No<br>◯ N/A<br>◯ Don't know |
| I30.02. Is this person taking ART? | ◯ Yes<br>◯ No<br>◯ N/A<br>◯ Don't know |
| I18.03. Name | ______________________ |
| I19.03. Study ID : [record_id]- | ______________________ |
| I20.03. Age | ______________________ |
| I21.03. Gender | ◯ Male<br>◯ Female |
| I22.03. Relationship to index | ◯ Husband/Wife/Partner<br>◯ Son/Daughter/Stepchild<br>◯ Brother/Sister<br>◯ Parent (mother/father)<br>◯ Parent-in-law<br>◯ Grandparent<br>◯ Grandchild<br>◯ Other |
| I22.03.1. Describe other | ______________________ |
| I23.03. Is this person is alive | ◯ Yes<br>◯ No |
| I24.03. New household member since baseline census? | ◯ Yes<br>◯ No |

| | |
|---|---|
| Does this person have TB symptoms now? | ◯ Yes<br>◯ No<br>◯ N/A |
| I25.03. Cough __ __ | __________________________________ |
| I26.03. Weight loss __ __ | __________________________________ |
| I27.03. Fever __ __ | __________________________________ |
| I28.03. Night sweats __ __ | __________________________________ |
| I29.03. Is this person taking TB treatment now? | ◯ Yes<br>◯ No<br>◯ N/A<br>◯ Don't know |
| I30.03. Is this person taking ART? | ◯ Yes<br>◯ No<br>◯ N/A<br>◯ Don't know |
| I18.04. Name | __________________________________ |
| I19.04. Study ID : [record_id]- | __________________________________ |
| I20.04. Age | __________________________________ |
| I21.04. Gender | ◯ Male<br>◯ Female |
| I22.04. Relationship to index | ◯ Husband/Wife/Partner<br>◯ Son/Daughter/Stepchild<br>◯ Brother/Sister<br>◯ Parent (mother/father)<br>◯ Parent-in-law<br>◯ Grandparent<br>◯ Grandchild<br>◯ Other |
| I22.04.1. Describe other | __________________________________ |
| I23.04. Is this person is alive | ◯ Yes<br>◯ No |
| I24.04. New household member since baseline census? | ◯ Yes<br>◯ No |

Does this person have TB symptoms now?
◯ Yes
◯ No
◯ N/A

I25.04. Cough __ __
______________________________

I26.04. Weight loss __ __
______________________________

I27.04. Fever __ __
______________________________

I28.04. Night sweats __ __
______________________________

I29.04. Is this person taking TB treatment now?
◯ Yes
◯ No
◯ N/A
◯ Don't know

I30.04. Is this person taking ART?
◯ Yes
◯ No
◯ N/A
◯ Don't know

#### Deceased

I31. Did anyone who lived in this household die in the past year?
◯ Yes
◯ No

I32. How many people died in the last year? __ __
______________________________

I33.1. Age at death __ __ years
______________________________

I34.1. Gender
◯ Male
◯ Female

I35.1. Died with TB?
◯ Yes
◯ No
◯ N/A
◯ Don't know

I36.1. Died whilst taking TB treatment?
◯ Yes
◯ No
◯ N/A
◯ Don't know

I37.1. Did they ever test HIV-positive?
◯ Yes
◯ No
◯ N/A
◯ Don't know

| | |
|---|---|
| I38.1. Taking ART at death? | ◯ Yes<br>◯ No<br>◯ N/A<br>◯ Don't know |
| I33.2. Age at death __ __ years | __________________ |
| I34.2. Gender | ◯ Male<br>◯ Female |
| I35.2. Died with TB? | ◯ Yes<br>◯ No<br>◯ N/A<br>◯ Don't know |
| I36.2. Died whilst taking TB treatment? | ◯ Yes<br>◯ No<br>◯ N/A<br>◯ Don't know |
| I37.2. Did they ever test HIV-positive? | ◯ Yes<br>◯ No<br>◯ N/A<br>◯ Don't know |
| I38.2. Taking ART at death? | ◯ Yes<br>◯ No<br>◯ N/A<br>◯ Don't know |
| I33.3. Age at death __ __ years | __________________ |
| I34.3. Gender | ◯ Male<br>◯ Female |
| I35.3. Died with TB? | ◯ Yes<br>◯ No<br>◯ N/A<br>◯ Don't know |
| I36.3. Died whilst taking TB treatment? | ◯ Yes<br>◯ No<br>◯ N/A<br>◯ Don't know |
| I37.3. Did they ever test HIV-positive? | ◯ Yes<br>◯ No<br>◯ N/A<br>◯ Don't know |
| I38.3. Taking ART at death? | ◯ Yes<br>◯ No<br>◯ N/A<br>◯ Don't know |

I33.4. Age at death __ __ years

I34.4. Gender
◯ Male
◯ Female

I35.4. Died with TB?
◯ Yes
◯ No
◯ N/A
◯ Don't know

I36.4. Died whilst taking TB treatment?
◯ Yes
◯ No
◯ N/A
◯ Don't know

I37.4. Did they ever test HIV-positive?
◯ Yes
◯ No
◯ N/A
◯ Don't know

I38.4. Taking ART at death?
◯ Yes
◯ No
◯ N/A
◯ Don't know

#### Form Completion

I39. Date form I completed

I40. Name of person completing this form

#### Audit Trail

Name of Data Entry Person

Date of Data Entry

(Kindly change the date every time a change is made to the data)

### Month 3 Household Member Form J

| | |
|---|---|
| Participant ID | ______________________ |
| Name | ______________________<br>(First 2 letters of First Name and first 2 letters of Last Name) |

#### Vital Status and TB Outcomes

| | |
|---|---|
| J01. Is the household member alive? | ◯ Yes<br>◯ No |
| J01.01 Age | ______________________ |
| J01.02. Age in | ◯ Years<br>◯ Months |
| J02. Date of death | ______________________ |
| J03. Household member currently taking TB treatment? | ◯ Yes<br>◯ No (but should be taking)<br>◯ Not required to take TB treatment (currently not taking) |
| J03.a. Showed the GREEN treatment card and study team confirmed current treatment? | ◯ Yes<br>◯ No (but should be taking) |
| J04. Type of treatment? | ◯ Rifafour<br>◯ Other |
| J04.a. Describe other | ______________________ |
| J05. Date last treatment dispensed | ______________________ |
| J06. If household member not taking treatment, what is the main reason for not taking treatment? | ◯ Could not travel to clinic to get more treatment<br>◯ Clinic ran out of treatment<br>◯ Tired of medicines<br>◯ Felt healthy<br>◯ Felt too sick<br>◯ Side effects (treatment making participant sick)<br>◯ Did not have food<br>◯ Religious beliefs<br>◯ Started traditional medicine or traditional healing<br>◯ Doctor told participant to stop<br>◯ Other |
| J06.a. Describe other | ______________________ |

#### ART

| | |
|---|---|
| J07. Household member currently taking ART? | ◯ Yes<br>◯ No (but should be taking)<br>◯ Not required to take ART (currently not taking) |
| J08. Showed current ART treatment card and study team confirmed current treatment? | ◯ Yes<br>◯ No (but should be taking) |
| J09. Type of ART? | ◯ FDC<br>◯ Other |
| J09.a. Describe other | __________________________________ |
| J10. Date last ART treatment dispensed | __________________________________ |
| J11. If household member not taking treatment, what is the main reason for not taking ART? | ◯ Could not travel to clinic to get more treatment<br>◯ Clinic ran out of treatment<br>◯ Tired of medicines<br>◯ Felt healthy<br>◯ Felt too sick<br>◯ Side effects (treatment making participant sick)<br>◯ Did not have food<br>◯ Religious beliefs<br>◯ Started traditional medicine or traditional healing<br>◯ Doctor told participant to stop<br>◯ Only took ART while pregnant<br>◯ Other |
| J11.a. Describe other | __________________________________ |

#### IPT

| | |
|---|---|
| J12. Household member currently taking IPT? | ◯ Yes<br>◯ No (but should be taking)<br>◯ Not required to take IPT (currently not taking) |
| J13. Showed current IPT treatment card and study team confirmed current treatment? | ◯ Yes<br>◯ No (but should be taking) |
| J14. Date IPT dispensed this month | __________________________________ |
| J15. If household member not taking treatment, what is the main reason for not taking IPT? | ◯ Could not travel to clinic to get more treatment<br>◯ Clinic ran out of treatment<br>◯ Tired of medicines<br>◯ Felt healthy<br>◯ Felt too sick<br>◯ Side effects (treatment making participant sick)<br>◯ Did not have food<br>◯ Religious beliefs<br>◯ Started traditional medicine or traditional healing<br>◯ Doctor told participant to stop<br>◯ Other |

J15.a. Describe other

________________________________

#### Diabetes

J16. Household member currently on diabetes medication?

- ◯ Yes
- ◯ No (but should be taking)
- ◯ Not required to take diabetes treatment (currently not taking)

J17. Type of diabetes treatment?

- ◯ Gliclazide
- ◯ Metformin
- ◯ Insulin
- ◯ Told not to have diabetes treatment
- ◯ Not known

J18. Date last diabetes treatment dispensed

________________________________

J19. If not on diabetes treatment, HGT today: __ __.__mmol/L

________________________________

J20. If household member not taking treatment, what is the main reason for not taking diabetes treatment?

- ◯ Could not travel to clinic to get more treatment
- ◯ Clinic ran out of treatment
- ◯ Tired of medicines
- ◯ Felt healthy
- ◯ Felt too sick
- ◯ Side effects (treatment making participant sick)
- ◯ Did not have food
- ◯ Religious beliefs
- ◯ Started traditional medicine or traditional healing
- ◯ Doctor told participant to stop
- ◯ Other

J20.a. Describe other

________________________________

#### Hypertension

J21. Household member taking hypertension treatment?

- ◯ Yes
- ◯ No (but should be taking)
- ◯ Not required to take hypertension treatment (currently not taking)

J22. Type of hypertension treatment?

- ◯ Hydrochlorthiazide
- ◯ Other

J22.a. Describe other

________________________________

J23. Date last hypertension treatment dispensed

________________________________

J24.1. BP recording [1] __ __ __/__ __ __

________________________________

J24.2. BP recording [2] __ __ __/__ __ __

________________________________

J24.3. BP recording [3] __ __ __/__ __ __

________________________________

J25. If household member not taking treatment, what is the main reason for not taking hypertension treatment?

◯ Could not travel to clinic to get more treatment
◯ Clinic ran out of treatment
◯ Tired of medicines
◯ Felt healthy
◯ Felt too sick
◯ Side effects (treatment making participant sick)
◯ Did not have food
◯ Religious beliefs
◯ Started traditional medicine or traditional healing
◯ Doctor told participant to stop
◯ Other

J25.a. Describe other

________________________________

#### Symptom Screen

J26. Cough (days) __ __ __

________________________________
(If NO record 000. If NOT KNOWN record 999)

J27. Productive cough (days) __ __ __

________________________________
(If NO record 000. If NOT KNOWN record 999)

J28. Blood stained or bloody cough (days) __ __ __

________________________________
(If NO record 000. If NOT KNOWN record 999)

J29. Loss of weight (days) __ __ __

________________________________
(If NO record 000. If NOT KNOWN record 999)

J30. Night sweats (days) __ __ __

________________________________
(If NO record 000. If NOT KNOWN record 999)

J31. Fever (days) __ __ __

________________________________
(If NO record 000. If NOT KNOWN record 999)

J32. Other (days) __ __ __

________________________________
(If NO record 000. If NOT KNOWN record 999)

J32.a. Describe other

________________________________

J33. Weight today __ __ __kg

________________________________

#### Investigations

J34. Sputum collected?
- ◯ Yes
- ◯ No

J35. Date of collection ________________________

J36. Time of collection ________________________

J36.1. AM/PM
- ◯ AM
- ◯ PM

J37. GeneXpert
- ◯ M Tuberculosis detected
- ◯ M Tuberculosis not detected
- ◯ Not done

J38. Date taken (GeneXpert) ________________________

J39. GeneXpert Rif result
- ◯ Rifampicin resistance detected
- ◯ Rifampicin resistance not detected
- ◯ Not done

J40. Sputum smear result
- ◯ Negative/Not seen
- ◯ Scanty
- ◯ 1+
- ◯ 2+
- ◯ 3+
- ◯ Not done

J41. Date taken (Sputum smear) ________________________

J42. Sputum culture result
- ◯ Positive for M tuberculosis
- ◯ Negative
- ◯ Contaminated or lab problem
- ◯ Not done

J43. Date taken (Sputum culture) ________________________

#### Drug Sensitivity from any result in file or in the lab for this episode of TB

J44. Isoniazid
- ◯ Resistant
- ◯ Sensitive
- ◯ Not done

J45. Rifampicin
- ◯ Resistant
- ◯ Sensitive
- ◯ Not done

J46. If resistant to Isoniazid and Rifampicin, resistant to second-line drugs?
- ◯ Yes
- ◯ No

J47.1. Resistant to

__________________________________

J47.2. Resistant to

__________________________________

J47.3. Resistant to

__________________________________

#### Form Completion

J48. Date form J completed

__________________________________

J49. Name of person completing this form

__________________________________

#### Audit Trail

Name of Data Entry Person

__________________________________

Date of Data Entry

__________________________________
(Kindly change the date every time a change is made to the data)

### Household Census at Month 15 Followup Form K-Default

Participant ID

______________________
(Format: 0-00-0000-00)

K01: This index case was recruited in

______________________

K02: Household was:

- ◯ Located
- ◯ Not Located

K03: Who provided information for the CRF

- ☐ Index Case
- ☐ Head of the household
- ☐ Other senior household member

K04: Child-headed household

- ◯ Yes
- ◯ No

K05: Number of household members

______________________

Please complete form "K - Household Visits"

Please complete form "K - New Household Members"

K34: Did anyone who lived in this house die in the past 15 months

- ◯ Yes
- ◯ No

K35: How many people died in the past 15 months?

______________________

Please complete form, "K-Household Deaths"

K42: Housing type (check only one response)

- ◯ Brick/concrete house
- ◯ Apartment / flats
- ◯ Traditional hut/dwelling
- ◯ Garage
- ◯ 1 Room
- ◯ Shack
- ◯ Other

Specify Other

______________________

K43: How many DINING ROOMS are there in the household?

______________________

K44: How many KITCHENS are there in the household?

______________________

K45: How many BEDROOMS are there in the household?

______________________

K46: How many LIVING ROOMS are there in the household? __________________________________

K47: How many BATHROOMS/TOILETS are there in the household? __________________________________

K48: Other rooms Type 1 __________________________________

K48: How many [k48] are there in the household? __________________________________

K49: Other rooms Type 2 __________________________________

K49: How many [k49] are there in the household? __________________________________

K50: How many rooms are there in the household in total (Sum K43-K49) __________________________________

K51: How many windows are there in the household? __________________________________

K52: How many doors are there in the household that lead outside __________________________________

K53: What are your main sources of energy for COOKING

- ◯ Electricity
- ◯ Gas
- ◯ Wood
- ◯ Coal
- ◯ Paraffin
- ◯ Candles
- ◯ Other

(Primary)

Specify Other __________________________________

K53: What are your main sources of energy for COOKING

- ◯ Electricity
- ◯ Gas
- ◯ Wood
- ◯ Coal
- ◯ Paraffin
- ◯ Candles
- ◯ Other

(Secondary)

Specify Other __________________________________

| | |
|---|---|
| K54: What are your main sources of energy for HEATING | ◯ Electricity<br>◯ Gas<br>◯ Wood<br>◯ Coal<br>◯ Paraffin<br>◯ Candles<br>◯ Other<br>(Primary) |
| Specify Other | __________________________ |
| K54: What are your main sources of energy for HEATING | ◯ Electricity<br>◯ Gas<br>◯ Wood<br>◯ Coal<br>◯ Paraffin<br>◯ Candles<br>◯ Other<br>(Secondary) |
| Specify Other | __________________________ |
| K55: What are your main sources of energy for LIGHTING | ◯ Electricity<br>◯ Gas<br>◯ Wood<br>◯ Coal<br>◯ Paraffin<br>◯ Candles<br>◯ Other<br>(Primary) |
| Specify Other | __________________________ |
| K55: What are your main sources of energy for LIGHTING | ◯ Electricity<br>◯ Gas<br>◯ Wood<br>◯ Coal<br>◯ Paraffin<br>◯ Candles<br>◯ Other<br>(Secondary) |
| Specify Other | __________________________ |
| K56: How many people smoke tobacco inside the household?<br>Total people: | __________________________<br>(000 = none & 99 = Don't know) |
| K57: For how many hours a day is there someone smoking inside the household? | __________________________ |

K58: Main source of water for drinking/cooking/cleaning

- ◯ Government piped water
- ◯ Borehole
- ◯ Spring
- ◯ Dam/pool/stagnant water
- ◯ River or stream
- ◯ Other

Specify Other

__________________________________

K59: If HH has access to piped water, what kind

- ◯ Piped water inside the dwelling
- ◯ Piped water inside the yard
- ◯ Piped water in a community stand < 100m of household
- ◯ Piped water within >100m of household but not inaccessible
- ◯ Piped water inaccessible

K60: Toilet facilities

- ◯ Flush toilet
- ◯ Chemical toilet
- ◯ Pit toilet
- ◯ Bucket toilet
- ◯ Other

Specify Other

__________________________________

K61: Does your household have any of the following and how many?

- ☐ Refrigerator (working)
- ☐ Radio (working)
- ☐ Phone/landline (working)
- ☐ Internet (working)
- ☐ Microwave (working)
- ☐ Cellphone (working)
- ☐ Smart phone (working)
- ☐ Computer (working)
- ☐ Washing machine (working)
- ☐ Satellite dish/DSTV (working)

K61: Refrigerator No.

__________________________________

K61: Radio No.

__________________________________

K61: Phone/landline No.

__________________________________

K61: Internet No.

__________________________________

K61: Microwave No.

__________________________________

K61: Cellphone No.

__________________________________

K61: Smart phone No.

__________________________________

| | |
|---|---|
| K61: Computer No. | __________________ |
| K61: Washing machine No. | __________________ |
| K61: Satellite dish/DSTV No. | __________________ |
| K62: Does household have a vehicle? (Working) If yes, how many | ◯ Yes<br>◯ No |
| K63: If Yes, Number of vehicles | __________________ |
| K64: If YES, is the vehicle more than 7 years old: | ◯ Yes<br>◯ No |
| K65: Does household have a Television (working) | ◯ Large, flat screen<br>◯ TV is at least 7 years old<br>◯ No TV |
| K66: Does household have a stove (working) | ◯ Gas stove<br>◯ Electric stove<br>◯ Paraffin stove<br>◯ Wood stove or cook with wood |
| K67: In the last 15 months have members of the household bought any of the household goods and appliances we have been asking you about?<br>If YES, which items in the last 15 months: | ☐ Refrigerator<br>☐ Radio<br>☐ TV<br>☐ Phone<br>☐ Car<br>☐ DSTV Dish<br>☐ Cellphone<br>☐ Smart phone<br>☐ Stove<br>☐ Microwave<br>☐ Computer<br>☐ Internet<br>☐ Other |
| Specify Other | __________________ |
| L68: If YES, how much has been spent on those items in the last 15 months | __________________<br>(R (ZAR)) |
| K69: In the last 15 months have members of the household bought any equipment for their work or business? | ◯ Yes<br>◯ No |
| L70: If YES, which items in the last 15 months: | ☐ Tools<br>☐ Car<br>☐ Farm animals<br>☐ Computer<br>☐ Other |

| | |
|---|---|
| Specify Other | __________________________ |
| L71: If YES, how much has been spent on those items in the last 15 months | __________________________ |
| K72: If the household has an emergency, how much money would the household have in its savings account for urgent expenses? | ◯ No savings<br>◯ < R100<br>◯ R100 - R500<br>◯ R500 - 1000<br>◯ R1000 - R2000<br>◯ R2000 - 5000<br>◯ > R5000 |
| K73: In the past 15 months, children in the household | ◯ Are in or have completed tertiary education<br>◯ Are in or have completed high school<br>◯ Are in or have completed primary school<br>◯ Are in or have completed preschool/cheche<br>◯ No children |
| K74: Does anyone who lives in the household ever go to bed hungry because there is no food available | ◯ Yes<br>◯ No |
| K75: If YES, how many days per week | __________________________ |
| K76: How many people in the household receive an income (including pensions and grants)? | __________________________ |
| K77: What is the total monthly income of the household for all people? | __________________________ |
| K78: Household occupational status<br>Has any other household member or person sending regular money lost or gained employment/earning ability in the last 15 months | ◯ Yes<br>◯ No |
| Lost<br>Household member study ID number | __________________________ |
| Lost<br>Household member study ID number | __________________________ |
| Lost<br>Household member study ID number | __________________________ |
| Or<br>Number of person/s sending money | __________________________ |
| Gained<br>Household member study ID number | __________________________ |
| Gained<br>Household member study ID number | __________________________ |
| Gained<br>Household member study ID number | __________________________ |

Or
Number of person/s sending money ________________

K79: In the past 15 months, has the household added or renovated a room/s
◯ Yes
◯ No

K80: If YES, what was the building cost of this renovation
________________
(R (ZAR))

#### HOUSEHOLD ILLNESS

K81: Has any household member not been able to earn money, or missed picking up grant or other money or payment because of an illness?
◯ Yes
◯ No

K82: If YES, Estimate how much money was not earned or collected due to illness
________________
(R (ZAR))

K83: Did any household member borrow money for healthcare to cover costs due to an illness?
◯ Yes
◯ No
◯ Don't know

K84: If YES, how much
________________
(R (ZAR))

K85: If YES, from whom was the money borrowed
◯ Family
◯ Neighbour
◯ Friend
◯ Boyfriend/Girlfriend
◯ Bank
◯ Stokvel
◯ Machonisa/Loan Shark
◯ Employer
◯ Didn't borrow
◯ Other

Specify Other ________________

K86: Did any Household member have to sell/pawn an asset/ livestock to pay for healthcare?
◯ Yes
◯ No

K87: If YES, what assets/livestock? ________________

K88: Has the household had to reduce spending because of a sickness affecting a household member/s?
◯ Yes
◯ No

K89: What did the household reduce spending on the most
1)

- ◯ 0. No reduction in spending
- ◯ 1. Food
- ◯ 2. Entertainment (DSTV or other/soccer matches etc)
- ◯ 3. Petrol/transport/taxi fare
- ◯ 4. School expenditure/uniforms/books/outings
- ◯ 5. Clothing
- ◯ 6. Alcohol/ other drinks
- ◯ 7. Airtime
- ◯ 8. Savings
- ◯ 9. Helper/Domestic
- ◯ 10. Payments on furniture, other household goods bought on instalments, car, other
- ◯ 11. Other

---

K89: What did the household reduce spending on the most
2)

- ◯ 0. No reduction in spending
- ◯ 1. Food
- ◯ 2. Entertainment (DSTV or other/soccer matches etc)
- ◯ 3. Petrol/transport/taxi fare
- ◯ 4. School expenditure/uniforms/books/outings
- ◯ 5. Clothing
- ◯ 6. Alcohol/ other drinks
- ◯ 7. Airtime
- ◯ 8. Savings
- ◯ 9. Helper/Domestic
- ◯ 10. Payments on furniture, other household goods bought on instalments, car, other
- ◯ 11. Other

---

K90: On what other things did they reduce spending?
1)

- ◯ 0. No reduction in spending
- ◯ 1. Food
- ◯ 2. Entertainment (DSTV or other/soccer matches etc)
- ◯ 3. Petrol/transport/taxi fare
- ◯ 4. School expenditure/uniforms/books/outings
- ◯ 5. Clothing
- ◯ 6. Alcohol/ other drinks
- ◯ 7. Airtime
- ◯ 8. Savings
- ◯ 9. Helper/Domestic
- ◯ 10. Payments on furniture, other household goods bought on instalments, car, other
- ◯ 11. Other

---

K90: On what other things did they reduce spending?
2)

- ◯ 0. No reduction in spending
- ◯ 1. Food
- ◯ 2. Entertainment (DSTV or other/soccer matches etc)
- ◯ 3. Petrol/transport/taxi fare
- ◯ 4. School expenditure/uniforms/books/outings
- ◯ 5. Clothing
- ◯ 6. Alcohol/ other drinks
- ◯ 7. Airtime
- ◯ 8. Savings
- ◯ 9. Helper/Domestic
- ◯ 10. Payments on furniture, other household goods bought on instalments, car, other
- ◯ 11. Other

K90: On what other things did they reduce spending? 3)

- ◯ 0. No reduction in spending
- ◯ 1. Food
- ◯ 2. Entertainment (DSTV or other/soccer matches etc)
- ◯ 3. Petrol/transport/taxi fare
- ◯ 4. School expenditure/uniforms/books/outings
- ◯ 5. Clothing
- ◯ 6. Alcohol/ other drinks
- ◯ 7. Airtime
- ◯ 8. Savings
- ◯ 9. Helper/Domestic
- ◯ 10. Payments on furniture, other household goods bought on instalments, car, other
- ◯ 11. Other

K91: If someone in the household died, how much did funeral arrangements cost this household

- ☐ Did not have a funeral

K91: Amount

______________________________
(R (ZAR))

K92: On a scale of 1 to 5, in which 1 is no impact and 5 is very serious impact, to what extent has the TB illness affected the family costs and income?

- ◯ No impact
- ◯ Little impact
- ◯ Moderate impact
- ◯ Serious impact
- ◯ Very serious impact

K93: Have any HH members been sent to other households because yours could not cope since the illness started?

- ◯ Yes
- ◯ No

K94: If YES, How many members

______________________________

K95: Have any learners/ or students in the household dropped out from learning since the illness started?

- ◯ Yes
- ◯ No

K94: If YES, How many members

______________________________

K97: Date form completed

______________________________

K98: Name of person completing this form

______________________________

#### Audit Trail

Name of Data Entry Person

______________________________

Date of Data Entry

______________________________
(Kindly change the date every time a change is made to the data)

### Household Census at Month 15 Form K-Household Visits

| | |
|---|---|
| Participant ID | ______________ (Format: 0-00-0000-00) |
| K06: Name | ______________ |
| K06: Surname | ______________ |
| K07: Study ID | ______________ |
| K08: Age | ◯ Years ◯ Months |
| K08: Age | ______________ |
| K09: Sex | ◯ Male ◯ Female |
| K10: Is this person alive | ◯ 1=Yes<br>◯ 2=No |
| K11: Left this household in the past 15 months | ◯ 1=Yes<br>◯ 2=No<br>◯ 3=NA |
| K12: Does this person have TB symptoms now? | ◯ Yes<br>◯ No |
| K13: Cough (of any duration) | ◯ 1. Present<br>◯ 2. Not present<br>◯ 3. Not applicable |
| K14: Weight loss | ◯ 1. Present<br>◯ 2. Not present<br>◯ 3. Not applicable |
| K15: Fever | ◯ 1. Present<br>◯ 2. Not present<br>◯ 3. Not applicable |
| K16: Night sweats | ◯ 1. Present<br>◯ 2. Not present<br>◯ 3. Not applicable |
| K17: Is this person taking TB treatment? | ◯ 1=Yes<br>◯ 2=No<br>◯ 3=NA<br>◯ 4=Don't know |

| | |
|---|---|
| K18: Is this person taking ART | ◯ 1=Yes<br>◯ 2=No<br>◯ 3=NA<br>◯ 4=Don't know |
| K19: Has participant consented to be included in the study | ◯ 1=Yes<br>◯ 2=No |

#### Audit Trail

| | |
|---|---|
| Name of Data Entry Person | ________________________________ |
| Date of Data Entry | ________________________________<br>(Kindly change the date every time a change is made to the data) |

### Household Census at Month 15 Followup Form K-New Household Members

| | |
|---|---|
| Participant ID | ________________ (Format: 0-00-0000-00) |
| K20: Name | ________________ |
| K20: Surname | ________________ |
| K21: Study ID | ________________ |
| K22: Age | ◯ Years ◯ Months |
| K22: Age | ________________ |
| K23: Sex | ◯ Male ◯ Female |
| K24: Relationship to Index | ◯ 1=Husband/wife/partner<br>◯ 2=Son/daughter/stepchild<br>◯ 3=Brother/sister<br>◯ 4=Parent(mother/father)<br>◯ 5=Parent-in-law<br>◯ 6=Grandparent<br>◯ 7=Grandchild<br>◯ 8=Other |
| K25: Is this person alive | ◯ 1=Yes<br>◯ 2=No |
| K26: New Household member entered in past 15 months | ◯ 1=Yes<br>◯ 2=No<br>◯ 3=NA |
| K26: Date they started living | ________________ |

**Does this person have TB symptoms now?**

| | |
|---|---|
| K27: Cough (of any duration) | ◯ 1. Present<br>◯ 2. Not Present<br>◯ 3. Not applicable |
| K28: Weight loss | ◯ 1. Present<br>◯ 2. Not Present<br>◯ 3. Not applicable |
| K29: Fever | ◯ 1. Present<br>◯ 2. Not Present<br>◯ 3. Not applicable |

| | |
|---|---|
| K30: Night sweats | ◯ 1. Present<br>◯ 2. Not Present<br>◯ 3. Not applicable |
| K31: Has this person taken TB treatment in the last 15 months | ◯ 1=Yes<br>◯ 2=No<br>◯ 3=NA<br>◯ 4=Don't know |
| K31: Start date of TB treatment | __________________________________ |
| K32: Is this person taking ART | ◯ 1=Yes<br>◯ 2=No<br>◯ 3=NA<br>◯ 4=Don't know |
| K33: Has participant consented to be included in the study | ◯ 1=Yes<br>◯ 2=No |

#### Audit Trail

| | |
|---|---|
| Name of Data Entry Person | __________________________________ |
| Date of Data Entry | __________________________________<br>(Kindly change the date every time a change is made to the data) |

### Household Census at Month 15 Followup Form
### K-Household Deaths

Participant ID

______________________________
(Format: 0-00-0000-00)

K36: Age at death

______________________________

K37: Gender

◯ Male
◯ Female

K38: Date of death (estimate date if not known)

______________________________

K38: Died with likely TB, not on treatment?

◯ Yes
◯ No
◯ Don't know

K39: Died whilst taking TB treatment?

◯ Yes
◯ No
◯ Don't know

K40: Did they ever test HIV-positive?

◯ Yes
◯ No
◯ Don't know

K41: Taking ART at death?

◯ Yes
◯ No
◯ Don't know

**Audit Trail**

Name of Data Entry Person

______________________________

Date of Data Entry

______________________________
(Kindly change the date every time a change is made to the data)

### Month 15 Household Member Form L

| | |
|---|---|
| i. Household was | ◯ Identified<br>◯ Not identified |
| ii. Date of Interview | ______________ |
| iii. Participant ID:<br>[d5] | ______________ |
| iv. Name :<br>[d4] | ______________<br>(First 2 letters of First Name and first 2 letters of Last Name) |
| v. Age:<br>[d6] | ______________ |
| v. Age in | ◯ Years<br>◯ Months |
| vi. Who was interviewed to get data on this household member? | ◯ This household member<br>◯ Another household member/Index case from this household<br>◯ Someone else(specify) |
| vi. Specify | ______________ |
| vii. Household member was: | ◯ Identified<br>◯ Not able to be contacted |

#### Health Information

| | |
|---|---|
| L01. Is the household member alive today? | ◯ Yes<br>◯ No |
| L02. Death confirmed by | ◯ Inspection of Death certificate<br>◯ Verbal report (from whom)<br>◯ Other (provide details) |
| L02.01. Verbal report from: | ______________ |
| L02.02. Other, provide details: | ______________ |
| L03. Date of death | ______________ |

| | |
|---|---|
| L04. If not alive what was the cause of death? (select all that apply.) Verbal response from family member is fine | ☐ TB<br>☐ HIV<br>☐ Diarrhoea<br>☐ Pneumonia<br>☐ Meningitis<br>☐ Accident(vehicle or other)<br>☐ Violence(stab, gunshot, blunt etc.)<br>☐ Stroke<br>☐ Other |
| L04.01. Describe other | ______________________ |
| L05. Weight (no shoes or jacket) if person is available | ______________________<br>(__ __ __kg) |
| L06. Height if younger than 18yrs | ______________________<br>(__ __ __cm) |
| L07. Blood pressure | ______________________<br>(__ __ __/__ __ __ (record the 3rd reading after 2 prior readings)) |

If in third reading, systolic BP>140 or diastolic BP>90, then refer to the nearest clinic for assessment or review of hypertension

| | |
|---|---|
| L08. MUAC | ______________________<br>(__ __mm) |

#### HIV/ART

| | |
|---|---|
| L09. Has the participant ever been tested for HIV? | ◯ Yes<br>◯ No |
| L10. HIV status before this visit HIV testing | ◯ Positive for HIV<br>◯ Negative (must offer HIV test at this visit)<br>◯ Unable to infer HIV status (must offer HIV test at this visit) |
| L11. Has the participant tested for HIV in the last 15 months? | ◯ Yes<br>◯ No |
| L12. What was the result of the participant's most recent HIV test? | ◯ Positive for HIV<br>◯ Negative<br>◯ Invalid<br>◯ Not tested |

#### ~If HIV-Positive~

| | |
|---|---|
| L13. Is participant currently taking ART drugs for treatment of HIV? | ◯ Yes<br>◯ No |
| L14. Showed current ART treatment card and study team confirmed current treatment? | ◯ Yes<br>◯ No |
| L15. Type of ART? | ◯ FDC<br>◯ Other |
| L15.01. Describe other | __________________________________ |
| L16. When did you first start ART? (estimate is OK) | __________________________________ |
| L17. If participant is HIV positive, but not taking ART, what is the main reason for not taking ART? | ◯ Could not travel to clinic to get more treatment<br>◯ Clinic ran out of treatment<br>◯ Tired of medicines<br>◯ Felt healthy<br>◯ Felt too sick to take ART<br>◯ Side effects (treatment making participant sick)<br>◯ Did not have food<br>◯ Religious beliefs<br>◯ Started traditional medicine or traditional healing<br>◯ Doctor told participant to stop<br>◯ Only took ART while pregnant<br>◯ Other |
| L17.01. Describe other | __________________________________ |
| L18. Has participant ever taken ART drugs for treatment of HIV? | ◯ Yes<br>◯ No |
| L19. Last date ART was dispensed to this patient? | __________________________________ |

#### ~If HIV-Negative~

| | |
|---|---|
| L20. HIV Rapid test done at this study visit? | ◯ Yes<br>◯ No |
| L21. Result of rapid HIV test | ◯ Positive<br>◯ Negative<br>◯ Invalid |
| L22. If rapid test Positive, result of confirmatory rapid test | ◯ Positive for HIV<br>◯ Negative<br>◯ Invalid |
| L23. If discordant or invalid was an ELISA taken? | ◯ Yes<br>◯ No |
| L24. Children younger 18 months, had an HIV PCRat this final study visit? | ◯ Yes<br>◯ No |

| | |
|---|---|
| L25. OraQuick done at this final study visit? | ◯ Yes<br>◯ No |
| L26. Lab result of oral fluid (OraQuick) | ◯ Positive for HIV<br>◯ Negative<br>◯ Invalid |
| L27. Anonymous oral fluid taken at this final study visit? | ◯ Yes<br>◯ No |
| L28. Lab result of oral fluid (Anonymous oral fluid) | ◯ Positive for HIV<br>◯ Negative<br>◯ Invalid |

#### Final Study HIV Status

| | |
|---|---|
| L29. Final HIV status of this participant using all results and all self-reports | ◯ Positive for HIV<br>◯ Negative<br>◯ Inconclusive (Must retest in two weeks)<br>◯ Not tested |

If HIV Positive, draw blood for CD4 count and complete Investigation section

#### TB Disease and Treatment

| | |
|---|---|
| L30. In the past 15 months, has this household member been diagnosed with TB; or, has this household member been prescribed (or told) to take TB treatment?<br><br>(Include any TB diagnosed in index cases, and also TB in HH members diagnosed by the study) | ◯ Yes<br>◯ No |
| L31. If yes how many episodes was this person told s/he had TB in the past 15 months? | ◯ One<br>◯ Two<br>◯ More than two |
| L32. If yes to TB episode, date that the participant was first told that s/he had TB | __________________________________ |
| L33. If YES to TB in the past 15 months, did the study team see a source document that confirmed TB including those diagnosed by the study at baseline?<br><br>(tick all that apply) | ☐ Green clinic TB card<br>☐ Lab result Xpert or culture: M TB detected<br>☐ TB register or other clinic document at local clinic<br>☐ No document able to be found that confirms TB |
| L34. Date this specimen confirming TB diagnosis for episode 1 was collected? | __________________________________ |
| L35. If YES to TB in past 15 months, date first dose of TB treatment taken/started | __________________________________ |
| L36. Date TB treatment completed | __________________________________ |
| L36. or, | ☐ Currently taking TB treatment |

L37. If YES to TB in past 15 months , what treatment was used in the first two months of TB treatment phase ?
(select all that apply)

- ☐ Rifafor (or equivalent four drug single dose tablet/s)
- ☐ A regimen that includes at least three of these TB drugs: Clofazamine; either amikacin or kanamycin; bedaquilin; levofloxacin or moxifloxacin; ethionamide
- ☐ Other regimen

L37.1. Describe other

________________________________

#### Episode 2

L38. Did the participant have a second episode of TB in the past 15 months?

- ◯ Yes
- ◯ No

L39. If YES to second episode of TB, date the participant was first told that s/he had the second episode of TB
(Please estimate if no source document available)

________________________________

L40. If YES to second episode of TB, did the study team see a source document that confirmed TB including those diagnosed by the study at baseline

- ☐ Green clinic TB card
- ☐ Lab result Xpert or culture: M TB detected
- ☐ TB register or other clinic document at local clinic
- ☐ No document able to be found that confirms TB

L41. Date this specimen confirming TB diagnosis for episode 2 was collected/taken?

________________________________

L42. If YES to second episode of TB, what type of TB based on resistance patterns

- ◯ Drug sensitive (no resistance detected)
- ◯ Rifampicin resistance
- ◯ MDR (Rifampicin and INH resistance)
- ◯ XDR

L43. If YES to second episode of TB in past 15 months, date TB treatment first dose taken/started

________________________________

L44. Date TB treatment completed

________________________________

L44. or,

- ☐ Currently taking TB treatment

#### Isoniazid Preventive Therapy (IPT)

L45. Did participant ever take any form of preventive therapy IPT or 3HP either prescribed by the study or a clinic?

- ◯ Yes
- ◯ No

L46. If yes did study team see some written confirmation of IPT treatment?

- ◯ Yes
- ◯ No

L47. Date IPT started

________________________________

L48. Estimate how many months of IPT the participant took over the past 15 months

________________________________
(months)

L49. If took fewer than six months of IPT, what main reason was given for stopping IPT?

- ◯ Could not travel to clinic to get more treatment
- ◯ Clinic ran out of IPT or refused to give IPT
- ◯ Tired of IPT
- ◯ Felt healthy
- ◯ Felt too sick to take IPT
- ◯ Side effects (treatment making participant sick)
- ◯ Did not have food
- ◯ Religious beliefs
- ◯ Started traditional medicine or traditional healing
- ◯ Doctor told participant to stop
- ◯ Other

L49.1. Describe other

________________________________

#### Symptom Screen, Today

L50. Cough (days) __ __ __

________________________________
(If NO record 000. If NOT KNOWN record 999)

L51. Productive cough (days) __ __ __

________________________________
(If NO record 000. If NOT KNOWN record 999)

L52. Blood stained or bloody cough (days) __ __ __

________________________________
(If NO record 000. If NOT KNOWN record 999)

L53. Loss of weight (days) __ __ __

________________________________
(If NO record 000. If NOT KNOWN record 999)

L54. Night sweats (days) __ __ __

________________________________
(If NO record 000. If NOT KNOWN record 999)

L55. Fever (days) __ __ __

________________________________
(If NO record 000. If NOT KNOWN record 999)

L56. Other symptom

________________________________

L56. [I56] (days) __ __ __

________________________________
(If NO record 000. If NOT KNOWN record 999)

If any symptom of TB, collect a sputum sample and complete Investigation section

#### Health Economics

L57. Nature of illness

- ☐ HIV
- ☐ TB
- ☐ No illness
- ☐ Other

L57.01. Describe other

__________________________________

L58. Date the participant started being sick or got diagnosed

__________________________________

L59. Member's occupational status before illness

- ◯ Formal employ (full-time)
- ◯ Formal employ (part-time)
- ◯ Self-employed
- ◯ Informal
- ◯ Piece jobs
- ◯ Student - FET/Tertiary
- ◯ In-school learner
- ◯ Pension
- ◯ Unemployed - Looking for work
- ◯ Unemployed - Not looking for work
- ◯ Pre-school going
- ◯ Other

L59.01. Describe other

__________________________________

L60. Member's occupation status now

- ◯ Formal employ (full-time)
- ◯ Formal employ (part-time)
- ◯ Self-employed
- ◯ Informal
- ◯ Piece jobs
- ◯ Student - FET/Tertiary
- ◯ In-school learner
- ◯ Pension
- ◯ Unemployed - Looking for work
- ◯ Unemployed - Not looking for work
- ◯ Pre-school going
- ◯ Other

L60.01. Describe other

__________________________________

#### ~Visits made for medical help related to that new illness~

L61.01. Visits to primary care clinic

__________________________________
(__ __ visits)

L61.02. Visits to community health centre

__________________________________
(__ __ visits)

L61.03. Visits to TB clinic

__________________________________
(__ __ visits)

L61.04. Visits to VCT centre

__________________
(__ __ visits)

L61.05. Visits to hospital

__________________
(__ __ visits)

L61.06. Visits to GP

__________________
(__ __ visits)

L61.07. Visits to pharmacist

__________________
(__ __ visits)

L61.08. Visits to traditional healer

__________________
(__ __ visits)

L61.09. Visits to "other"

__________________
(__ __ visits)

L61.09.01. Describe other

__________________

L62. Was the participant admitted to hospital(s) for the illness in the past 15 months?

○ Yes
○ No

L62.01.01. Hospital [1st admission]

__________________

L62.01.02. Number of nights stay [1st admission]

__________________

L62.01.03 Date of admission [1st admission]

__________________

L62.02.01. Hospital [2nd admission]

__________________

L62.02.02. Number of nights stay [2nd admission]

__________________

L62.02.03 Date of admission [2nd admission]

__________________

L62.03.01. Hospital [3rd admission]

__________________

L62.03.02. Number of nights stay [3rd admission]

__________________

L62.03.03 Date of admission [3rd admission]

__________________

L62.04. Total number of admissions if more that 3 hospital admissions

__________________________________

L63. How does the participant usually travel to the hospital or clinic to get care?

- ◯ Private car
- ◯ Minibus or other taxi
- ◯ Bus
- ◯ Train
- ◯ Walk
- ◯ Other

L63.01. Describe other

__________________________________

L64. What is the cost of one trip to get care?
R

__________________________________

#### Quality of Life

L65. What do you expect will happen to the quality of life for the people in this household over the next year?

- ◯ Much better than 1 year ago
- ◯ Somewhat better than 1 year ago
- ◯ About the same
- ◯ Somewhat worse than 1 year ago
- ◯ Much worse than 1 year ago

L66. In general your health is

- ◯ Excellent
- ◯ Very good
- ◯ Good
- ◯ Fair
- ◯ Poor

#### Investigations for this visit

L67. Date blood was collected for an HIV ELISA

__________________________________

L68. Result of ELISA

- ◯ Positive for HIV
- ◯ Negative

L69. Date sample was collected for children < 18 months for an HIV PCR

__________________________________

L70. Lab result for HIV PCR

- ◯ Positive for HIV
- ◯ Negative

L71. Date blood drawn for CD4 count

__________________________________

L72. Laboratory CD4 result

__________________________________
(__ __ __ __cells/mm3)

#### Investigations done on this participant

| | |
|---|---|
| L73. Sputum collected?<br>(Sputum must be collected if participant reports any TB symptom) | ◯ Yes<br>◯ No |
| L74. Date of sputum collection<br>(must be atleast 4ml) | __________________ |
| L75. Time of sputum collection | __________________ |
| L76. GeneXpert result of this sputum | ◯ M Tuberculosis detected<br>◯ M Tuberculosis not detected<br>◯ Not done |
| L77. GeneXpert Rif result if M Tuberculosis detected | ◯ Rifampicin resistance detected<br>◯ Rifampicin resistance not detected |
| L78. Sputum smear result | ◯ Negative/Not seen<br>◯ Scanty<br>◯ 1+<br>◯ 2+<br>◯ 3+<br>◯ Not done |
| L79. Sputum culture result | ◯ Positive for M tuberculosis<br>◯ Negative<br>◯ Contaminated or lab problem<br>◯ Not done |
| L80. If sputum culture result was contaminated or lab problem, was a second sputum sample collected? | ◯ Yes<br>◯ No |
| L81. Date of second sputum collection<br>(must be atleast 4ml) | __________________ |
| L82. Time of second sputum collection | __________________ |
| L83. Second sputum culture result | ◯ Positive for M tuberculosis<br>◯ Negative<br>◯ Contaminated or lab problem<br>◯ Not done |
| L84. If second sputum culture result was contaminated or lab problem, was a third sputum sample collected? | ◯ Yes<br>◯ No |
| L85. Date of third sputum collection<br>(must be atleast 4ml) | __________________ |
| L86. Time of third sputum collection | __________________ |

L87. Third sputum culture result

- ◯ Positive for M tuberculosis
- ◯ Negative
- ◯ Contaminated or lab problem
- ◯ Not done

#### Drug Sensitivity from any result in file or in the lab for this episode of TB

L88. Isoniazid

- ◯ Resistant
- ◯ Sensitive
- ◯ Not done

L89. Rifampicin

- ◯ Resistant
- ◯ Sensitive
- ◯ Not done

#### TST Testing (only for children 14 years and younger)

L90. TST placed?

- ◯ Yes
- ◯ No

L91. Date TST placed

______________________________

L92. Date TST read

______________________________

L92. or,

- ☐ TST not read

L93. Induration?

- ◯ Yes
- ◯ No

L94. Transverse diameter of induration __ __mm

______________________________

#### Other Investigations

L95. Finger prick HGT

______________________________
(mmol/L)

L96. Did you draw blood for HbA1c?

- ◯ Yes
- ◯ No

L97. HbA1c result

______________________________
(%)

If more than 5.5%, refer for treatment with results

L98. Diabetes

- ◯ No diabetes
- ◯ Diabetes - Insulin injection
- ◯ Diabetes - Oral treatment only
- ◯ Diabetes - Not treated

L99. Hypertension

◯ No hypertension
◯ Hypertension - Oral treatment only
◯ Hypertension - Not treated

#### Form Completion

L100. Date Form L finally completed (including all Lab results) ______________________________

L101. Initials of person who did visual review ______________________________

#### Audit Trail

Name of Data Entry Person ______________________________

Date of Data Entry ______________________________
(Kindly change the date every time a change is made to the data)

### Untoward Household Event Form M

M01. Date of Contact

__________________________________

M02. Contact person

__________________________________

M03. Date event happened

__________________________________

M04. Time event happened

__________________________________

M04.1. AM/PM

◯ AM
◯ PM

M04.2. Time of day

◯ Morning
◯ Afternoon
◯ Evening

M05. Status of event

◯ New
◯ Ongoing

M06. Location where event occured

◯ At the household
◯ Neighbours house
◯ In the community
◯ At the clinic/hospital
◯ At the shops
◯ Other

M06.1. Describe other

__________________________________

M07. Tick all people that were involved in the event that is being reported

☐ Husband/Wife/Partner
☐ Son/Daughter/Stepchild
☐ Brother/Sister
☐ Parent (mother/father)
☐ Parent-in-law
☐ Grandparent
☐ Grandchild
☐ Neighbours
☐ Community members
☐ Friends
☐ Study staff
☐ Other

M07.1. Describe other

__________________________________

| | |
|---|---|
| M08. Summary of event | ☐ Verbal assault<br>☐ Physical assault<br>☐ Adverse drug event (INH related)<br>☐ Adverse drug event (Non-INH related)<br>☐ Suicide attempt<br>☐ Police intervention<br>☐ Job loss<br>☐ Kicked out of home<br>☐ Stigmatization<br>☐ Other |
| M08.1. Describe other | ______________________________ |
| M09. Provide a detailed description of the event | _____________________________________ |
| M10. Has study PI been informed about the event? | ◯ Yes<br>◯ No |
| M11. Describe the action that has been taken following the event | _____________________________________ |
| M12. Is the event that is being reported related to the intervention? | ◯ Yes<br>◯ No<br>◯ Unclear |
| M13. Outcome | ◯ Resolved<br>◯ Ongoing |

#### Form Completion

| | |
|---|---|
| M14. Date form M completed | ______________________________ |
| M15. Name of person completing this form | ______________________________ |

#### Audit Trail

| | |
|---|---|
| Name of Data Entry Person | ______________________________ |
| Date of Data Entry | ______________________________<br>(Kindly change the date every time a change is made to the data) |

### Study Termination Form N

Participant ID
________________________________

Name
________________________________
(First 2 letters of First Name and first 2 letters of Last Name)

N01. Person terminated from study
◯ Index case
◯ Household member

N01.1. Household member Participant ID
________________________________

N02. Date of termination
________________________________

N03. Participant terminated due to
◯ Death (died during study period)
◯ Lost to follow-up
◯ Relocated
◯ Following PI's decision
◯ Participant/Next-of-kin/Legal guardian withdrew consent
◯ Other

N04. Provide reason for withdrawal of consent
________________________________

N05. Describe other
________________________________

#### Form Completion

N06. Date form N completed
________________________________

N07. Name of person completing this form
________________________________

#### Audit Trail

Name of Data Entry Person
________________________________

Date of Data Entry
________________________________
(Kindly change the date every time a change is made to the data)

### Health Economics Form HE

| | |
|---|---|
| Form completed at: | ◯ Baseline<br>◯ Month 3 |
| i. Start Time | __________________________ |

#### Baseline

| | |
|---|---|
| ii. Date of Interview | __________________________ |
| iii. Status of Index Case | ◯ Alive<br>◯ Deceased |
| iii.i. Date decesaed | __________________________ |
| iv. Date of TB diagnosis | __________________________ |
| v. Who is the main person providing information for this questionnaire? | ◯ Index case<br>◯ Other |
| v.i. Relation to Index case | ◯ Wife/Mother<br>◯ Husband/Father<br>◯ Extended family<br>◯ Son/Daughter<br>◯ Other |
| v.i.i. Describe other | __________________________ |

#### Household Socio-economic Status

| | |
|---|---|
| HE01. How many people in the household receive an income?<br>(including pensions and grants) | __________________________ |
| HE02. Who is the primary income earner in the household? | ◯ Index<br>◯ Wife/Mother<br>◯ Husband/Father<br>◯ Extended family<br>◯ Son/Daughter<br>◯ Other |
| HE02.1 Describe other | __________________________ |

| | |
|---|---|
| HE03. Does your household have any of the following and how many? | ☐ Refrigerator<br>☐ Stove<br>☐ Television<br>☐ Car<br>☐ Radio<br>☐ Phone-landline<br>☐ Internet<br>☐ Microwave<br>☐ Cellphone<br>☐ Smartphone<br>☐ Computer<br>☐ Washing machine<br>☐ Satellite dish/DSTV |
| HE03.01. Number - Refrigirator | __________________________________ |
| HE03.02. Number - Stove | __________________________________ |
| HE03.03. Number - Television | __________________________________ |
| HE03.04. Number - Car | __________________________________ |
| HE03.05. Number - Radio | __________________________________ |
| HE03.06. Number - Phone-landline | __________________________________ |
| HE03.07. Number - Internet | __________________________________ |
| HE03.08. Number - Microwave | __________________________________ |
| HE03.09. Number - Cellphone | __________________________________ |
| HE03.10. Number - Smartphone | __________________________________ |
| HE03.11. Number - Computer | __________________________________ |
| HE03.12. Number - Washing machine | __________________________________ |
| HE03.13. Number - Satellite dish/DSTV | __________________________________ |

| | |
|---|---|
| HE04. Household food production | ☐ Livestock<br>☐ Mielies/Sorghum<br>☐ Vegetables<br>☐ Poultry<br>☐ Other |
| HE04.1. Describe other | __________________________________ |
| HE05. Do you have additional income sent by household members not living with you? | ◯ Yes<br>◯ No |
| HE05.1. How many? | __________________________________ |
| HE06. Amount.R- | __________________________________ |
| HE07. Household members | __________________________________ |

**Household member - 01**

| | |
|---|---|
| HE08. Occupation status | ◯ Formal employ (full-time)<br>◯ Formal employ (part-time)<br>◯ Self-employed<br>◯ Informal<br>◯ Piece jobs<br>◯ Student - FET/Tertiary<br>◯ In-school learner<br>◯ Pension<br>◯ Unemployed - Looking for work<br>◯ Unemployed - Not looking for work<br>◯ Pre-school going<br>◯ Other |
| HE08.1 Describe other | __________________________________ |
| HE09. Amount earned per month | ◯ R0<br>◯ R1-R999<br>◯ R1000-R4999<br>◯ R5000-R9999<br>◯ R10000+ |
| HE10. Source | ◯ Wages/Income<br>◯ Pension<br>◯ Disability grant<br>◯ Child grant<br>◯ Other |
| HE10.1 Describe other | __________________________________ |

#### Household member - 02

HE08. Occupation status

- ◯ Formal employ (full-time)
- ◯ Formal employ (part-time)
- ◯ Self-employed
- ◯ Informal
- ◯ Piece jobs
- ◯ Student - FET/Tertiary
- ◯ In-school learner
- ◯ Pension
- ◯ Unemployed - Looking for work
- ◯ Unemployed - Not looking for work
- ◯ Pre-school going
- ◯ Other

HE08.1 Describe other

__________________________________

HE09. Amount earned per month

- ◯ R0
- ◯ R1-R999
- ◯ R1000-R4999
- ◯ R5000-R9999
- ◯ R10000+

HE10. Source

- ◯ Wages/Income
- ◯ Pension
- ◯ Disability grant
- ◯ Child grant
- ◯ Other

HE10.1 Describe other

__________________________________

#### Household member - 03

HE08. Occupation status

- ◯ Formal employ (full-time)
- ◯ Formal employ (part-time)
- ◯ Self-employed
- ◯ Informal
- ◯ Piece jobs
- ◯ Student - FET/Tertiary
- ◯ In-school learner
- ◯ Pension
- ◯ Unemployed - Looking for work
- ◯ Unemployed - Not looking for work
- ◯ Pre-school going
- ◯ Other

HE08.1 Describe other

__________________________________

HE09. Amount earned per month

- ◯ R0
- ◯ R1-R999
- ◯ R1000-R4999
- ◯ R5000-R9999
- ◯ R10000+

| | |
|---|---|
| HE10. Source | ◯ Wages/Income<br>◯ Pension<br>◯ Disability grant<br>◯ Child grant<br>◯ Other |
| HE10.1 Describe other | __________________________________ |

**Household member - 04**

| | |
|---|---|
| HE08. Occupation status | ◯ Formal employ (full-time)<br>◯ Formal employ (part-time)<br>◯ Self-employed<br>◯ Informal<br>◯ Piece jobs<br>◯ Student - FET/Tertiary<br>◯ In-school learner<br>◯ Pension<br>◯ Unemployed - Looking for work<br>◯ Unemployed - Not looking for work<br>◯ Pre-school going<br>◯ Other |
| HE08.1 Describe other | __________________________________ |
| HE09. Amount earned per month | ◯ R0<br>◯ R1-R999<br>◯ R1000-R4999<br>◯ R5000-R9999<br>◯ R10000+ |
| HE10. Source | ◯ Wages/Income<br>◯ Pension<br>◯ Disability grant<br>◯ Child grant<br>◯ Other |
| HE10.1 Describe other | __________________________________ |

**Household member - 05**

| | |
|---|---|
| HE08. Occupation status | ◯ Formal employ (full-time)<br>◯ Formal employ (part-time)<br>◯ Self-employed<br>◯ Informal<br>◯ Piece jobs<br>◯ Student - FET/Tertiary<br>◯ In-school learner<br>◯ Pension<br>◯ Unemployed - Looking for work<br>◯ Unemployed - Not looking for work<br>◯ Pre-school going<br>◯ Other |
| HE08.1 Describe other | __________________________________ |

| | |
|---|---|
| HE09. Amount earned per month | ◯ R0<br>◯ R1-R999<br>◯ R1000-R4999<br>◯ R5000-R9999<br>◯ R10000+ |
| HE10. Source | ◯ Wages/Income<br>◯ Pension<br>◯ Disability grant<br>◯ Child grant<br>◯ Other |
| HE10.1 Describe other | __________________________________ |

**Household member - 06**

| | |
|---|---|
| HE08. Occupation status | ◯ Formal employ (full-time)<br>◯ Formal employ (part-time)<br>◯ Self-employed<br>◯ Informal<br>◯ Piece jobs<br>◯ Student - FET/Tertiary<br>◯ In-school learner<br>◯ Pension<br>◯ Unemployed - Looking for work<br>◯ Unemployed - Not looking for work<br>◯ Pre-school going<br>◯ Other |
| HE08.1 Describe other | __________________________________ |
| HE09. Amount earned per month | ◯ R0<br>◯ R1-R999<br>◯ R1000-R4999<br>◯ R5000-R9999<br>◯ R10000+ |
| HE10. Source | ◯ Wages/Income<br>◯ Pension<br>◯ Disability grant<br>◯ Child grant<br>◯ Other |
| HE10.1 Describe other | __________________________________ |

#### Household member - 07

| | |
|---|---|
| HE08. Occupation status | ◯ Formal employ (full-time)<br>◯ Formal employ (part-time)<br>◯ Self-employed<br>◯ Informal<br>◯ Piece jobs<br>◯ Student - FET/Tertiary<br>◯ In-school learner<br>◯ Pension<br>◯ Unemployed - Looking for work<br>◯ Unemployed - Not looking for work<br>◯ Pre-school going<br>◯ Other |
| HE08.1 Describe other | __________________________ |
| HE09. Amount earned per month | ◯ R0<br>◯ R1-R999<br>◯ R1000-R4999<br>◯ R5000-R9999<br>◯ R10000+ |
| HE10. Source | ◯ Wages/Income<br>◯ Pension<br>◯ Disability grant<br>◯ Child grant<br>◯ Other |
| HE10.1 Describe other | __________________________ |

#### Household member - 08

| | |
|---|---|
| HE08. Occupation status | ◯ Formal employ (full-time)<br>◯ Formal employ (part-time)<br>◯ Self-employed<br>◯ Informal<br>◯ Piece jobs<br>◯ Student - FET/Tertiary<br>◯ In-school learner<br>◯ Pension<br>◯ Unemployed - Looking for work<br>◯ Unemployed - Not looking for work<br>◯ Pre-school going<br>◯ Other |
| HE08.1 Describe other | __________________________ |
| HE09. Amount earned per month | ◯ R0<br>◯ R1-R999<br>◯ R1000-R4999<br>◯ R5000-R9999<br>◯ R10000+ |

HE10. Source

◯ Wages/Income
◯ Pension
◯ Disability grant
◯ Child grant
◯ Other

HE10.1 Describe other

__________________________________

#### Household member - 09

HE08. Occupation status

◯ Formal employ (full-time)
◯ Formal employ (part-time)
◯ Self-employed
◯ Informal
◯ Piece jobs
◯ Student - FET/Tertiary
◯ In-school learner
◯ Pension
◯ Unemployed - Looking for work
◯ Unemployed - Not looking for work
◯ Pre-school going
◯ Other

HE08.1 Describe other

__________________________________

HE09. Amount earned per month

◯ R0
◯ R1-R999
◯ R1000-R4999
◯ R5000-R9999
◯ R10000+

HE10. Source

◯ Wages/Income
◯ Pension
◯ Disability grant
◯ Child grant
◯ Other

HE10.1 Describe other

__________________________________

#### Household member - 10

HE08. Occupation status

◯ Formal employ (full-time)
◯ Formal employ (part-time)
◯ Self-employed
◯ Informal
◯ Piece jobs
◯ Student - FET/Tertiary
◯ In-school learner
◯ Pension
◯ Unemployed - Looking for work
◯ Unemployed - Not looking for work
◯ Pre-school going
◯ Other

HE08.1 Describe other

__________________________________

| | |
|---|---|
| HE09. Amount earned per month | ◯ R0<br>◯ R1-R999<br>◯ R1000-R4999<br>◯ R5000-R9999<br>◯ R10000+ |
| HE10. Source | ◯ Wages/Income<br>◯ Pension<br>◯ Disability grant<br>◯ Child grant<br>◯ Other |
| HE10.1 Describe other | __________________________________ |

**Health system and household Costs**

| | |
|---|---|
| HE11. Can you tell us if you/they had any other chronic illnesses before - or in addition to - TB for which they are receiving treatment? | ◯ HIV<br>◯ High blood pressure/Hypertension<br>◯ Diabetes (needing injection)<br>◯ Diabetes (needing tablets)<br>◯ Heart problems<br>◯ Cancer<br>◯ Other<br>◯ Not receiving treatment for any other chronic disease |
| HE11.1. Site of Cancer | __________________________________ |
| HE11.2. Describe other | __________________________________ |
| HE12. How many visits to the following services were made by the Index case in relation to symptoms of this TB(or TB/HIV) illness in the 6 months before the diagnosis? | ☐ 1. Government clinic or health centre<br>☐ 2. Private GP<br>☐ 3. Government hospital outpatient<br>☐ 4. Government hospital casualty<br>☐ 5. Traditional healer/sangoma/nyanga<br>☐ 6. Faith healer |
| HE12.1. Number of visits - 12.1. | __________________________________ |
| HE12.1. Number of visits - 12.2. | __________________________________ |
| HE12.1. Number of visits - 12.3. | __________________________________ |
| HE12.1. Number of visits - 12.4. | __________________________________ |
| HE12.1. Number of visits - 12.5. | __________________________________ |
| HE12.1. Number of visits - 12.6. | __________________________________ |

HE13. Did the person with TB or the Index case with TB had to be admitted to a hospital in the 6 months before this TB illness?

◯ Yes
◯ No

HE14.1. Hospital

__________________________________

HE14.2. Number of nights

__________________________________

HE15. Complaint/Diagnosis

__________________________________

HE16. From-

__________________________________

HE16. To-

__________________________________

Add another?

◯ Yes
◯ No

HE17.1. Hospital

__________________________________

HE17.2. Number of nights

__________________________________

HE18. Complaint/Diagnosis

__________________________________

HE19. From-

__________________________________

HE19. To-

__________________________________

Add another?

◯ Yes
◯ No

HE20.1. Hospital

__________________________________

HE20.2. Number of nights

__________________________________

HE21. Complaint/Diagnosis

__________________________________

HE22. From-

__________________________________

HE22. To-

__________________________________

HE23. How much did you pay for the services for each visit that the Index Case used?

- ☐ 1. Government clinic or health centre
- ☐ 2. Private GP
- ☐ 3. Government hospital outpatient
- ☐ 4. Government hospital casualty
- ☐ 5. Private hospital casualty
- ☐ 6. Traditional healer/sangoma/nyanga
- ☐ 7. Faith healer

---

HE24. ________ZAR

__________________________________

(ZAR. ( 0 if no cost. 9999 if unknown))

---

HE25. ________ZAR

__________________________________

(ZAR. ( 0 if no cost. 9999 if unknown))

---

HE26. ________ZAR

__________________________________

(ZAR. ( 0 if no cost. 9999 if unknown))

---

HE27. ________ZAR

__________________________________

(ZAR. ( 0 if no cost. 9999 if unknown))

---

HE28. ________ZAR

__________________________________

(ZAR. ( 0 if no cost. 9999 if unknown))

---

HE29. ________ZAR

__________________________________

(ZAR. ( 0 if no cost. 9999 if unknown))

---

HE30. ________ZAR

__________________________________

(ZAR. ( 0 if no cost. 9999 if unknown))

---

HE31. Do you have any kind of medical aid or other assistance with paying the health care costs?

- ◯ Yes - in above amount
- ◯ Yes - on top of above
- ◯ No

---

HE32. Medical Aid name

__________________________________

---

HE33. Medical Aid cost

__________________________________

(ZAR. ( 0 if no cost. 9999 if unknown))

---

HE34. Employer help

__________________________________

(ZAR. ( 0 if no cost. 9999 if unknown))

---

HE35. Other

__________________________________

(ZAR. ( 0 if no cost. 9999 if unknown))

| | |
|---|---|
| HE36. Has index case/or you taken off from work because of this illness? | ◯ Yes<br>◯ No<br>◯ Does not work |
| HE37. How many working days | __________________ |
| HE38. Has index case/you lost income because of this illness? | ◯ Yes<br>◯ No<br>◯ Does not have an income |
| HE39. | __________________<br>(ZAR. ( 0 if no cost. 9999 if unknown)) |
| HE40. How does the index case/you usually travel to the hospital to get care? | ☐ Private car<br>☐ Minibus or other taxi<br>☐ Bus<br>☐ Train<br>☐ Walk<br>☐ Other |
| HE40. Other | __________________ |
| HE41. What is the cost of one trip to get care | __________________<br>(ZAR. ( 0 if no cost. 9999 if unknown)) |
| HE42. How far does the index case have to travel to get care? | __________________<br>(km) |
| HE43. How much does the index spend on special treatments, supplements, vitamins or foods because of their sickness? | |
| HE43a. Fruit and Vegetables | ◯ Yes ◯ No |
| HE43b. Milk | ◯ Yes ◯ No |
| HE43c. Other drinks | ◯ Yes ◯ No |
| HE43d. Vitamins/Herbs | ◯ Yes ◯ No |
| HE43e. Meat | ◯ Yes ◯ No |
| HE43f. Eggs | ◯ Yes ◯ No |
| HE43g. Traditional Medicines | ◯ Yes ◯ No |
| HE43h. Other | ◯ Yes ◯ No |
| Specify | __________________ |

HE44. Overall amount spent on special treatments, supplements, vitamins or foods per month

________________ (ZAR)

HE45. Did anyone go with the index/you to the clinic or hospital?

○ Yes ○ No

HE46. How many adults

________________

HE46. Under 18 yrs

________________

HE47. How many times did that person/those people go with the index to the clinic or hospital?

________________ (times)

HE48. How much does the person(s) who accompanied the index earn per day?

HE49. Person 1

________________ (ZAR. ( 0 if no cost. 9999 if unknown))

HE50. Person 2

________________ (ZAR. ( 0 if no cost. 9999 if unknown))

HE51. Person 3

________________ (ZAR. ( 0 if no cost. 9999 if unknown))

HE52. Why did the index need to be accompanied to the clinic or hospital?

☐ Friendship/comfort/love/emotional support
☐ Security
☐ Index too sick to manage alone (short of breath, weak etc.)
☐ Index mentally incapable of managing alone
☐ Speak to health staff to explain condition & understand treatment
☐ Other (specify)
☐ Mostly went alone

HE52. Other

________________

HE53a. If index was admitted to hospital, how many people visited?

________________

HE53b. Number of visits

________________

HE53a. If index was admitted to hospital, how many people visited?

________________

HE53b. Number of visits

________________

HE53a. If index was admitted to hospital, how many people visited?

______________________________

HE53b. Number of visits

______________________________

#### Coping costs

HE54. Did the person with TB or someone in the household use their own money in the bank, or own money in a stokvel or elsewhere to pay for costs or expenses related to their TB?

- ◯ Yes
- ◯ No
- ◯ Don't know

HE55. How much

______________________________
(ZAR. ( 0 if no cost. 9999 if unknown))

HE56. Did the index case/family member borrow money for care or cover costs due to the illness ? (separate from own money in QHE54)

- ◯ Yes
- ◯ No
- ◯ Don't know

HE57. How much

______________________________
(ZAR)

HE58. When was the money borrowed?

______________________________

HE58. or,

- ☐ Don't know

HE59. From whom did you/they borrow money from?

- ☐ Family
- ☐ Neighbours/Friend
- ☐ Bank
- ☐ Stokvel
- ☐ Machonisa/loan shark
- ☐ Employer
- ☐ Didn't borrow
- ☐ Other

HE59. Other

______________________________

HE60. How much will you have to pay back and when?

Amount 1

- ☐

Specify amount 1

______________________________
(ZAR. ( 0 if no cost. 9999 if unknown))

Amount 1, after how long

______________________________

Amount 1: after how long

- ◯ weeks
- ◯ months

| | |
|---|---|
| Amount 2 | ☐ |
| Specify amount 2 | ______________________ (ZAR. ( 0 if no cost. 9999 if unknown)) |
| Amount 2, after how long | ______________________ |
| Amount 2: after how long | ◯ weeks<br>◯ months |
| Amount 3 | ☐ |
| Specify amount 3 | ______________________ (ZAR. ( 0 if no cost. 9999 if unknown)) |
| Amount 3, after how long | ______________________ |
| Amount 3: after how long | ◯ weeks<br>◯ months |
| Interest | ☐ |
| Specify interest rate | ______________________ (%) |
| Interest over how long | ______________________ |
| Interest over how long | ◯ weeks<br>◯ months |
| I am not expected to pay back the money | ☐ |
| Didn't borrow | ☐ |
| HE61. Have you paid the money back in full? | ◯ Yes paid in full<br>◯ Paid partly<br>◯ Only paid the interest<br>◯ Didn't borrow |
| HE62. Did the index case/family member have to sell an asset/livestock to get care? | ◯ Yes ◯ No |
| HE63. What asset/s or livestock? | ______________________ |
| HE64. Total value of all asset/s or livestock sold: | ______________________ (ZAR. ( 0 if no cost. 9999 if unknown)) |

HE65. How much did you actually get when you sold?

______________________ (ZAR)

HE66. Has the household had to reduce spending elsewhere?

◯ Yes ◯ No

HE67. What did the household reduce spending on the most?

☐ 1. Food
☐ 2. Petrol
☐ 3. Clothing
☐ 4. Airtime
☐ 5. Entertainment
☐ 6. School expenditure
☐ 7. Alcohol
☐ 8. Other

HE67. Other

______________________

HE68. What other things did they reduce spending on?

☐ 1. Food
☐ 2. Petrol
☐ 3. Clothing
☐ 4. Airtime
☐ 5. Entertainment
☐ 6. School expenditure
☐ 7. Alcohol
☐ 8. Other

HE69. Are other people involved with the care of the index case?

◯ Yes ◯ No

HE70. Did these people/person have to stop working to care for the index case?

◯ Yes ◯ No

HE70a Person 1

______________________ (weeks)

HE70b Person 2

______________________ (weeks)

HE70c Person 3

______________________ (weeks)

HE71. What is the monthly income earned by the people/person that looks after the index case?

HE71a: Person 1

◯ R0
◯ R1-R999
◯ R1000-R4999
◯ R5000-R9999
◯ +R10000

| | |
|---|---|
| HE71b: Person 2 | ◯ R0<br>◯ R1-R999<br>◯ R1000-R4999<br>◯ R5000-R9999<br>◯ +R10000 |
| HE71c: Person 3 | ◯ R0<br>◯ R1-R999<br>◯ R1000-R4999<br>◯ R5000-R9999<br>◯ +R10000 |
| HE72. Did any person lose or leave their job permanently to take of the index case? | ◯ Yes ◯ No |
| HE72a: Person 1 | ◯ R0<br>◯ R1-R999<br>◯ R1000-R4999<br>◯ R5000-R9999<br>◯ +R10000 |
| HE72b: Person 2 | ◯ R0<br>◯ R1-R999<br>◯ R1000-R4999<br>◯ R5000-R9999<br>◯ +R10000 |
| HE72c: Person 3 | ◯ R0<br>◯ R1-R999<br>◯ R1000-R4999<br>◯ R5000-R9999<br>◯ +R10000 |

#### Other illness and Additional cost of TB illness

| | |
|---|---|
| HE73. have there been any additional costs or health service visits for the index because of the other chronic illnesses mentioned above | ◯ Yes ◯ No |
| HE74. What are the other costs? | ☐ Clinic visits (per 6 months)<br>☐ Hospital outpatient visits (per 6 months)<br>☐ Hospital admission (nights in last 6 months)<br>☐ Tests or drugs<br>☐ Transport or food<br>☐ Other |
| HE74. Tests or drugs, cost: | ______________________<br>(ZAR. ( 0 if no cost. 9999 if unknown)) |
| HE74. Transport or food, cost: | ______________________<br>(ZAR. ( 0 if no cost. 9999 if unknown)) |
| HE74. Other | ______________________ |

HE75. On average, how much did the index spend per month on health care costs before the TB illness?

______________________
(ZAR. ( 0 if no cost. 9999 if unknown))

HE76. How much does the index spend per month towards health care costs now after the TB illness?

______________________
(ZAR. ( 0 if no cost. 9999 if unknown))

**If the index case has died**

HE77. How much did funeral arrangements cost your household?

______________________
(ZAR)

HE78. How much was paid with your household money?

______________________
(ZAR)

HE79. How much was paid by guests, friends or family?

______________________
(ZAR)

HE80. How much was paid by a burial society or funeral insurance?

______________________
(ZAR)

HE81. How much was borrowed by your household?

______________________
(ZAR)

HE82 If loan(s) has to be repaid, how much and by when?

◯ How much ◯ Interest rate

HE82. How much

______________________
(ZAR)

HE82. What interest rate

______________________
(%)

HE83. By when:

______________________

HE84. How much do you still owe?

______________________
(ZAR)

HE85. How many livestock did the household contribute to the funeral?
(Do not include any that were bought and included in payments under HE78, 79 or 80)

- ☐ Cow
- ☐ Goat
- ☐ Chicken
- ☐ Sheep
- ☐ Pigs
- ☐ Not applicable

HE85.1. Number of cows

______________________

HE85.2. Number of goats

________________________________

HE85.3. Number of chicken

________________________________

HE85.4. Number of sheep

________________________________

HE85.5. Number of pigs

________________________________

#### Overall impact

HE87. On a scale of 1 to 5, in which 1 is no impact and 5 is very serious impact, to what extent has the TB illness affected the family costs and income?

- ◯ 1-No impact
- ◯ 2-Little impact
- ◯ 3-Moderate impact
- ◯ 4-Serious impact
- ◯ 5-Very serious impact

HE88. Have any household members been sent to other households because yours could not cope since the index illness started?

◯ Yes ◯ No

HE89. How many members?

________________________________

HE90. Have any learners/ students in the household dropped out from learning since the index illness started?

◯ Yes ◯ No

Dropped for more than 3 months ☐

HE90. No.

________________________________

Dropped for more than 3 months ☐

HE90. No.

________________________________

#### Quality of Life

HE91. What do you expect will happen to the quality of life for the people in your household over the next year?

- ◯ Much better than 1 year ago
- ◯ Somewhat better than 1 year ago
- ◯ About the same
- ◯ Somewhat worse than one year ago
- ◯ Much worse than one year ago

HE92. In general the health of the index case is?

- ◯ Excellent
- ◯ Very good
- ◯ Good
- ◯ Fair
- ◯ Poor

| | |
|---|---|
| HE93. Compared to one year ago the index is? | ◯ Much better than 1 year ago<br>◯ Somewhat better than 1 year ago<br>◯ About the same<br>◯ Somewhat worse than one year ago<br>◯ Much worse than one year ago |
| HE94. Mobility | ◯ I have no problems in walking about<br>◯ I have slight problems in walking about<br>◯ I have moderate problems in walking about<br>◯ I have severe problems in walking about<br>◯ I am unable to walk about |
| HE95. Self Care | ◯ I have no problems washing or dressing myself<br>◯ I have slight problems washing or dressing myself<br>◯ I have moderate problems washing or dressing myself<br>◯ I have severe problems washing or dressing myself<br>◯ I am unable to wash or dress myself |
| HE96. Usual activities | ◯ I have no problems doing my usual activities<br>◯ I have slight problems doing my usual activities<br>◯ I have moderate problems doing my usual activities<br>◯ I have severe problems doing my usual activities<br>◯ I am unable to do my usual activities |
| HE97. Pain/discomfort | ◯ I have no pain or discomfort<br>◯ I have slight pain or discomfort<br>◯ I have moderate pain or discomfort<br>◯ I have severe pain or discomfort<br>◯ I have extreme pain or discomfort |
| HE98. Anxiety/depression | ◯ I am not anxious or depressed<br>◯ I am slightly anxious or depressed<br>◯ I am moderately anxious or depressed<br>◯ I am severely anxious or depressed<br>◯ I am extremely anxious or depressed |
| HE93. In the past two weeks | |
| HE93.1 Does the index case have enough energy for everyday life? | ☐ |
| HE93.1 Score | ◯ 1<br>◯ 2<br>◯ 3<br>◯ 4<br>◯ 5 |
| HE93.2 Does the index case have enough money to meet his/her needs? | ☐ |
| HE93.2 Score | ◯ 1<br>◯ 2<br>◯ 3<br>◯ 4<br>◯ 5 |
| HE93.3 Does the index case feel accepted by the people that he/she knows? | ☐ |

| | |
|---|---|
| HE93.3 Score | ◯ 1<br>◯ 2<br>◯ 3<br>◯ 4<br>◯ 5 |
| HE93.4 Does the index case feel life is meaningful? | ☐ |
| HE93.4 Score | ◯ 1<br>◯ 2<br>◯ 3<br>◯ 4<br>◯ 5 |
| HE93.5 How much does the index case enjoy life? | ☐ |
| HE93.5 Score | ◯ 1<br>◯ 2<br>◯ 3<br>◯ 4<br>◯ 5 |
| HE100. How satisfied is the index case with his/her quality of sleep? | ◯ Very good<br>◯ Good<br>◯ Neither good nor bad<br>◯ Poor<br>◯ Very poor |
| HE101. To what extent is the index case troubled by people knowing that he/she has TB ?<br><br>(rate out of 5, with 5 being very troubled and 1 least troubled)<br>(If a child: to what extent will it create trouble for the child if people know that he/she has TB?) | ◯ 1<br>◯ 2<br>◯ 3<br>◯ 4<br>◯ 5 |
| HE102. To what extent is the index case troubled by people knowing that he/she has HIV? [if relevant]<br><br>(rate out of 5, with 5 being very troubled and 1 least troubled)<br>(If a child: to what extent will it create trouble for the child if people know that he/she has TB?) | ◯ 1<br>◯ 2<br>◯ 3<br>◯ 4<br>◯ 5 |

#### Form Completion

| | |
|---|---|
| HE103. Date form HE completed | __________________________________ |
| HE104. Name of person completing this form | __________________________________ |

#### Audit Trail

Name of Data Entry Person

__________________________________

Date of Data Entry

__________________________________
(Kindly change the date every time a change is made to the data)

Form Version: 1.1 dated 14 June 2017

__________________________________

### Health Economics Form HE_M15

| | |
|---|---|
| Form completed at: | ◯ Month 15 |
| i. Start Time | __________________ |

#### Baseline

| | |
|---|---|
| ii. Date of Interview | __________________ |
| iii. Status of Index Case | ◯ Alive<br>◯ Deceased |
| iii.i. Date decesaed | __________________ |
| iv. Date of TB diagnosis | __________________ |
| v. Who is the main person providing information for this questionnaire? | ◯ Index case<br>◯ Other |
| v.i. Relation to Index case | ◯ Wife/Mother<br>◯ Husband/Father<br>◯ Extended family<br>◯ Son/Daughter<br>◯ Other |
| v.i.i. Describe other | __________________ |

#### Household Socio-economic Status

| | |
|---|---|
| HE01. How many people in the household receive an income?<br>(including pensions and grants) | __________________ |
| HE02. Who is the primary income earner in the household? | ◯ Index<br>◯ Wife/Mother<br>◯ Husband/Father<br>◯ Extended family<br>◯ Son/Daughter<br>◯ Other |
| HE02.1 Describe other | __________________ |

HE03. Does your household have any of the following and how many?

- ☐ Refrigerator
- ☐ Stove
- ☐ Television
- ☐ Car
- ☐ Radio
- ☐ Phone-landline
- ☐ Internet
- ☐ Microwave
- ☐ Cellphone
- ☐ Smartphone
- ☐ Computer
- ☐ Washing machine
- ☐ Satellite dish/DSTV

HE03.01. Number - Refrigirator ______________________________

HE03.02. Number - Stove ______________________________

HE03.03. Number - Television ______________________________

HE03.04. Number - Car ______________________________

HE03.05. Number - Radio ______________________________

HE03.06. Number - Phone-landline ______________________________

HE03.07. Number - Internet ______________________________

HE03.08. Number - Microwave ______________________________

HE03.09. Number - Cellphone ______________________________

HE03.10. Number - Smartphone ______________________________

HE03.11. Number - Computer ______________________________

HE03.12. Number - Washing machine ______________________________

HE03.13. Number - Satellite dish/DSTV ______________________________

| | |
|---|---|
| HE04. Household food production | ☐ Livestock<br>☐ Mielies/Sorghum<br>☐ Vegetables<br>☐ Poultry<br>☐ Other |
| HE04.1. Describe other | __________________________ |
| HE05. Do you have additional income sent by household members not living with you? | ○ Yes<br>○ No |
| HE05.1. How many? | __________________________ |
| HE06. Amount.R- | __________________________ |
| HE07. Household members | __________________________ |

#### Household member - 01

| | |
|---|---|
| HE08. Occupation status | ○ Formal employ (full-time)<br>○ Formal employ (part-time)<br>○ Self-employed<br>○ Informal<br>○ Piece jobs<br>○ Student - FET/Tertiary<br>○ In-school learner<br>○ Pension<br>○ Unemployed - Looking for work<br>○ Unemployed - Not looking for work<br>○ Pre-school going<br>○ Other |
| HE08.1 Describe other | __________________________ |
| HE09. Amount earned per month | ○ R0<br>○ R1-R999<br>○ R1000-R4999<br>○ R5000-R9999<br>○ R10000+ |
| HE10. Source | ○ Wages/Income<br>○ Pension<br>○ Disability grant<br>○ Child grant<br>○ Other |
| HE10.1 Describe other | __________________________ |

#### Household member - 02

HE08. Occupation status

- ◯ Formal employ (full-time)
- ◯ Formal employ (part-time)
- ◯ Self-employed
- ◯ Informal
- ◯ Piece jobs
- ◯ Student - FET/Tertiary
- ◯ In-school learner
- ◯ Pension
- ◯ Unemployed - Looking for work
- ◯ Unemployed - Not looking for work
- ◯ Pre-school going
- ◯ Other

HE08.1 Describe other

__________________________________

HE09. Amount earned per month

- ◯ R0
- ◯ R1-R999
- ◯ R1000-R4999
- ◯ R5000-R9999
- ◯ R10000+

HE10. Source

- ◯ Wages/Income
- ◯ Pension
- ◯ Disability grant
- ◯ Child grant
- ◯ Other

HE10.1 Describe other

__________________________________

#### Household member - 03

HE08. Occupation status

- ◯ Formal employ (full-time)
- ◯ Formal employ (part-time)
- ◯ Self-employed
- ◯ Informal
- ◯ Piece jobs
- ◯ Student - FET/Tertiary
- ◯ In-school learner
- ◯ Pension
- ◯ Unemployed - Looking for work
- ◯ Unemployed - Not looking for work
- ◯ Pre-school going
- ◯ Other

HE08.1 Describe other

__________________________________

HE09. Amount earned per month

- ◯ R0
- ◯ R1-R999
- ◯ R1000-R4999
- ◯ R5000-R9999
- ◯ R10000+

HE10. Source

- ◯ Wages/Income
- ◯ Pension
- ◯ Disability grant
- ◯ Child grant
- ◯ Other

HE10.1 Describe other

__________________________________

#### Household member - 04

HE08. Occupation status

- ◯ Formal employ (full-time)
- ◯ Formal employ (part-time)
- ◯ Self-employed
- ◯ Informal
- ◯ Piece jobs
- ◯ Student - FET/Tertiary
- ◯ In-school learner
- ◯ Pension
- ◯ Unemployed - Looking for work
- ◯ Unemployed - Not looking for work
- ◯ Pre-school going
- ◯ Other

HE08.1 Describe other

__________________________________

HE09. Amount earned per month

- ◯ R0
- ◯ R1-R999
- ◯ R1000-R4999
- ◯ R5000-R9999
- ◯ R10000+

HE10. Source

- ◯ Wages/Income
- ◯ Pension
- ◯ Disability grant
- ◯ Child grant
- ◯ Other

HE10.1 Describe other

__________________________________

#### Household member - 05

HE08. Occupation status

- ◯ Formal employ (full-time)
- ◯ Formal employ (part-time)
- ◯ Self-employed
- ◯ Informal
- ◯ Piece jobs
- ◯ Student - FET/Tertiary
- ◯ In-school learner
- ◯ Pension
- ◯ Unemployed - Looking for work
- ◯ Unemployed - Not looking for work
- ◯ Pre-school going
- ◯ Other

HE08.1 Describe other

__________________________________

| | |
|---|---|
| HE09. Amount earned per month | ◯ R0<br>◯ R1-R999<br>◯ R1000-R4999<br>◯ R5000-R9999<br>◯ R10000+ |
| HE10. Source | ◯ Wages/Income<br>◯ Pension<br>◯ Disability grant<br>◯ Child grant<br>◯ Other |
| HE10.1 Describe other | __________________________________ |

**Household member - 06**

| | |
|---|---|
| HE08. Occupation status | ◯ Formal employ (full-time)<br>◯ Formal employ (part-time)<br>◯ Self-employed<br>◯ Informal<br>◯ Piece jobs<br>◯ Student - FET/Tertiary<br>◯ In-school learner<br>◯ Pension<br>◯ Unemployed - Looking for work<br>◯ Unemployed - Not looking for work<br>◯ Pre-school going<br>◯ Other |
| HE08.1 Describe other | __________________________________ |
| HE09. Amount earned per month | ◯ R0<br>◯ R1-R999<br>◯ R1000-R4999<br>◯ R5000-R9999<br>◯ R10000+ |
| HE10. Source | ◯ Wages/Income<br>◯ Pension<br>◯ Disability grant<br>◯ Child grant<br>◯ Other |
| HE10.1 Describe other | __________________________________ |

#### Household member - 07

HE08. Occupation status

- ◯ Formal employ (full-time)
- ◯ Formal employ (part-time)
- ◯ Self-employed
- ◯ Informal
- ◯ Piece jobs
- ◯ Student - FET/Tertiary
- ◯ In-school learner
- ◯ Pension
- ◯ Unemployed - Looking for work
- ◯ Unemployed - Not looking for work
- ◯ Pre-school going
- ◯ Other

HE08.1 Describe other

__________________________________

HE09. Amount earned per month

- ◯ R0
- ◯ R1-R999
- ◯ R1000-R4999
- ◯ R5000-R9999
- ◯ R10000+

HE10. Source

- ◯ Wages/Income
- ◯ Pension
- ◯ Disability grant
- ◯ Child grant
- ◯ Other

HE10.1 Describe other

__________________________________

#### Household member - 08

HE08. Occupation status

- ◯ Formal employ (full-time)
- ◯ Formal employ (part-time)
- ◯ Self-employed
- ◯ Informal
- ◯ Piece jobs
- ◯ Student - FET/Tertiary
- ◯ In-school learner
- ◯ Pension
- ◯ Unemployed - Looking for work
- ◯ Unemployed - Not looking for work
- ◯ Pre-school going
- ◯ Other

HE08.1 Describe other

__________________________________

HE09. Amount earned per month

- ◯ R0
- ◯ R1-R999
- ◯ R1000-R4999
- ◯ R5000-R9999
- ◯ R10000+

| | |
|---|---|
| HE10. Source | ◯ Wages/Income<br>◯ Pension<br>◯ Disability grant<br>◯ Child grant<br>◯ Other |
| HE10.1 Describe other | __________________________ |

#### Household member - 09

| | |
|---|---|
| HE08. Occupation status | ◯ Formal employ (full-time)<br>◯ Formal employ (part-time)<br>◯ Self-employed<br>◯ Informal<br>◯ Piece jobs<br>◯ Student - FET/Tertiary<br>◯ In-school learner<br>◯ Pension<br>◯ Unemployed - Looking for work<br>◯ Unemployed - Not looking for work<br>◯ Pre-school going<br>◯ Other |
| HE08.1 Describe other | __________________________ |
| HE09. Amount earned per month | ◯ R0<br>◯ R1-R999<br>◯ R1000-R4999<br>◯ R5000-R9999<br>◯ R10000+ |
| HE10. Source | ◯ Wages/Income<br>◯ Pension<br>◯ Disability grant<br>◯ Child grant<br>◯ Other |
| HE10.1 Describe other | __________________________ |

#### Household member - 10

| | |
|---|---|
| HE08. Occupation status | ◯ Formal employ (full-time)<br>◯ Formal employ (part-time)<br>◯ Self-employed<br>◯ Informal<br>◯ Piece jobs<br>◯ Student - FET/Tertiary<br>◯ In-school learner<br>◯ Pension<br>◯ Unemployed - Looking for work<br>◯ Unemployed - Not looking for work<br>◯ Pre-school going<br>◯ Other |
| HE08.1 Describe other | __________________________ |

| | |
|---|---|
| HE09. Amount earned per month | ◯ R0<br>◯ R1-R999<br>◯ R1000-R4999<br>◯ R5000-R9999<br>◯ R10000+ |
| HE10. Source | ◯ Wages/Income<br>◯ Pension<br>◯ Disability grant<br>◯ Child grant<br>◯ Other |
| HE10.1 Describe other | ______________________ |

**Health system and Household Costs**

| | |
|---|---|
| HE11. Can you tell us if you/they had any other chronic illnesses before - or in addition to - TB for which they are receiving treatment? | ◯ HIV<br>◯ High blood pressure/Hypertension<br>◯ Diabetes (needing injection)<br>◯ Diabetes (needing tablets)<br>◯ Heart problems<br>◯ Cancer<br>◯ Other<br>◯ Not receiving treatment for any other chronic disease |
| HE11.1. Site of Cancer | ______________________ |
| HE11.2. Describe other | ______________________ |
| HE12. How many visits to the following services were made by the Index case in relation to symptoms of this TB(or TB/HIV) illness in the 6 months before the diagnosis? | ☐ 1. Government clinic or health centre<br>☐ 2. Private GP<br>☐ 3. Government hospital outpatient<br>☐ 4. Government hospital casualty<br>☐ 5. Traditional healer/sangoma/nyanga<br>☐ 6. Faith healer |
| HE12.1. Number of visits - 12.1. | ______________________ |
| HE12.1. Number of visits - 12.2. | ______________________ |
| HE12.1. Number of visits - 12.3. | ______________________ |
| HE12.1. Number of visits - 12.4. | ______________________ |
| HE12.1. Number of visits - 12.5. | ______________________ |
| HE12.1. Number of visits - 12.6. | ______________________ |

| | |
|---|---|
| HE13. Did the person with TB or the Index case with TB had to be admitted to a hospital in the 6 months before this TB illness? | ◯ Yes<br>◯ No |
| HE14.1. Hospital | __________________________ |
| HE14.2. Number of nights | __________________________ |
| HE15. Complaint/Diagnosis | __________________________ |
| HE16. From- | __________________________ |
| HE16. To- | __________________________ |
| Add another? | ◯ Yes<br>◯ No |
| HE17.1. Hospital | __________________________ |
| HE17.2. Number of nights | __________________________ |
| HE18. Complaint/Diagnosis | __________________________ |
| HE19. From- | __________________________ |
| HE19. To- | __________________________ |
| Add another? | ◯ Yes<br>◯ No |
| HE20.1. Hospital | __________________________ |
| HE20.2. Number of nights | __________________________ |
| HE21. Complaint/Diagnosis | __________________________ |
| HE22. From- | __________________________ |
| HE22. To- | __________________________ |

HE23. How much did you pay for the services for each visit that the Index Case used?

- [ ] 1. Government clinic or health centre
- [ ] 2. Private GP
- [ ] 3. Government hospital outpatient
- [ ] 4. Government hospital casualty
- [ ] 5. Private hospital casualty
- [ ] 6. Traditional healer/sangoma/nyanga
- [ ] 7. Faith healer

HE24. ________ZAR

________________________________
(ZAR. ( 0 if no cost. 9999 if unknown))

HE25. ________ZAR

________________________________
(ZAR. ( 0 if no cost. 9999 if unknown))

HE26. ________ZAR

________________________________
(ZAR. ( 0 if no cost. 9999 if unknown))

HE27. ________ZAR

________________________________
(ZAR. ( 0 if no cost. 9999 if unknown))

HE28. ________ZAR

________________________________
(ZAR. ( 0 if no cost. 9999 if unknown))

HE29. ________ZAR

________________________________
(ZAR. ( 0 if no cost. 9999 if unknown))

HE30. ________ZAR

________________________________
(ZAR. ( 0 if no cost. 9999 if unknown))

HE31. Do you have any kind of medical aid or other assistance with paying the health care costs?

○ Yes - in above amount
○ Yes - on top of above
○ No

HE32. Medical Aid name

________________________________

HE33. Medical Aid cost

________________________________
(ZAR. ( 0 if no cost. 9999 if unknown))

HE34. Employer help

________________________________
(ZAR. ( 0 if no cost. 9999 if unknown))

HE35. Other

________________________________
(ZAR. ( 0 if no cost. 9999 if unknown))

| | |
|---|---|
| HE36. Has index case/or you taken off from work because of this illness? | ◯ Yes<br>◯ No<br>◯ Does not work |
| HE37. How many working days | ______________________ |
| HE38. Has index case/you lost income because of this illness? | ◯ Yes<br>◯ No<br>◯ Does not have an income |
| HE39. | ______________________<br>(ZAR. ( 0 if no cost. 9999 if unknown)) |
| HE40. How does the index case/you usually travel to the hospital to get care? | ☐ Private car<br>☐ Minibus or other taxi<br>☐ Bus<br>☐ Train<br>☐ Walk<br>☐ Other |
| HE40. Other | ______________________ |
| HE41. What is the cost of one trip to get care | ______________________<br>(ZAR. ( 0 if no cost. 9999 if unknown)) |
| HE42. How far does the index case have to travel to get care? | ______________________<br>(km) |
| HE43. How much does the index spend on special treatments, supplements, vitamins or foods because of their sickness? | |
| HE43a. Fruit and Vegetables | ◯ Yes ◯ No |
| HE43b. Milk | ◯ Yes ◯ No |
| HE43c. Other drinks | ◯ Yes ◯ No |
| HE43d. Vitamins/Herbs | ◯ Yes ◯ No |
| HE43e. Meat | ◯ Yes ◯ No |
| HE43f. Eggs | ◯ Yes ◯ No |
| HE43g. Traditional Medicines | ◯ Yes ◯ No |
| HE43h. Other | ◯ Yes ◯ No |
| Specify | ______________________ |

| | |
|---|---|
| HE44. Overall amount spent on special treatments, supplements, vitamins or foods per month | ______________________ (ZAR) |
| HE45. Did anyone go with the index/you to the clinic or hospital? | ◯ Yes ◯ No |
| HE46. How many adults | ______________________ |
| HE46. Under 18 yrs | ______________________ |
| HE47. How many times did that person/those people go with the index to the clinic or hospital? | ______________________ (times) |
| HE48. How much does the person(s) who accompanied the index earn per day? | |
| HE49. Person 1 | ______________________ (ZAR. ( 0 if no cost. 9999 if unknown)) |
| HE50. Person 2 | ______________________ (ZAR. ( 0 if no cost. 9999 if unknown)) |
| HE51. Person 3 | ______________________ (ZAR. ( 0 if no cost. 9999 if unknown)) |
| HE52. Why did the index need to be accompanied to the clinic or hospital? | ☐ Friendship/comfort/love/emotional support<br>☐ Security<br>☐ Index too sick to manage alone (short of breath, weak etc.)<br>☐ Index mentally incapable of managing alone<br>☐ Speak to health staff to explain condition & understand treatment<br>☐ Other (specify)<br>☐ Mostly went alone |
| HE52. Other | ______________________ |
| HE53a. If index was admitted to hospital, how many people visited? | ______________________ |
| HE53b. Number of visits | ______________________ |
| HE53a. If index was admitted to hospital, how many people visited? | ______________________ |
| HE53b. Number of visits | ______________________ |

HE53a. If index was admitted to hospital, how many people visited?

______________________________

HE53b. Number of visits

______________________________

#### Coping costs

HE54. Did the person with TB or someone in the household use their own money in the bank, or own money in a stokvel or elsewhere to pay for costs or expenses related to their TB?

○ Yes
○ No
○ Don't know

HE55. How much

______________________________
(ZAR. ( 0 if no cost. 9999 if unknown))

HE56. Did the index case/family member borrow money for care or cover costs due to the illness ? (separate from own money in QHE54)

○ Yes
○ No
○ Don't know

HE57. How much

______________________________
(ZAR)

HE58. When was the money borrowed?

______________________________

HE58. or,

☐ Don't know

HE59. From whom did you/they borrow money from?

☐ Family
☐ Neighbours/Friend
☐ Bank
☐ Stokvel
☐ Machonisa/loan shark
☐ Employer
☐ Didn't borrow
☐ Other

HE59. Other

______________________________

HE60. How much will you have to pay back and when?

Amount 1

☐

Specify amount 1

______________________________
(ZAR. ( 0 if no cost. 9999 if unknown))

Amount 1, after how long

______________________________

Amount 1: after how long

○ weeks
○ months

| | |
|---|---|
| Amount 2 | ☐ |
| Specify amount 2 | ______________________ (ZAR. ( 0 if no cost. 9999 if unknown)) |
| Amount 2, after how long | ______________________ |
| Amount 2: after how long | ◯ weeks<br>◯ months |
| Amount 3 | ☐ |
| Specify amount 3 | ______________________ (ZAR. ( 0 if no cost. 9999 if unknown)) |
| Amount 3, after how long | ______________________ |
| Amount 3: after how long | ◯ weeks<br>◯ months |
| Interest | ☐ |
| Specify interest rate | ______________________ (%) |
| Interest over how long | ______________________ |
| Interest over how long | ◯ weeks<br>◯ months |
| I am not expected to pay back the money | ☐ |
| Didn't borrow | ☐ |
| HE61. Have you paid the money back in full? | ◯ Yes paid in full<br>◯ Paid partly<br>◯ Only paid the interest<br>◯ Didn't borrow |
| HE62. Did the index case/family member have to sell an asset/livestock to get care? | ◯ Yes ◯ No |
| HE63. What asset/s or livestock? | ______________________ |
| HE64. Total value of all asset/s or livestock sold: | ______________________ (ZAR. ( 0 if no cost. 9999 if unknown)) |

HE65. How much did you actually get when you sold?

______________________
(ZAR)

HE66. Has the household had to reduce spending elsewhere?

◯ Yes ◯ No

HE67. What did the household reduce spending on the most?

☐ 1. Food
☐ 2. Petrol
☐ 3. Clothing
☐ 4. Airtime
☐ 5. Entertainment
☐ 6. School expenditure
☐ 7. Alcohol
☐ 8. Other

HE67. Other

______________________

HE68. What other things did they reduce spending on?

☐ 1. Food
☐ 2. Petrol
☐ 3. Clothing
☐ 4. Airtime
☐ 5. Entertainment
☐ 6. School expenditure
☐ 7. Alcohol
☐ 8. Other

HE69. Are other people involved with the care of the index case?

◯ Yes ◯ No

HE70. Did these people/person have to stop working to care for the index case?

◯ Yes ◯ No

HE70a Person 1

______________________
(weeks)

HE70b Person 2

______________________
(weeks)

HE70c Person 3

______________________
(weeks)

HE71. What is the monthly income earned by the people/person that looks after the index case?

HE71a: Person 1

◯ R0
◯ R1-R999
◯ R1000-R4999
◯ R5000-R9999
◯ +R10000

| | |
|---|---|
| HE71b: Person 2 | ◯ R0<br>◯ R1-R999<br>◯ R1000-R4999<br>◯ R5000-R9999<br>◯ +R10000 |
| HE71c: Person 3 | ◯ R0<br>◯ R1-R999<br>◯ R1000-R4999<br>◯ R5000-R9999<br>◯ +R10000 |
| HE72. Did any person lose or leave their job permanently to take of the index case? | ◯ Yes ◯ No |
| HE72a: Person 1 | ◯ R0<br>◯ R1-R999<br>◯ R1000-R4999<br>◯ R5000-R9999<br>◯ +R10000 |
| HE72b: Person 2 | ◯ R0<br>◯ R1-R999<br>◯ R1000-R4999<br>◯ R5000-R9999<br>◯ +R10000 |
| HE72c: Person 3 | ◯ R0<br>◯ R1-R999<br>◯ R1000-R4999<br>◯ R5000-R9999<br>◯ +R10000 |

#### Other illness and Additional cost of TB illness

| | |
|---|---|
| HE73. have there been any additional costs or health service visits for the index because of the other chronic illnesses mentioned above | ◯ Yes ◯ No |
| HE74. What are the other costs? | ☐ Clinic visits (per 6 months)<br>☐ Hospital outpatient visits (per 6 months)<br>☐ Hospital admission (nights in last 6 months)<br>☐ Tests or drugs<br>☐ Transport or food<br>☐ Other |
| HE74. Tests or drugs, cost: | ______________________<br>(ZAR. ( 0 if no cost. 9999 if unknown)) |
| HE74. Transport or food, cost: | ______________________<br>(ZAR. ( 0 if no cost. 9999 if unknown)) |
| HE74. Other | ______________________ |

HE75. On average, how much did the index spend per month on health care costs before the TB illness?

________________________
(ZAR. ( 0 if no cost. 9999 if unknown))

HE76. How much does the index spend per month towards health care costs now after the TB illness?

________________________
(ZAR. ( 0 if no cost. 9999 if unknown))

#### If the index case has died

HE77. How much did funeral arrangements cost your household?

________________________
(ZAR)

HE78. How much was paid with your household money?

________________________
(ZAR)

HE79. How much was paid by guests, friends or family?

________________________
(ZAR)

HE80. How much was paid by a burial society or funeral insurance?

________________________
(ZAR)

HE81. How much was borrowed by your household?

________________________
(ZAR)

HE82 If loan(s) has to be repaid, how much and by when?

◯ How much ◯ Interest rate

HE82. How much

________________________
(ZAR)

HE82. What interest rate

________________________
(%)

HE83. By when:

________________________

HE84. How much do you still owe?

________________________
(ZAR)

HE85. How many livestock did the household contribute to the funeral?
(Do not include any that were bought and included in payments under HE78, 79 or 80)

☐ Cow
☐ Goat
☐ Chicken
☐ Sheep
☐ Pigs
☐ Not applicable

HE85.1. Number of cows

________________________

HE85.2. Number of goats ________________________

HE85.3. Number of chicken ________________________

HE85.4. Number of sheep ________________________

HE85.5. Number of pigs ________________________

#### Overall impact

HE87. On a scale of 1 to 5, in which 1 is no impact and 5 is very serious impact, to what extent has the TB illness affected the family costs and income?

- ◯ 1-No impact
- ◯ 2-Little impact
- ◯ 3-Moderate impact
- ◯ 4-Serious impact
- ◯ 5-Very serious impact

HE88. Have any household members been sent to other households because yours could not cope since the index illness started?

◯ Yes ◯ No

HE89. How many members? ________________________

HE90. Have any learners/ students in the household dropped out from learning since the index illness started?

◯ Yes ◯ No

Dropped for more than 3 months ☐

HE90. No. ________________________

Dropped for more than 3 months ☐

HE90. No. ________________________

#### Quality of Life

HE91. What do you expect will happen to the quality of life for the people in your household over the next year?

- ◯ Much better than 1 year ago
- ◯ Somewhat better than 1 year ago
- ◯ About the same
- ◯ Somewhat worse than one year ago
- ◯ Much worse than one year ago

HE92. In general the health of the index case is?

- ◯ Excellent
- ◯ Very good
- ◯ Good
- ◯ Fair
- ◯ Poor

| | |
|---|---|
| HE93. Compared to one year ago the index is? | ◯ Much better than 1 year ago<br>◯ Somewhat better than 1 year ago<br>◯ About the same<br>◯ Somewhat worse than one year ago<br>◯ Much worse than one year ago |
| HE94. Mobility | ◯ I have no problems in walking about<br>◯ I have slight problems in walking about<br>◯ I have moderate problems in walking about<br>◯ I have severe problems in walking about<br>◯ I am unable to walk about |
| HE95. Self Care | ◯ I have no problems washing or dressing myself<br>◯ I have slight problems washing or dressing myself<br>◯ I have moderate problems washing or dressing myself<br>◯ I have severe problems washing or dressing myself<br>◯ I am unable to wash or dress myself |
| HE96. Usual activities | ◯ I have no problems doing my usual activities<br>◯ I have slight problems doing my usual activities<br>◯ I have moderate problems doing my usual activities<br>◯ I have severe problems doing my usual activities<br>◯ I am unable to do my usual activities |
| HE97. Pain/discomfort | ◯ I have no pain or discomfort<br>◯ I have slight pain or discomfort<br>◯ I have moderate pain or discomfort<br>◯ I have severe pain or discomfort<br>◯ I have extreme pain or discomfort |
| HE98. Anxiety/depression | ◯ I am not anxious or depressed<br>◯ I am slightly anxious or depressed<br>◯ I am moderately anxious or depressed<br>◯ I am severely anxious or depressed<br>◯ I am extremely anxious or depressed |
| HE93. In the past two weeks | |
| HE93.1 Does the index case have enough energy for everyday life? | ☐ |
| HE93.1 Score | ◯ 1<br>◯ 2<br>◯ 3<br>◯ 4<br>◯ 5 |
| HE93.2 Does the index case have enough money to meet his/her needs? | ☐ |
| HE93.2 Score | ◯ 1<br>◯ 2<br>◯ 3<br>◯ 4<br>◯ 5 |
| HE93.3 Does the index case feel accepted by the people that he/she knows? | ☐ |

| | |
|---|---|
| HE93.3 Score | ◯ 1<br>◯ 2<br>◯ 3<br>◯ 4<br>◯ 5 |
| HE93.4 Does the index case feel life is meaningful? | ☐ |
| HE93.4 Score | ◯ 1<br>◯ 2<br>◯ 3<br>◯ 4<br>◯ 5 |
| HE93.5 How much does the index case enjoy life? | ☐ |
| HE93.5 Score | ◯ 1<br>◯ 2<br>◯ 3<br>◯ 4<br>◯ 5 |
| HE100. How satisfied is the index case with his/her quality of sleep? | ◯ Very good<br>◯ Good<br>◯ Neither good nor bad<br>◯ Poor<br>◯ Very poor |
| HE101. To what extent is the index case troubled by people knowing that he/she has TB ?<br><br>(rate out of 5, with 5 being very troubled and 1 least troubled)<br>(If a child: to what extent will it create trouble for the child if people know that he/she has TB?) | ◯ 1<br>◯ 2<br>◯ 3<br>◯ 4<br>◯ 5 |
| HE102. To what extent is the index case troubled by people knowing that he/she has HIV? [if relevant]<br><br>(rate out of 5, with 5 being very troubled and 1 least troubled)<br>(If a child: to what extent will it create trouble for the child if people know that he/she has TB?) | ◯ 1<br>◯ 2<br>◯ 3<br>◯ 4<br>◯ 5 |

#### Form Completion

| | |
|---|---|
| HE103. Date form HE completed | ____________________ |
| HE104. Name of person completing this form | ____________________ |

#### Audit Trail

Name of Data Entry Person

______________________________

Date of Data Entry

______________________________
(Kindly change the date every time a change is made to the data)

Form Version: 1.1 dated 14 June 2017

______________________________

### Costing Of Intervention

HH Study ID: ______________________

Visit Date: ______________________

Departure time from office/previous household ______________________

Return time to office/ arrival at next HH ______________________

Start time of HH visit (on site) ______________________

Departure time from HH ______________________

Nearest Clinic/Health Centre to HH (name): ______________________

Nearest Clinic GPS coordinates ______________________

Household GPS coordinates ______________________

Type of visit:
- ◯ 1st
- ◯ Results Feedback
- ◯ Start treatment
- ◯ Other

No one at home so visit cancelled/rescheduled? ◯ Yes ◯ No

Repeat visit due to absent HH member at last visit? ◯ Yes ◯ No

#### Personnel

Professional nurses

Grade and rank ______________________

Total Time for visit (Mins) for HH* ______________________
(* Including preparation, waiting; Travel to and from HH; General introduction, find HH members; Patient records; Informed consent; CRF data collection; Intervention tests etc)

Total time ______________________

| | |
|---|---|
| Enrolled nurse | |
| Grade and rank | ______________________ |
| Total Time for visit (Mins) for HH* | ______________________<br>(* Including preparation, waiting; Travel to and from HH; General introduction, find HH members; Patient records; Informed consent; CRF data collection; Intervention tests etc) |
| Total time | ______________________ |
| Counsellor | |
| Grade and rank | ______________________ |
| Total Time for visit (Mins) for HH* | ______________________<br>(* Including preparation, waiting; Travel to and from HH; General introduction, find HH members; Patient records; Informed consent; CRF data collection; Intervention tests etc) |
| Total time | ______________________ |
| Other (please state) | |
| Grade and rank | ______________________ |
| Total Time for visit (Mins) for HH* | ______________________<br>(* Including preparation, waiting; Travel to and from HH; General introduction, find HH members; Patient records; Informed consent; CRF data collection; Intervention tests etc) |
| Total time | ______________________ |
| Other (please state) | |
| Grade and rank | ______________________ |
| Total Time for visit (Mins) for HH* | ______________________<br>(* Including preparation, waiting; Travel to and from HH; General introduction, find HH members; Patient records; Informed consent; CRF data collection; Intervention tests etc) |

Total time ______________________

#### Tests at household

Sputum for Xpert

Number Done ______________________

Sputum for culture

Number Done ______________________

HIV test

Number Done ______________________

TST

Number Done ______________________

Isoniazid

Number Done ______________________

Vit B12

Number Done ______________________

#### Transport

Vehicle type
- ◯ Car
- ◯ 4x4
- ◯ bakkie
- ◯ minibus
- ◯ other

Distance to HH from nearest clinic/ health centre (not Study Office) ______________________

#### Other costs

Calls

Number ______________________

Items ______________________

Cost per item ______________________

Stationary

Number ______________________

Items ______________________

Cost per item ______________________

Consumables

Number ______________________

Items ______________________

Cost per item ______________________

Airtime

Number ______________________

Items ______________________

Cost per item ______________________

Other

Number ______________________

Items ______________________

Cost per item ______________________

#### Home Visit Log for staff time direct observation of time and motion

Visit Date ______________________

HH Visit Date ______________________

Visit Start time ______________________

Visit End Time ______________________

Number of HH members screened > 15yrs ______________________

Number of HH members screened < 15yrs ______________________

DID THE PROVIDER AGREE TO PARTICIPATE? ◯ Yes ◯ No

Add Obs. 1 ◯ Yes ◯ No

Obs. 1 - Activity Code
◯ A
◯ B
◯ C
◯ D
◯ E
◯ F
◯ G
◯ H
◯ I
◯ J
◯ K
◯ L
◯ M
◯ N
◯ O
◯ P
◯ Q
◯ R
◯ S

Obs. 1 - Start Time ______________________

Obs. 1 - End Time ______________________

Obs. 1 - Notes ______________________

| | |
|---|---|
| Add Obs. 2 | ◯ Yes ◯ No |
| Obs. 2 - Activity Code | ◯ A<br>◯ B<br>◯ C<br>◯ D<br>◯ E<br>◯ F<br>◯ G<br>◯ H<br>◯ I<br>◯ J<br>◯ K<br>◯ L<br>◯ M<br>◯ N<br>◯ O<br>◯ P<br>◯ Q<br>◯ R<br>◯ S |
| Obs. 2 - Start Time | ________________________________ |
| Obs. 2 - End Time | ________________________________ |
| Obs. 2 - Notes | ________________________________ |
| Add Obs. 3 | ◯ Yes ◯ No |
| Obs. 3 - Activity Code | ◯ A<br>◯ B<br>◯ C<br>◯ D<br>◯ E<br>◯ F<br>◯ G<br>◯ H<br>◯ I<br>◯ J<br>◯ K<br>◯ L<br>◯ M<br>◯ N<br>◯ O<br>◯ P<br>◯ Q<br>◯ R<br>◯ S |
| Obs. 3 - End Time | ________________________________ |
| Obs. 3 - Start Time | ________________________________ |

| | |
|---|---|
| Obs. 3 - Notes | __________________________ |
| Add Obs. 4 | ◯ Yes ◯ No |
| Obs. 4 - Activity Code | ◯ A<br>◯ B<br>◯ C<br>◯ D<br>◯ E<br>◯ F<br>◯ G<br>◯ H<br>◯ I<br>◯ J<br>◯ K<br>◯ L<br>◯ M<br>◯ N<br>◯ O<br>◯ P<br>◯ Q<br>◯ R<br>◯ S |
| Obs. 4 - Start Time | __________________________ |
| Obs. 4 - End Time | __________________________ |
| Obs. 4 - Notes | __________________________ |
| Add Obs. 5 | ◯ Yes ◯ No |
| Obs. 5 - Activity Code | ◯ A<br>◯ B<br>◯ C<br>◯ D<br>◯ E<br>◯ F<br>◯ G<br>◯ H<br>◯ I<br>◯ J<br>◯ K<br>◯ L<br>◯ M<br>◯ N<br>◯ O<br>◯ P<br>◯ Q<br>◯ R<br>◯ S |
| Obs. 5 - Start Time | __________________________ |

Obs. 5 - End Time

__________________________________

Obs. 5 - Notes

__________________________________

Add Obs. 6 ◯ Yes ◯ No

Obs. 6 - Activity Code
◯ A
◯ B
◯ C
◯ D
◯ E
◯ F
◯ G
◯ H
◯ I
◯ J
◯ K
◯ L
◯ M
◯ N
◯ O
◯ P
◯ Q
◯ R
◯ S

Obs. 6 - Start Time

__________________________________

Obs. 6 - End Time

__________________________________

Obs. 6 - Notes

__________________________________

Add Obs. 7 ◯ Yes ◯ No

Obs. 7 - Activity Code
◯ A
◯ B
◯ C
◯ D
◯ E
◯ F
◯ G
◯ H
◯ I
◯ J
◯ K
◯ L
◯ M
◯ N
◯ O
◯ P
◯ Q
◯ R
◯ S

| | |
|---|---|
| Obs. 7 - Start Time | __________________________________ |
| Obs. 7 - End Time | __________________________________ |
| Obs. 7 - Notes | __________________________________ |
| Add Obs. 8 | ◯ Yes ◯ No |
| Obs. 8 - Activity Code | ◯ A<br>◯ B<br>◯ C<br>◯ D<br>◯ E<br>◯ F<br>◯ G<br>◯ H<br>◯ I<br>◯ J<br>◯ K<br>◯ L<br>◯ M<br>◯ N<br>◯ O<br>◯ P<br>◯ Q<br>◯ R<br>◯ S |
| Obs. 8 - Start Time | __________________________________ |
| Obs. 8 - End Time | __________________________________ |
| Obs. 8 - Notes | __________________________________ |
| Add Obs. 9 | ◯ Yes ◯ No |

| | |
|---|---|
| Obs. 9 - Activity Code | ◯ A<br>◯ B<br>◯ C<br>◯ D<br>◯ E<br>◯ F<br>◯ G<br>◯ H<br>◯ I<br>◯ J<br>◯ K<br>◯ L<br>◯ M<br>◯ N<br>◯ O<br>◯ P<br>◯ Q<br>◯ R<br>◯ S |
| Obs. 9 - Start Time | __________________________________ |
| Obs. 9 - End Time | __________________________________ |
| Obs. 9 - Notes | __________________________________ |
| Add Obs. 10 | ◯ Yes ◯ No |
| Obs. 10 - Activity Code | ◯ A<br>◯ B<br>◯ C<br>◯ D<br>◯ E<br>◯ F<br>◯ G<br>◯ H<br>◯ I<br>◯ J<br>◯ K<br>◯ L<br>◯ M<br>◯ N<br>◯ O<br>◯ P<br>◯ Q<br>◯ R<br>◯ S |
| Obs. 10 - Start Time | __________________________________ |
| Obs. 10 - End Time | __________________________________ |
| Obs. 10 - Notes | __________________________________ |

| | |
|---|---|
| Add Obs. 11 | ◯ Yes ◯ No |
| Obs. 11 - Activity Code | ◯ A<br>◯ B<br>◯ C<br>◯ D<br>◯ E<br>◯ F<br>◯ G<br>◯ H<br>◯ I<br>◯ J<br>◯ K<br>◯ L<br>◯ M<br>◯ N<br>◯ O<br>◯ P<br>◯ Q<br>◯ R<br>◯ S |
| Obs. 11 - Start Time | __________________________________ |
| Obs. 11 - End Time | __________________________________ |
| Obs. 11 - Notes | __________________________________ |
| Add Obs. 12 | ◯ Yes ◯ No |
| Obs. 12 - Activity Code | ◯ A<br>◯ B<br>◯ C<br>◯ D<br>◯ E<br>◯ F<br>◯ G<br>◯ H<br>◯ I<br>◯ J<br>◯ K<br>◯ L<br>◯ M<br>◯ N<br>◯ O<br>◯ P<br>◯ Q<br>◯ R<br>◯ S |
| Obs. 12 - Start Time | __________________________________ |
| Obs. 12 - End Time | __________________________________ |

| | |
|---|---|
| Obs. 12 - Notes | ______________________ |
| Add Obs. 13 | ◯ Yes ◯ No |
| Obs. 13 - Activity Code | ◯ A<br>◯ B<br>◯ C<br>◯ D<br>◯ E<br>◯ F<br>◯ G<br>◯ H<br>◯ I<br>◯ J<br>◯ K<br>◯ L<br>◯ M<br>◯ N<br>◯ O<br>◯ P<br>◯ Q<br>◯ R<br>◯ S |
| Obs. 13 - Start Time | ______________________ |
| Obs. 13 - End Time | ______________________ |
| Obs. 13 - Notes | ______________________ |
| Add Obs. 14 | ◯ Yes ◯ No |
| Obs. 14 - Activity Code | ◯ A<br>◯ B<br>◯ C<br>◯ D<br>◯ E<br>◯ F<br>◯ G<br>◯ H<br>◯ I<br>◯ J<br>◯ K<br>◯ L<br>◯ M<br>◯ N<br>◯ O<br>◯ P<br>◯ Q<br>◯ R<br>◯ S |
| Obs. 14 - Start Time | ______________________ |

| | |
|---|---|
| Obs. 14 - End Time | ________________________ |
| Obs. 14 - Notes | ________________________ |
| Add Obs. 15 | ◯ Yes ◯ No |
| Obs. 15 - Activity Code | ◯ A<br>◯ B<br>◯ C<br>◯ D<br>◯ E<br>◯ F<br>◯ G<br>◯ H<br>◯ I<br>◯ J<br>◯ K<br>◯ L<br>◯ M<br>◯ N<br>◯ O<br>◯ P<br>◯ Q<br>◯ R<br>◯ S |
| Obs. 15 - Start Time | ________________________ |
| Obs. 15 - End Time | ________________________ |
| Obs. 15 - Notes | ________________________ |
| Add Obs. 16 | ◯ Yes ◯ No |
| Obs. 16 - Activity Code | ◯ A<br>◯ B<br>◯ C<br>◯ D<br>◯ E<br>◯ F<br>◯ G<br>◯ H<br>◯ I<br>◯ J<br>◯ K<br>◯ L<br>◯ M<br>◯ N<br>◯ O<br>◯ P<br>◯ Q<br>◯ R<br>◯ S |

| | |
|---|---|
| Obs. 16 - Start Time | __________________________________ |
| Obs. 16 - End Time | __________________________________ |
| Obs. 16 - Notes | __________________________________ |
| Add Obs. 17 | ◯ Yes ◯ No |
| Obs. 17 - Activity Code | ◯ A<br>◯ B<br>◯ C<br>◯ D<br>◯ E<br>◯ F<br>◯ G<br>◯ H<br>◯ I<br>◯ J<br>◯ K<br>◯ L<br>◯ M<br>◯ N<br>◯ O<br>◯ P<br>◯ Q<br>◯ R<br>◯ S |
| Obs. 17 - Start Time | __________________________________ |
| Obs. 17 - End Time | __________________________________ |
| Obs. 17 - Notes | __________________________________ |
| Add Obs. 18 | ◯ Yes ◯ No |

| | |
|---|---|
| Obs. 18 - Activity Code | ◯ A<br>◯ B<br>◯ C<br>◯ D<br>◯ E<br>◯ F<br>◯ G<br>◯ H<br>◯ I<br>◯ J<br>◯ K<br>◯ L<br>◯ M<br>◯ N<br>◯ O<br>◯ P<br>◯ Q<br>◯ R<br>◯ S |
| Obs. 18 - Start Time | __________________________ |
| Obs. 18 - End Time | __________________________ |
| Obs. 18 - Notes | __________________________ |
| Add Obs. 19 | ◯ Yes ◯ No |
| Obs. 19 - Activity Code | ◯ A<br>◯ B<br>◯ C<br>◯ D<br>◯ E<br>◯ F<br>◯ G<br>◯ H<br>◯ I<br>◯ J<br>◯ K<br>◯ L<br>◯ M<br>◯ N<br>◯ O<br>◯ P<br>◯ Q<br>◯ R<br>◯ S |
| Obs. 19 - Start Time | __________________________ |
| Obs. 19 - End Time | __________________________ |
| Obs. 19 - Notes | __________________________ |

| | |
|---|---|
| Add Obs. 20 | ◯ Yes ◯ No |
| Obs. 20 - Activity Code | ◯ A<br>◯ B<br>◯ C<br>◯ D<br>◯ E<br>◯ F<br>◯ G<br>◯ H<br>◯ I<br>◯ J<br>◯ K<br>◯ L<br>◯ M<br>◯ N<br>◯ O<br>◯ P<br>◯ Q<br>◯ R<br>◯ S |
| Obs. 20 - Start Time | __________________________________ |
| Obs. 20 - End Time | __________________________________ |
| Obs. 20 - Notes | __________________________________ |
| Add Obs. 21 | ◯ Yes ◯ No |
| Obs. 21 - Activity Code | ◯ A<br>◯ B<br>◯ C<br>◯ D<br>◯ E<br>◯ F<br>◯ G<br>◯ H<br>◯ I<br>◯ J<br>◯ K<br>◯ L<br>◯ M<br>◯ N<br>◯ O<br>◯ P<br>◯ Q<br>◯ R<br>◯ S |
| Obs. 21 - Start Time | __________________________________ |
| Obs. 21 - End Time | __________________________________ |

Obs. 21 - Notes ________________________________

Add Obs. 22 ◯ Yes ◯ No

Obs. 22 - Activity Code
◯ A
◯ B
◯ C
◯ D
◯ E
◯ F
◯ G
◯ H
◯ I
◯ J
◯ K
◯ L
◯ M
◯ N
◯ O
◯ P
◯ Q
◯ R
◯ S

Obs. 22 - Start Time ________________________________

Obs. 22 - End Time ________________________________

Obs. 22 - Notes ________________________________

Add Obs. 23 ◯ Yes ◯ No

Obs. 23 - Activity Code
◯ A
◯ B
◯ C
◯ D
◯ E
◯ F
◯ G
◯ H
◯ I
◯ J
◯ K
◯ L
◯ M
◯ N
◯ O
◯ P
◯ Q
◯ R
◯ S

Obs. 23 - Start Time ________________________________

| | |
|---|---|
| Obs. 23 - End Time | __________________________________ |
| Obs. 23 - Notes | __________________________________ |
| Add Obs. 24 | ◯ Yes ◯ No |
| Obs. 24 - Activity Code | ◯ A<br>◯ B<br>◯ C<br>◯ D<br>◯ E<br>◯ F<br>◯ G<br>◯ H<br>◯ I<br>◯ J<br>◯ K<br>◯ L<br>◯ M<br>◯ N<br>◯ O<br>◯ P<br>◯ Q<br>◯ R<br>◯ S |
| Obs. 24 - Start Time | __________________________________ |
| Obs. 24 - End Time | __________________________________ |
| Obs. 24 - Notes | __________________________________ |
| Add Obs. 25 | ◯ Yes ◯ No |
| Obs. 25 - Activity Code | ◯ A<br>◯ B<br>◯ C<br>◯ D<br>◯ E<br>◯ F<br>◯ G<br>◯ H<br>◯ I<br>◯ J<br>◯ K<br>◯ L<br>◯ M<br>◯ N<br>◯ O<br>◯ P<br>◯ Q<br>◯ R<br>◯ S |

Obs. 25 - Start Time

______________________

Obs. 25 - End Time

______________________

Obs. 25 - Notes

______________________

Add Obs. 26

◯ Yes ◯ No

Obs. 26 - Activity Code

◯ A
◯ B
◯ C
◯ D
◯ E
◯ F
◯ G
◯ H
◯ I
◯ J
◯ K
◯ L
◯ M
◯ N
◯ O
◯ P
◯ Q
◯ R
◯ S

Obs. 26 - Start Time

______________________

Obs. 26 - End Time

______________________

Obs. 26 - Notes

______________________

Add Obs. 27

◯ Yes ◯ No

| | |
|---|---|
| Obs. 27 - Activity Code | ◯ A<br>◯ B<br>◯ C<br>◯ D<br>◯ E<br>◯ F<br>◯ G<br>◯ H<br>◯ I<br>◯ J<br>◯ K<br>◯ L<br>◯ M<br>◯ N<br>◯ O<br>◯ P<br>◯ Q<br>◯ R<br>◯ S |
| Obs. 27 - Start Time | __________________ |
| Obs. 27 - End Time | __________________ |
| Obs. 27 - Notes | __________________ |
| Add Obs. 28 | ◯ Yes ◯ No |
| Obs. 28 - Activity Code | ◯ A<br>◯ B<br>◯ C<br>◯ D<br>◯ E<br>◯ F<br>◯ G<br>◯ H<br>◯ I<br>◯ J<br>◯ K<br>◯ L<br>◯ M<br>◯ N<br>◯ O<br>◯ P<br>◯ Q<br>◯ R<br>◯ S |
| Obs. 28 - Start Time | __________________ |
| Obs. 28 - End Time | __________________ |
| Obs. 28 - Notes | __________________ |

| | |
|---|---|
| Add Obs. 29 | ◯ Yes ◯ No |
| Obs. 29 - Activity Code | ◯ A<br>◯ B<br>◯ C<br>◯ D<br>◯ E<br>◯ F<br>◯ G<br>◯ H<br>◯ I<br>◯ J<br>◯ K<br>◯ L<br>◯ M<br>◯ N<br>◯ O<br>◯ P<br>◯ Q<br>◯ R<br>◯ S |
| Obs. 29 - Start Time | __________________________________ |
| Obs. 29 - End Time | __________________________________ |
| Obs. 29 - Notes | __________________________________ |
| Add Obs. 30 | ◯ Yes ◯ No |
| Obs. 30 - Activity Code | ◯ A<br>◯ B<br>◯ C<br>◯ D<br>◯ E<br>◯ F<br>◯ G<br>◯ H<br>◯ I<br>◯ J<br>◯ K<br>◯ L<br>◯ M<br>◯ N<br>◯ O<br>◯ P<br>◯ Q<br>◯ R<br>◯ S |
| Obs. 30 - Start Time | __________________________________ |
| Obs. 30 - End Time | __________________________________ |

Obs. 30 - Notes

______________________________

Comments:

______________________________________

#### Audit Trail

Name of Data Entry Person

______________________________

Date of Data Entry

______________________________
(Kindly change the date every time a change is made to the data)

### Livestock Form LS

| | |
|---|---|
| LS01. What happens to your household garbage? | ◯ Municipal waste<br>◯ Burnt<br>◯ In the yard<br>◯ Dump<br>◯ Other |
| Specify Other | __________________________________ |
| LS02. Does the household own, keep or look after any animals (cats, dogs, cattle, chickens, ducks etc.)? | ◯ Yes<br>◯ No |
| LS03. Do your animals have access to the household garbage? | ◯ Yes<br>◯ No |
| LS04. What domestic animal/s does the household own, keep or look after? | ☐ Cattle<br>☐ Fowl (eg. Chickens, geese, duck)<br>☐ Pig<br>☐ Sheep<br>☐ Goat<br>☐ Horse<br>☐ Donkey<br>☐ Cat<br>☐ Dog<br>☐ Pigeon<br>☐ Other |
| Cattle Count | __________________________________ |
| Fowl Count | __________________________________ |
| Pig Count | __________________________________ |
| Sheep Count | __________________________________ |
| Goat Count | __________________________________ |
| Horse Count | __________________________________ |
| Donkey Count | __________________________________ |
| Cat Count | __________________________________ |
| Dog Count | __________________________________ |

Pigeon Count

__________________________________

Specify Other

__________________________________

Other Count

__________________________________

LS05. Do any animals live or stay overnight inside the house where other people sleep?

- ◯ Yes
- ◯ No

LS05. Tick all the animals that sleep in the house

- ☐ Cattle
- ☐ Fowl (eg. Chickens, geese, duck)
- ☐ Pig
- ☐ Sheep
- ☐ Goat
- ☐ Horse
- ☐ Donkey
- ☐ Cat
- ☐ Dog
- ☐ Pigeon
- ☐ Other

Specify Other

__________________________________

LS06. Do any of the animals live or stay overnight in the yard or kraal around the household?

- ◯ Yes
- ◯ No

LS06. Tick all those that sleep in the yard or kraal around the household

- ☐ Cattle
- ☐ Fowl (eg. Chickens, geese, duck)
- ☐ Pig
- ☐ Sheep
- ☐ Goat
- ☐ Horse
- ☐ Donkey
- ☐ Cat
- ☐ Dog
- ☐ Pigeon
- ☐ Other

Specify Other

__________________________________

LS07. Do any of the animals sleep in a field or kraal far from the household?

- ◯ Yes
- ◯ No

| | |
|---|---|
| LS07. Tick all the animals that sleep in a field far from the household | ☐ Cattle<br>☐ Fowl (eg. Chickens, geese, duck)<br>☐ Pig<br>☐ Sheep<br>☐ Goat<br>☐ Horse<br>☐ Donkey<br>☐ Cat<br>☐ Dog<br>☐ Pigeon<br>☐ Other |
| Specify Other | __________________________________ |
| L08. How far is the closest kraal from where people sleep and/or eat? | ◯ 1-10 metres<br>◯ 11-50 metres<br>◯ 51-100 metres<br>◯ 101-500 metres<br>◯ .501-1km<br>◯ +1km<br>◯ Not Applicable |

**Only answer this section for those households that report owning at least one of the following animals: poultry, cattle, sheep, pigs, goat**

| | |
|---|---|
| LS09. Who looks after the animal/s | ☐ Animals looked after by someone who is not a household member |
| LS09. Who looks after the animal/s 1) | __________________________________ |
| LS09. Who looks after the animal/s 2) | __________________________________ |
| LS09. Who looks after the animal/s 3) | __________________________________ |
| LS09. Who looks after the animal/s 4) | __________________________________ |
| LS10. In the last 6 months have any of your animals been slaughtered/killed? | ◯ Yes<br>◯ No |
| LS11. Tick all the animals that were slaughtered/killed in the and indicate how many of each animal were slaughtered/killed | ☐ Cattle<br>☐ Sheep<br>☐ Pig<br>☐ Goat<br>☐ Fowls (eg. Chickens, geese, ducks)<br>☐ Pigeon |
| Cattle slaughtered/killed | __________________________________ |
| Sheep slaughtered/killed | __________________________________ |

Pig slaughtered/killed

__________________________________

Goat slaughtered/killed

__________________________________

Fowls slaughtered/killed

__________________________________

Pigeon slaughtered/killed

__________________________________

LS12. In the last 6 months, were any of the animals slaughtered/killed at the household?

◯ Yes
◯ No

List the household members that slaughtered and/or assisted in slaughtering the animals

☐ Animals slaughtered by someone who is not a household member

LS13. List the household members that slaughtered and/or assisted in slaughtering the animals
1)

__________________________________

LS13. List the household members that slaughtered and/or assisted in slaughtering the animals
2)

__________________________________

LS13. List the household members that slaughtered and/or assisted in slaughtering the animals
3)

__________________________________

LS13. List the household members that slaughtered and/or assisted in slaughtering the animals
4)

__________________________________

LS13. List the household members that slaughtered and/or assisted in slaughtering the animals
5)

__________________________________

LS14. Tick all the animals that were slaughtered at the household in the last 6 months

☐ Cattle
☐ Sheep
☐ Pig
☐ Goat
☐ Fowls (eg. Chickens, geese, ducs)
☐ Pigeon
☐ Not Applicable

LS15. Were any of the animals taken to a slaughter house?

☐ Cattle
☐ Sheep
☐ Pig
☐ Goat
☐ Fowls (eg. Chickens, geese, ducs)
☐ Pigeon
☐ Not Applicable

| | |
|---|---|
| LS16. What happened with the slaughtered/killed animal? | ◯ It was eaten by the household members<br>◯ It was sold for money<br>◯ It was thrown away/disposed of<br>◯ It was shared with the community<br>◯ Ritual purposes<br>◯ Other |
| Specify Other | __________________________________ |
| LS17. Do you and members in your household drink milk that comes from your cows/goat/sheep? | ◯ Yes<br>◯ No |
| LS18. If 'Yes', how do you drink the milk? | ◯ Raw milk - no heating or preparation<br>◯ Boil milk for a short time (a few minutes)<br>◯ Boil milk for a long time (around half an hour)<br>◯ Other |
| Specify Other | __________________________________ |
| LS19. Do you know if any of your animals have been tested for Tuberculosis in the past 12 months? | ◯ Yes<br>◯ No |
| LS20. Tick all the animals that were tested for TB in the last 12 months | ☐ Cattle<br>☐ Pig<br>☐ Sheep<br>☐ Goat<br>☐ Horse<br>☐ Donkey<br>☐ Cat<br>☐ Dog<br>☐ Other |
| Specify Other | __________________________________ |

#### Animals with Sickness and Disease

| | |
|---|---|
| LS21. In the last 6 months have any of your animals had to go to the vet for sickness or disease? | ◯ Yes<br>◯ No |
| LS22. Have any of your animals and any of these symptoms: | ☐ |
| LS23. In the last 6 months have any of your animals died because of sickness or disease? | ◯ Yes<br>◯ No |

LS24. Tick all the animals that in last 6 months died because of sickness or disease and indicate how many of each animal died

- ☐ Cattle
- ☐ Fowls (eg. Chickens, geese, ducks)
- ☐ Pig
- ☐ Sheep
- ☐ Goat
- ☐ Horse
- ☐ Donkey
- ☐ Cat
- ☐ Dog
- ☐ Pigeon
- ☐ Other

Specify Other ______________________________

Cattle Count ______________________________

Fowl Count ______________________________

Pig Count ______________________________

Sheep Count ______________________________

Goat Count ______________________________

Horse Count ______________________________

Donkey Count ______________________________

Cat Count ______________________________

Dog Count ______________________________

Pigeon Count ______________________________

[ls24_other] Count ______________________________

LS24. What happened to the meat of the animal that died?

- ◯ It was eaten
- ◯ It was sold
- ◯ It was given to people
- ◯ It was thrown out
- ◯ Other

Specify Other ______________________________

| | |
|---|---|
| LS25. Would you consider having a small skin test done on all of your animals? | ◯ Yes<br>◯ No |
| LS26. If 'No', why not? | ____________________________________ |
| LS27. On average, how much would you sell each of your animals to us if we wanted to buy it? | ☐ Cattle<br>☐ Fowls (eg. Chickens, geese, ducks)<br>☐ Pig<br>☐ Sheep<br>☐ Goat<br>☐ Horse<br>☐ Donkey<br>☐ Cat<br>☐ Dog<br>☐ Pigeon<br>☐ Other |
| Specify Other | ______________________________ |
| Cattle Selling Price | ______________________________<br>(R (ZAR)) |
| Fowl Selling Price | ______________________________<br>(R (ZAR)) |
| Pig Selling Price | ______________________________<br>(R (ZAR)) |
| Sheep Selling Price | ______________________________<br>(R (ZAR)) |
| Goat Selling Price | ______________________________<br>(R (ZAR)) |
| Horse Selling Price | ______________________________<br>(R (ZAR)) |
| Donkey Selling Price | ______________________________<br>(R (ZAR)) |
| Cat Selling Price | ______________________________<br>(R (ZAR)) |
| Dog Selling Price | ______________________________<br>(R (ZAR)) |

| | |
|---|---|
| Pigeon Selling Price | ____________ (R (ZAR)) |
| [ls27_other] Selling Price | ____________ (R (ZAR)) |

**Only answer this section for those households that do NOT own any animals**

| | |
|---|---|
| LS28. Do you and members in your household ever drink raw (unpasteurized, un-boiled milk)? | ◯ Yes<br>◯ No |
| LS29: If yes, how often | ◯ Always<br>◯ Very Often<br>◯ Sometimes<br>◯ Rarely |
| LS30. Do you and members in your household ever buy meat from an informal butcher in the area? | ◯ Yes<br>◯ No |
| LS31: If yes, ask how often | ◯ Always<br>◯ Very Often<br>◯ Sometimes<br>◯ Rarely |
| LS32. Do any of the household members look after other people's/neighbours/community members' animals (example Herding cattle)? | ◯ Yes<br>◯ No |
| LS33. In the last 6 months, has anyone in the household attended a function (funeral, wedding) where an animal was slaughtered and the meat was eaten? | ◯ Yes<br>◯ No |
| How many functions | ____________ |
| LS34. In the last 6 months, have your neighbours/community members slaughtered any animals? | ◯ Yes<br>◯ No |
| LS35. In the last 6 months, have you eaten any meat that you got/were given from your neighbours/community members after they slaughtered the animal? | ◯ Yes<br>◯ No |
| LS36: Date form completed | ____________ |
| LS37: Name of Person completing this form | ____________ |

#### Audit Trail

Name of Data Entry Person

________________________________

Date of Data Entry

________________________________
(Kindly change the date every time a change is made to the data)
