## Supplementary material for "Household contact tracing with intensified tuberculosis and HIV screening in South Africa: a cluster randomised trial": S3 Checklist

**S3 Checklist: CONSORT 2010 checklist of information to include when reporting a cluster randomised trial**

| Section/Topic | Item No | Standard Checklist item | Extension for cluster designs | Section/Paragraph No * |
| --- | --- | --- | --- | --- |
| Title and abstract | | | |  |
|  | 1a | Identification as a randomised trial in the title | Identification as a cluster randomised trial in the title | Title |
|  | 1b | Structured summary of trial design, methods, results, and conclusions (for specific guidance see CONSORT for abstracts)^[1],^^[2]^ | See table 2 | Abstract |
| Introduction | | | |  |
| Background and objectives | 2a | Scientific background and explanation of rationale | Rationale for using a cluster design | Introduction, Paragraph 1 |
|  | 2b | Specific objectives or hypotheses | Whether objectives pertain to the the cluster level, the individual participant level or both | Introduction, Paragraph 3 |
| Methods | | | |  |
| Trial design | 3a | Description of trial design (such as parallel, factorial) including allocation ratio | Definition of cluster and description of how the design features apply to the clusters | Methods, Paragraph 1 |
|  | 3b | Important changes to methods after trial commencement (such as eligibility criteria), with reasons |  | Methods, Paragraph 12 |
| Participants | 4a | Eligibility criteria for participants | Eligibility criteria for clusters | Methods, Paragraph 2 |
|  | 4b | Settings and locations where the data were collected |  | Methods, Paragraph 1 |
| Interventions | 5 | The interventions for each group with sufficient details to allow replication, including how and when they were actually administered | Whether interventions pertain to the cluster level, the individual participant level or both | Methods, Paragraph 5-9 |
| Outcomes | 6a | Completely defined pre-specified primary and secondary outcome measures, including how and when they were assessed | Whether outcome measures pertain to the cluster level, the individual participant level or both | Methods, Paragraph 11-12 |
|  | 6b | Any changes to trial outcomes after the trial commenced, with reasons |  | Methods, Paragraph 12. |
| Sample size | 7a | How sample size was determined | Method of calculation, number of clusters(s) (and whether equal or unequal cluster sizes are assumed), cluster size, a coefficient of intracluster correlation (ICC or *k*), and an indication of its uncertainty | Methods, Paragraph 13 |
|  | 7b | When applicable, explanation of any interim analyses and stopping guidelines |  | NA |
| Randomisation: | | | |  |
| Sequence generation | 8a | Method used to generate the random allocation sequence |  | Methods, Paragraph 4 |
|  | 8b | Type of randomisation; details of any restriction (such as blocking and block size) | Details of stratification or matching if used | Methods, Paragraph 4 |
| Allocation concealment mechanism | 9 | Mechanism used to implement the random allocation sequence (such as sequentially numbered containers), describing any steps taken to conceal the sequence until interventions were assigned | Specification that allocation was based on clusters rather than individuals and whether allocation concealment (if any) was at the cluster level, the individual participant level or both | Methods, Paragraph 4 |
| Implementation | 10 | Who generated the random allocation sequence, who enrolled participants, and who assigned participants to interventions | Replace by 10a, 10b and 10c | Methods, Paragraph 4 |
|  | 10a |  | Who generated the random allocation sequence, who enrolled clusters, and who assigned clusters to interventions | Methods, Paragraph 4 |
|  | 10b |  | Mechanism by which individual participants were included in clusters for the purposes of the trial (such as complete enumeration, random sampling) | Methods, Paragraph 3 |
|  | 10c |  | From whom consent was sought (representatives of the cluster, or individual cluster members, or both), and whether consent was sought before or after randomisation | Methods, Paragraph 5 |
| Blinding | 11a | If done, who was blinded after assignment to interventions (for example, participants, care providers, those assessing outcomes) and how |  | Methods, Paragraph 4 |
|  | 11b | If relevant, description of the similarity of interventions |  | Methods, Paragraphs 6-10 |
| Statistical methods | 12a | Statistical methods used to compare groups for primary and secondary outcomes | How clustering was taken into account | Methods, Paragraph 17 |
|  | 12b | Methods for additional analyses, such as subgroup analyses and adjusted analyses |  | Methods, Paragraph 17 |
| Results | | | |  |
| Participant flow (a diagram is strongly recommended) | 13a | For each group, the numbers of participants who were randomly assigned, received intended treatment, and were analysed for the primary outcome | For each group, the numbers of clusters that were randomly assigned, received intended treatment, and were analysed for the primary outcome | Results, Paragraph 1-3, Figure 1. |
|  | 13b | For each group, losses and exclusions after randomisation, together with reasons | For each group, losses and exclusions for both clusters and individual cluster members | Results, Paragraph 4, Figure 1. |
| Recruitment | 14a | Dates defining the periods of recruitment and follow-up |  | Results, Paragraph 1 |
|  | 14b | Why the trial ended or was stopped |  |  |
| Baseline data | 15 | A table showing baseline demographic and clinical characteristics for each group | Baseline characteristics for the individual and cluster levels as applicable for each group | Results, Paragraphs 2-3. Tables 1, 2 |
| Numbers analysed | 16 | For each group, number of participants (denominator) included in each analysis and whether the analysis was by original assigned groups | For each group, number of clusters included in each analysis | Results, Paragraph 4, Figure 1. |
| Outcomes and estimation | 17a | For each primary and secondary outcome, results for each group, and the estimated effect size and its precision (such as 95% confidence interval) | Results at the individual or cluster level as applicable and a coefficient of intracluster correlation (ICC or k) for each primary outcome | Results, Table 3 |
|  | 17b | For binary outcomes, presentation of both absolute and relative effect sizes is recommended |  | Not done |
| Ancillary analyses | 18 | Results of any other analyses performed, including subgroup analyses and adjusted analyses, distinguishing pre-specified from exploratory |  | Results, Paragraph 6, 11 |
| Harms | 19 | All important harms or unintended effects in each group (for specific guidance see CONSORT for harms^[3]^) |  | NA |
| Discussion | | | |  |
| Limitations | 20 | Trial limitations, addressing sources of potential bias, imprecision, and, if relevant, multiplicity of analyses |  | Discussion, Paragraph 2 |
| Generalisability | 21 | Generalisability (external validity, applicability) of the trial findings | Generalisability to clusters and/or individual participants (as relevant) | Discussion, Paragraph 7 |
| Interpretation | 22 | Interpretation consistent with results, balancing benefits and harms, and considering other relevant evidence |  | Discussion, Paragraph 9 |
| Other information | | |  |  |
| Registration | 23 | Registration number and name of trial registry |  | Abstract |
| Protocol | 24 | Where the full trial protocol can be accessed, if available |  | S1 Text (Protocol) |
| Funding | 25 | Sources of funding and other support (such as supply of drugs), role of funders |  | Sources of Funding section |

** Note: page numbers optional depending on journal requirements*

**Table 2:	Extension of CONSORT for abstracts**1**^,^**2 **to reports of cluster randomised trials**

| Item | Standard Checklist item | Extension for cluster trials |
| --- | --- | --- |
| Title | Identification of study as randomised | Identification of study as cluster randomised |
| Trial design | Description of the trial design (e.g. parallel, cluster, non-inferiority) |  |
| Methods |  |  |
| Participants | Eligibility criteria for participants and the settings where the data were collected | Eligibility criteria for clusters |
| Interventions | Interventions intended for each group |  |
| Objective | Specific objective or hypothesis | Whether objective or hypothesis pertains to the cluster level, the individual participant level or both |
| Outcome | Clearly defined primary outcome for this report | Whether the primary outcome pertains to the cluster level, the individual participant level or both |
| Randomization | How participants were allocated to interventions | How clusters were allocated to interventions |
| Blinding (masking) | Whether or not participants, care givers, and those assessing the outcomes were blinded to group assignment |  |
| Results |  |  |
| Numbers randomized | Number of participants randomized to each group | Number of clusters randomized to each group |
| Recruitment | Trial status^[4]^ |  |
| Numbers analysed | Number of participants analysed in each group | Number of clusters analysed in each group |
| Outcome | For the primary outcome, a result for each group and the estimated effect size and its precision | Results at the cluster or individual participant level as applicable for each primary outcome |
| Harms | Important adverse events or side effects |  |
| Conclusions | General interpretation of the results |  |
| Trial registration | Registration number and name of trial register |  |
| Funding | Source of funding |  |
