## Supplementary material for "Household contact tracing with intensified tuberculosis and HIV screening in South Africa: a cluster randomised trial": S4 Table

### S4 Table: Effect of intervention versus standard of care on primary trial outcome, sensitivity analyses

| **Outcome** | **Standard of care** | **Household intervention** | **Hazard ratio (95% CI)** |
| --- | --- | --- | --- |
| **Among all household contacts including those not present at baseline, any TB** | | | |
| Contacts diagnosed with TB | 36/2723 (1.3%) | 51/3301 (1.5%) | 1.18 (0.75, 1.88) |
| Contact deaths | 50/4132 (1.2%) | 46/4361 (1.1%) | 0.79 (0.52, 1.20) |
| TB or death | 86/2773 (3.1%) | 97/3347 (2.9%) | 0.91 (0.67, 1.23) |
| **Among household contacts present at baseline, bacteriologically-confirmed TB** | | | |
| Contacts diagnosed with TB | 16/2551 (0.6%) | 23/3188 (0.7%) | 1.17 (0.60, 2.28) |
| Contact deaths | 49/3961 (1.2%) | 42/4242 (1.0%) | 0.72 (0.47, 1.10) |
| TB or death | 65/2600 (2.5%) | 65/3230 (2.0%) | 0.76 (0.53, 1.09) |
| **Among all household contacts including those not present at baseline, bacteriologically-confirmed TB** | | | |
| Contacts diagnosed with TB | 19/2723 (0.7%) | 23/3301 (0.7%) | 1.01 (0.54, 1.91) |
| Contact deaths | 50/4132 (1.2%) | 46/4361 (1.1%) | 0.79 (0.52, 1.20) |
| TB or death | 69/2773 (2.5%) | 69/3347 (2.1%) | 0.79 (0.56, 1.11) |
