## Supplementary material for "Household contact tracing with intensified tuberculosis and HIV screening in South Africa: a cluster randomised trial": S5 Table

### S5 Table: Subgroup analyses of primary and secondary outcomes by study site

|  | **Mangaung** | | | **Capricorn** | | | **Interaction p-value^1^** |
| --- | --- | --- | --- | --- | --- | --- | --- |
| **Outcome** | **Standard of care** | **Household Intervention** | **Effect estimate** | **Standard of care** | **Household Intervention** | **Effect estimate** |  |
| **Primary outcome** |  |  | **Hazard ratio (95% CI)** |  |  | **Hazard ratio (95% CI)** |  |
| Contacts diagnosed with TB | 21/1263 (1.7%) | 36/1487 (2.4%) | 1.48 (0.83, 2.67) | 10/1288 (0.8%) | 15/1701 (0.9%) | 1.16 (0.51, 2.65) | 0.64 |
| Contact deaths | 16/1853 (0.9%) | 22/1958 (1.1%) | 1.08 (0.54, 2.15) | 33/2108 (1.6%) | 20/2281 (0.9%) | 0.56 (0.32, 0.97) | 0.15 |
| TB or death | 37/1279 (2.9%) | 58/1509 (3.8%) | 1.26 (0.81, 1.97) | 43/1321 (3.3%) | 35/1721 (2.0%) | 0.63 (0.40, 1.00) | 0.03 |
| **Secondary outcomes** |  |  | **Odds ratio (95% CI)** |  |  | **Odds ratio (95% CI)** |  |
| Prevalence of TST positivity (≥10mm) among children ≤14 years | 14/370 (3.8%) | 37/464 (8.0%) | 2.05 (0.95, 4.44) | 1/430 (0.2%) | 1/381 (0.3%) | 1.13 (0.07, 18.04) | 0.66 |
| Prevalence of undiagnosed or untreated HIV infection | 26/1260 (2.1%) | 25/1486 (1.7%) | 0.81 (0.47, 1.41) | 6/1283 (0.5%) | 16/1699 (0.9%) | 2.02 (0.79, 5.18) | 0.10 |

^1^P-value calculated from Wald test of interaction term
