## Supplementary material for "Household contact tracing with intensified tuberculosis and HIV screening in South Africa: a cluster randomised trial": S6 Table

### S6 Table: Subgroup analyses of prevalence of TST positivity, by age of child

|  | **Children<5 years** | | | **Children ≥5 years** | | |  |
| --- | --- | --- | --- | --- | --- | --- | --- |
| **Outcome** | **Standard of care** | **Household Intervention** | **Odds ratio (95% CI)** | **Standard of care** | **Household Intervention** | **Odds ratio (95% CI)** | **Interaction p-value^1^** |
| Prevalence of TST positivity (≥10mm) among children ≤14 years | 7/185 (3.8%) | 14/229 (6.1%) | 1.51 (0.53, 4.33) | 8/615 (1.3%) | 24/616 (3.9%) | 2.62 (1.09, 6.33) | 0.39 |

^1^P-value calculated from Wald test of interaction term
