## Supplementary material for "Household contact tracing with intensified tuberculosis and HIV screening in South Africa: a cluster randomised trial": S7 Checklist

**S4 Checklist: CONSERVE-CONSORT Checklist**

| **CONSERVE-CONSORT** | | | | | |
| --- | --- | --- | --- | --- | --- |
| **Item** | **Item Title** | **Description** | | | **Section/Paragraph Number** |
| I. | Extenuating Circumstances | Describe the circumstances and how they constitute extenuating circumstances. | | | Methods, Paragraph 12 |
| II. | Important Modifications | a. Describe how the modifications are important modifications. | | | Methods, Paragraph 14 |
|  |  | b. Describe the impacts and mitigating strategies, including their rationale and implications for the trial. | | | Methods, Paragraph 14 |
|  |  | c. Provide a modification timeline. | | | Methods, Paragraph 14 |
| III. | Responsible Parties | State who planned, reviewed and approved the modifications. | | | Methods, Paragraph 14 |
| IV. | Interim data | If modifications were informed by trial data, describe how the interim data were used, including whether they were examined by study group, and whether the individuals reviewing the data were blinded to the treatment allocation. | | | Methods, Paragraph 14 |
| **CONSORT Number and Item** | | For each row, if important modifications occurred check “direct impact” and/or “mitigating strategy” and describe the changes in the trial manuscript or supplement. Check “no change” for items that are unaffected in the extenuating circumstance. | | | **Section/Paragraph Number** |
|  |  | **No Change** | **Impact*** | **Mitigating Strategy*** |  |
| 1 | Title and abstract | X |  |  |  |
| 2 | Introduction | X |  |  |  |
| 3 | Methods: Trial design |  | X |  | Methods, Paragraphs 12, 14 |
| 4 | Methods: Participants | X |  |  |  |
| 5 | Methods: Interventions | X |  |  |  |
| 6 | Methods: Outcomes | X |  |  |  |
| 7 | Methods: Sample Size |  | X |  | Methods, Paragraphs 12, 14 |
| 8-10 | Methods: Randomisation | X |  |  |  |
| 11 | Methods: Blinding | X |  |  |  |
| 12 | Methods: Statistical Methods | X |  |  |  |
| 13 | Results: Participant Flow | X |  |  |  |
| 14 | Results: Recruitment | X |  |  |  |
| 15 | Results: Baseline Data | X |  |  |  |
| 16 | Results: Numbers analysed | X |  |  |  |
| 17 | Results: Outcomes and estimation | X |  |  |  |
| 18 | Results: Ancillary analysis | X |  |  |  |
| 19 | Results: Harms | X |  |  |  |
| 20 | Discussion: Limitations |  |  | X | Discussion, Paragraph 2 |
| 21 | Discussion: Generalisability | x |  |  |  |
| 22 | Other information: Registration | X |  |  |  |
| 23 | Other information: Protocol | X |  |  |  |
| 24 | Other information: Funding | X |  |  |  |
| *Aspects of the trial that are directly affected or changed by the extenuating circumstance and are not under the control of investigators, sponsor or funder. **Aspects of the trial that are modified by the study investigators, sponsor or funder to respond to the extenuating circumstance or manage the direct impacts on the trial.  The CONSERVE-CONSORT Checklist is licensed by the CONSERVE Group under the Creative Commons Attribution-NonCommercial-NoDerivs 4.0 International license. | | | | | |
